# Voice patterns as markers of schizophrenia: building a cumulative generalizable approach via a cross-linguistic and meta-analysis based investigation

**DOI:** 10.1101/2022.04.03.22273354

**Authors:** Alberto Parola, Arndis Simonsen, Jessica Mary Lin, Yuan Zhou, Huiling Wang, Shiho Ubukata, Katja Koelkebeck, Vibeke Bliksted, Riccardo Fusaroli

**Affiliations:** Department of Linguistics, Cognitive Science and Semiotics, Aarhus University, Aarhus, Denmark; The Interacting Minds Center - Institute of Culture and Society, Aarhus University, Aarhus, Denmark; Department of Psychology, University of Turin, Turin, Italy; Psychosis Research Unit - Department of Clinical Medicine, Aarhus University, Aarhus, Denmark; Institute of Psychology, Chinese Academy of Sciences, Beijing, China; Department of Psychiatry, Renmin Hospital of Wuhan University, Wuhan, China; Department of Psychiatry, Kyoto University, Kyoto, Japan; LVR-Hospital Essen, Department of Psychiatry and Psychotherapy, Hospital and Institute of the University of Duisburg-Essen, Essen, Germany; Center for Translational Neuro- & Behavioral Sciences (C-TNBS), University Duisburg-Essen, Germany; Linguistic Data Consortium, University of Pennsylvania, USA

**Keywords:** vocal analysis, psychosis, speech signal, digital phenotyping, prosody, negative symptoms

## Abstract

**Background and Hypothesis:** Voice atypicalities are potential markers of clinical features of schizophrenia (e.g., negative symptoms). A recent meta-analysis identified an acoustic profile associated with schizophrenia (reduced pitch variability and increased pauses), but also highlighted shortcomings in the field: small sample sizes, little attention to the heterogeneity of the disorder, and to generalizing findings to diverse samples and languages.

**Study Design:** We provide a critical cumulative approach to vocal atypicalities in schizophrenia, where we conceptually and statistically build on previous studies. We aim at identifying a cross-linguistically reliable acoustic profile of schizophrenia and assessing sources of heterogeneity (symptomatology, pharmacotherapy, clinical and social characteristics). We relied on previous meta-analysis to build and analyze a large cross-linguistic dataset of audio recordings of 231 patients with schizophrenia and 238 matched controls (>4.000 recordings in Danish, German, Mandarin and Japanese). We used multilevel Bayesian modeling, contrasting meta-analytically informed and skeptical inferences.

**Study Results:** We found only a minimal generalizable acoustic profile of schizophrenia (reduced pitch variability), while duration atypicalities replicated only in some languages. We identified reliable associations between acoustic profile and individual differences in clinical ratings of negative symptoms, medication, age and gender. However, these associations vary across languages.

**Conclusions:** The findings indicate that a strong cross-linguistically reliable acoustic profile of schizophrenia is unlikely. Rather, if we are to devise effective clinical applications able to target different ranges of patients, we need first to establish larger and more diverse cross-linguistic datasets, focus on individual differences, and build self-critical cumulative approaches.

## Introduction

From its very first definitions, schizophrenia has been associated with voice atypicalities^1,2^, qualitatively described in terms of e.g., poverty of speech, increased pauses, distinctive tone and intensity of voice. A recent systematic meta-analysis^3^ indicates a plausible acoustic profile associated with schizophrenia. In this study, we assess how generalizable that profile is within a large cross-linguistic dataset, as well as sources of heterogeneity in vocal patterns of patients with schizophrenia.

Atypical voice patterns are included amongst negative symptoms of schizophrenia, such as alogia and blunted affect, which are among the primary diagnostic criteria and prognostic indicators of the disorder (e.g. response to treatment and reduced likelihood of remission^4–7^). Vocal behavior may constitute a window into the underlying social and cognitive features of the disorder^8^. For example, the social and cognitive impairments frequently reported in schizophrenia^9–11^ may be reflected in difficulties in speech fluency (e.g. increased pauses), or in controlling the voice to express affective and emotional contents and to mark relevant information^12,13^. In other words, not only can the quantitative analysis of vocal behavior scaffold the current evaluation of negative symptoms^14–19^, but it could also offer a more fine-grained perspective on its clinical, social and cognitive dimensions, e.g., social and cognitive functioning over time^8,18,20,21^.

However, while extensive literature exists on vocal atypicalities in schizophrenia, with studies spanning back to the 60’s, the findings are often contradictory and difficult to interpret. A recent meta-analysis^3^ identified weak atypicalities in pitch variability - potentially related to flat affect- and stronger atypicalities in proportion of spoken time, speech rate, and pauses - potentially related to alogia and flat affect. The effects had large heterogeneity, and were modest compared to clinical judgments of vocal atypicalities^22^. The studies were noted to have small sample sizes, high heterogeneity in methods and features analyzed, with little to no attention to the heterogeneity of the disorder^23–25^ and to the replicability and generalizability of previous results on diverse samples^20,25–27^. Further, voice quality features - highlighted as important by speech pathologists and speech processing research^28,29^-had been largely neglected.

To assess the robustness and potential clinical impact of biobehavioral vocal markers of schizophrenia, we need to understand under which conditions they vary and under which they can be relied upon. We need to map which variations in clinical and socio-cognitive features might underlie voice atypicalities, and how stable they are across languages, time and recordings. For example, although vocal behavior has been shown to be influenced by linguistic and cultural factors, all studies of vocal markers of schizophrenia have investigated single monolingual samples. We need to assess how voice atypicalities relate to the development of the disorder, for instance, whether they are a long-term consequence of chronicity, already present at illness onset, and vary along with symptom severity, thus being potentially useful for tracking the development of the disorder and monitoring the symptomatology over time^30–32^. Another crucial issue is how antipsychotic drugs relate to these atypicalities and impact our ability to use them as biobehavioral markers. For example, effects of antipsychotic medication have been hypothesized to affect language in different ways, such as causing extrapyramidal motor symptoms or increasing negative symptoms by blocking dopamine receptors^33^.

In other words, there is a need for a more rigorous cumulative scientific approach to understand vocal and prosodic atypicalities in schizophrenia: the synthesis and integration of data across studies and laboratories, in order to assess which patterns might generalize across contexts and samples, identify possible sources of variation and improve estimations. In this work, we provide the first steps towards such an approach.

First, we made use of the recommendations developed in the previous meta-analysis^3^ and collected a large dataset of multiple audio-recordings of patients with schizophrenia and controls in four different languages and three language families (Germanic, Mandarin-Chinese, Japonic). Such a setup provides a stronger basis for estimating the robustness of vocal atypicalities, within and between subjects and samples, as well as their variability. Further, this design allows us to explicitly compare results across different languages for the first time in schizophrenia, thus accounting for the natural differences in vocal patterns across languages.

Second, we provided a more systematic investigation of the acoustic features potentially associated with schizophrenia. While a previous meta-analysis^3^ identified relevant estimates for a limited set of features (n = 8), pertaining to pitch and rhythm, recent investigations into Parkinson’s disease and depression^28,34^ suggest that voice quality and articulatory features might have higher discriminative power compared to traditional acoustic features. In other words, they might be particularly involved in the mechanisms underlying the disorders. We therefore extended the acoustic features investigated to include them.

Third, we cumulatively build on previous findings by explicitly (but critically) including the meta-analytic findings in the statistical Bayesian analysis of the current study. This practice - referred to as “informed priors” or “posterior passing”^35^ - allows us to directly estimate how well our results match previous findings and in which ways they deviate, but also potentially increases the precision of our estimates.

Fourth, we include a more comprehensive assessment of the patients’ symptomatology and clinical profile. Specifically, we model the relationships between acoustic features and pharmacotherapy, relevant clinical aspects, demographic and social features, relationships that have rarely been jointly investigated in previous studies^36^.

Finally, we rely on an open methodology: not only do we carefully describe the methodology used and test the robustness of the results to variations in the methodology; but we also use open-source software, extracting the features in a reproducible manner with openly available scripts.

By providing an initial consolidation and test of acoustic atypicalities in schizophrenia that systematically builds on and extends the previous literature, we aim to set the foundations for more critical theory development, more cumulative and holistic approaches to the understanding of the underlying mechanisms and potentially the development of applications that constructively support clinical practices.

## Methods

### Participants

We collected a Danish (DK), German (GE), Chinese (CH), and Japanese (JP) cross-linguistic dataset involving 231 participants with schizophrenia (105 DK, 61GE, 51CH, 14JP) and 238 matched controls (HC) (116DK, 62GE, 43CH, 17JP). The samples for the present study were collected in separate studies assessing mentalizing ability in patients with schizophrenia and healthy controls. Information on demographics, IQ, psychopathology, and social functioning is summarized in **Table 1**. Detailed information on each study is reported in the **Supplementary Material (SM)** - **S1**.

**Table 1.** Demographic and clinical characteristics of patients with schizophrenia and healthy controls (HC).

| Corpus | Danish |  | German |  | Chinese |  | Japanese |  |
| --- | --- | --- | --- | --- | --- | --- | --- | --- |
| **Diagnosis** | SCZ<br>N = 105 | HC<br>N =116 | SCZ<br>N= 61 | HC<br>N=62 | SCZ<br>N =51 | HC<br>N =43 | SCZ<br>N= 14 | HC<br>N =17 |
| **N. of recording** | N = 900 | N = 989 | N=612 | N= 609 | N = 401 | N = 340 | N= 121 | N= 144 |
| **Age** | 26.5<br>(8.82) | 26.4<br>(8.96) | 31.7<br>(9.92) | 33.2<br>(8.79) | 27.2<br>(7.25) | 29.7<br>(8.72) | 28,6<br>(6.71) | 41.9<br>(13.7) |
| **Education** | 12.9<br>(2.74) | 14.9<br>(2.61) | 12.1<br>(1.48) | 12.3<br>(1.11) | 12.7<br>(2.69) | 14.1<br>(2.37) | 11.9<br>(1.37) | 14.7<br>(2.93) |
| **Sex (n. of females and %)** | 45<br>(43%) | 50<br>(43%) | 22<br>(36%) | 24<br>(38%) | 23<br>(45%) | 19<br>(44%) | 4<br>(29%) | 11<br>(65%) |
| **Verbal IQ** | 89.13<br>(18.7) | 102.12<br>(16.1) | 112.0<br>(15.6) | 116.5<br>(16.9) | 96.3<br>(16.6) | 100.27<br>(14.7) | NA | NA |
| **SANS total** | 9.66<br>(4.41) | NA | NA | NA | 7.61<br>(2.99) | NA | NA | NA |
| **SAPS total** | 10.32<br>(4.91) | NA | NA | NA | 7.23<br>(4.79) | NA | NA | NA |
| **PANSS total** | NA | NA | 54.65<br>(10.63) | NA | 75.79<br>(10.44) | NA | 58.86<br>(15.25) | NA |
| **PANSS negative** | NA | NA | 14.61<br>(4.19) | NA | 20.09<br>(5.24) | NA | 15.28<br>(3.95) | NA |
| **PANSS positive** | NA | NA | 11.31<br>(3.13) | NA | 18.51<br>(4.37) | NA | 12.86<br>(4.10) | NA |
| **Illness duration (months)** | 8.87<br>(6.68) | NA | 54.38<br>(83.24) | NA | 63.13<br>(68.87) | NA | 83.86<br>(91.33) | NA |
| **PSP** | 56.01<br>(15.70) | 86.46<br>(6.72) | NA | NA | 54.09<br>(8.02) | NA | NA | NA |

**Note:** The table displays means and standard deviations of demographic (age, education and sex) and clinical information. Clinical symptoms were measured using the Scale for the Assessment of Negative Symptoms (SANS)<sup>37</sup>, the Scale for the Assessment of Positive Symptoms (SAPS)<sup>38</sup>, and the Positive and Negative Syndrome Scale (PANSS)<sup>39</sup>. Social functioning was measured using the Personal and Social Performance scale (PSP)<sup>40</sup>. NA = data not available; SCZ = patients with schizophrenia, HC = healthy controls.

### Voice recordings

The dataset included a total of 2.034 recordings of individuals with schizophrenia (mean recording length = 17.3 seconds (sec), sd = 15.6sec) and 2.082 recordings of control participants (recording length = 18.5sec, sd = 12.7sec). Voice recordings were collected using the Animated Triangles Task (ATT)^41,42^. The task is generally used to measure theory of mind (ToM) and involves twelve video clips representing an interaction between animated geometrical shapes (triangles). The participants were asked to provide an interpretation of what was going on in each animation and their answers were audio-recorded. A detailed description of the task and its validity for assessing speech production is included in **SM2**. Recording setting and experimental procedure are described in detail in **SM3**, and were kept constant across the different sites. All recordings were manually pre-processed to remove background noise and interviewer speech when present, and to ensure that all recordings analyzed had adequate audio quality. A full description of the process and of the extracted acoustic features is available in **SM3**.

### Statistical Modeling

#### Analysis of effect of diagnosis on acoustic features

To estimate the differences between individuals with schizophrenia and HC in the different acoustic measures, we used Bayesian multilevel Gaussian regression models on the current data with each acoustic feature as outcome, and diagnosis (schizophrenia vs. HC) and language (DK, GE, CH, JP) as predictors. Within the same model, we separately assessed the effect of diagnosis for each language, and modeled varying effects of participants, i.e., intercepts and slopes, separately for each group and language. For each acoustic feature, we built a first model with weakly informative priors, i.e., expectations of no effects of diagnosis, thus conservatively regularizing the model parameters, reducing overfitting and leading to improved predictions^43^. We then built a second model with an informed prior (when available), that is meta-analytic effect size (ES), and compared results across the two models. We aimed to assess whether the effects of diagnosis are robust across changes of priors, and whether the skeptical and informed priors led to more robust inference, that is, in lower estimated out-of-sample error - measured in terms of Leave-One-Out based stacking weights^44^. To evaluate the potential role of biological sex (male vs. female), age and level of intelligence, we built additional models, one per each moderator interacting with group separately in the four languages. We then reported the model estimates for the interaction, including credible (i.e., Bayesian confidence) intervals (CIs) and evidence ratios (ERs), i.e., evidence in favor of the effect observed against alternative hypotheses (see **SM4**). When ER was weak (below 10, that is, less than ten times as much evidence for the effect as for alternative hypotheses), we also calculated the ER in favor of the null hypothesis. Further details are presented in the **SM4**. In addition to the more traditional acoustic features included in the previous meta-analysis, we extracted 24 novel voice quality acoustic features including both spectral and glottal properties of voice^28^, for a total of 32 including those of rhythm. Median and interquartile range (IQR) were calculated for each of these measures (see **SM3**). We then estimated the differences between individuals with schizophrenia and HC for all measures (see **SM5**). We report additional analyses in **SM7** to assess the robustness of the findings: we repeat all analyses on audio segments of 6 sec. to control for recording length. The results generally support our main findings and we report here only qualitative divergences.

#### The relationship between acoustic features and clinical features, pharmacotherapy and social functioning

To assess the relationship between acoustic features and clinical ratings, we built Bayesian multilevel regression models with each acoustic feature as outcome, and clinical features (one at a time) as ordinal predictors. We separately assessed the relationship between the different acoustic features and clinical ratings for each language, and modeled varying effects of participants, i.e., intercepts and slopes, separately for each language. This analysis was performed on the schizophrenia group only (see **SM4** for more details).

To assess the effect of medication, patients were divided into two categories based on the mechanism of action of antipsychotic medication^45^, namely patients with (1) low D2R occupancy, i.e., Clozapine, Olanzapine, Paliperidone, Quetiapine or (2) high D2R occupancy, i.e., Aripiprazole, Amisulpride, Risperidone, Ziprasidone, Sertindole (see SM6). Antipsychotic dose was converted to chlorpromazine (CPZ) equivalents^46^. We used Bayesian multilevel regression models, as described above, with each acoustic feature as outcome, and antipsychotic medication (high D2R, low D2R) and language (DK, CH, GE) as predictors. For each acoustic feature we built a first model with weakly informative priors, that is expectations of no effects of medication, and we then built a second model with informed prior (relying on DeBoer et al. 2020^33^, see **SM6**). Our main aim was to assess the generalizability of previous findings on the effect of medication on speech production in schizophrenia, more than evaluating the causal pathways of medication on speech production, which would have required a larger sample and more detailed information on potential confounders (see discussion section). To assess the role of drug dosage, duration of illness (DUI) and social functioning (PSP scale), we built Bayesian multilevel regression models with each acoustic feature as outcome, and chlorpromazine equivalents, DUI and PSP scale score (one at a time) as predictors. We also compared acoustic patterns between patients with first-episode (FES) schizophrenia and chronic patients. We used Bayesian multilevel regression models with each acoustic feature as outcome, illness onset (FES patients, chronic patients and healthy controls) and language as predictors (see **SM6** for more details).

The code used for the analysis and the extracted features are openly available (see **SM8**).

## Results

### Effect of Diagnosis

The detailed results and comparisons to meta-analytic effects are reported in **Table 2** and **Figure 1**. We only partially replicated previous meta-analytic findings: only reduced pitch variability was found across all datasets, and with smaller effect sizes. Duration atypicalities (reduced speech rate, increased pause duration) were replicated only in the German and Danish corpora, and with smaller effect sizes. We also identified a new potential marker across languages: longer utterance duration. In agreement with the inconsistent replications, meta-analytically informed models were more robust and generalizable to new data (LOO weights for skeptic models above .75) only in about half of the models, indicating that meta-analytic findings were not fully representative of the current samples.

**Table 2.**
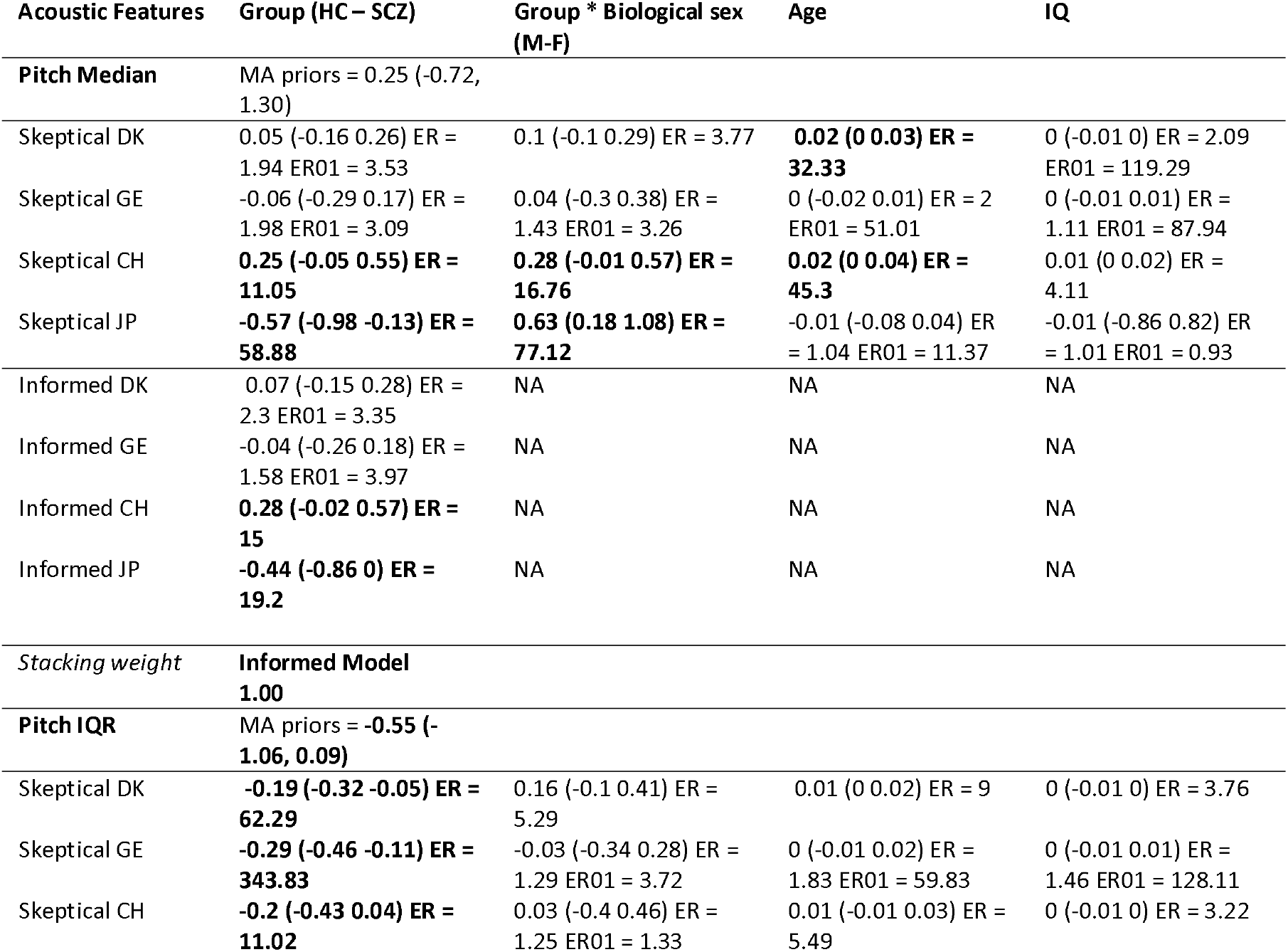
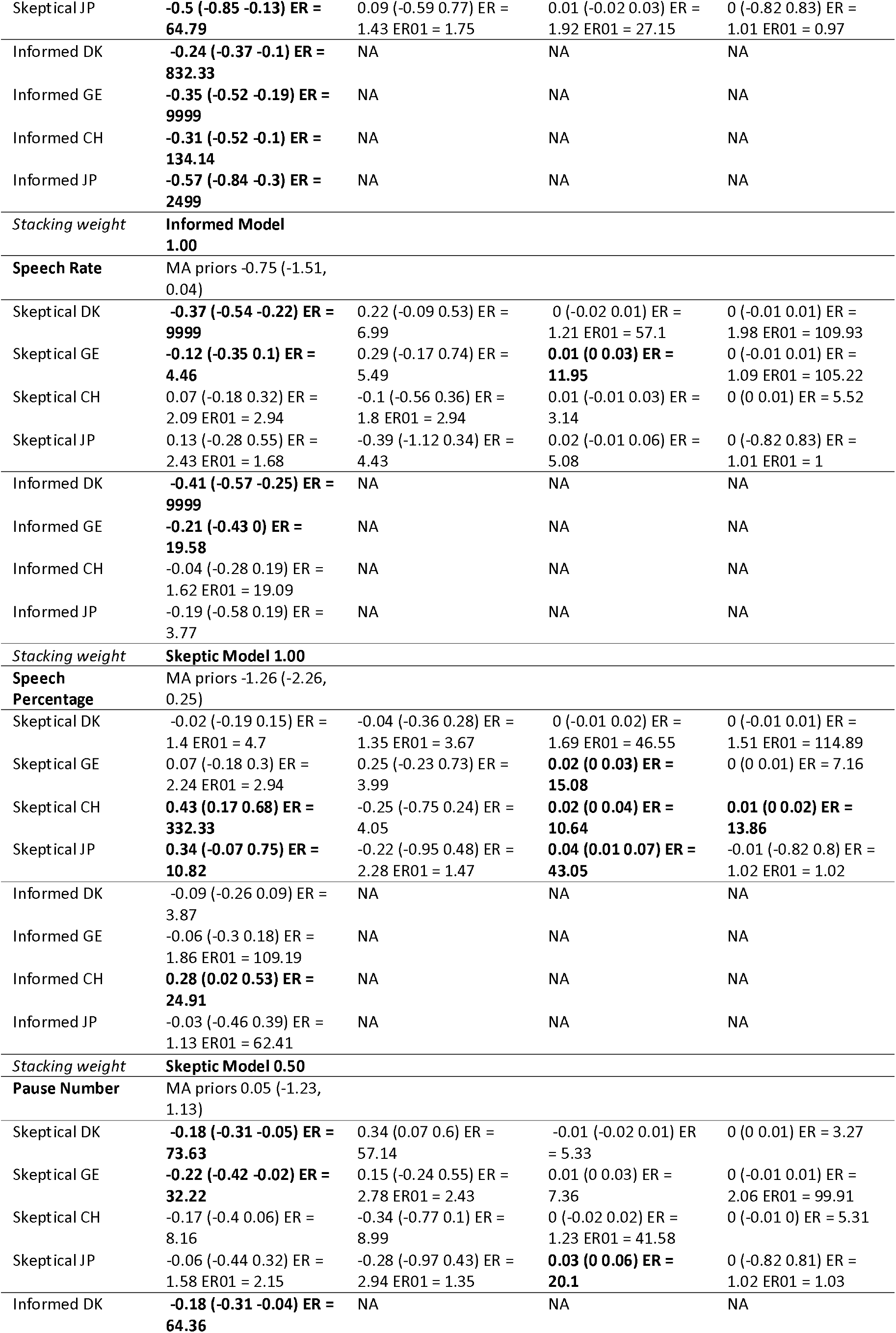
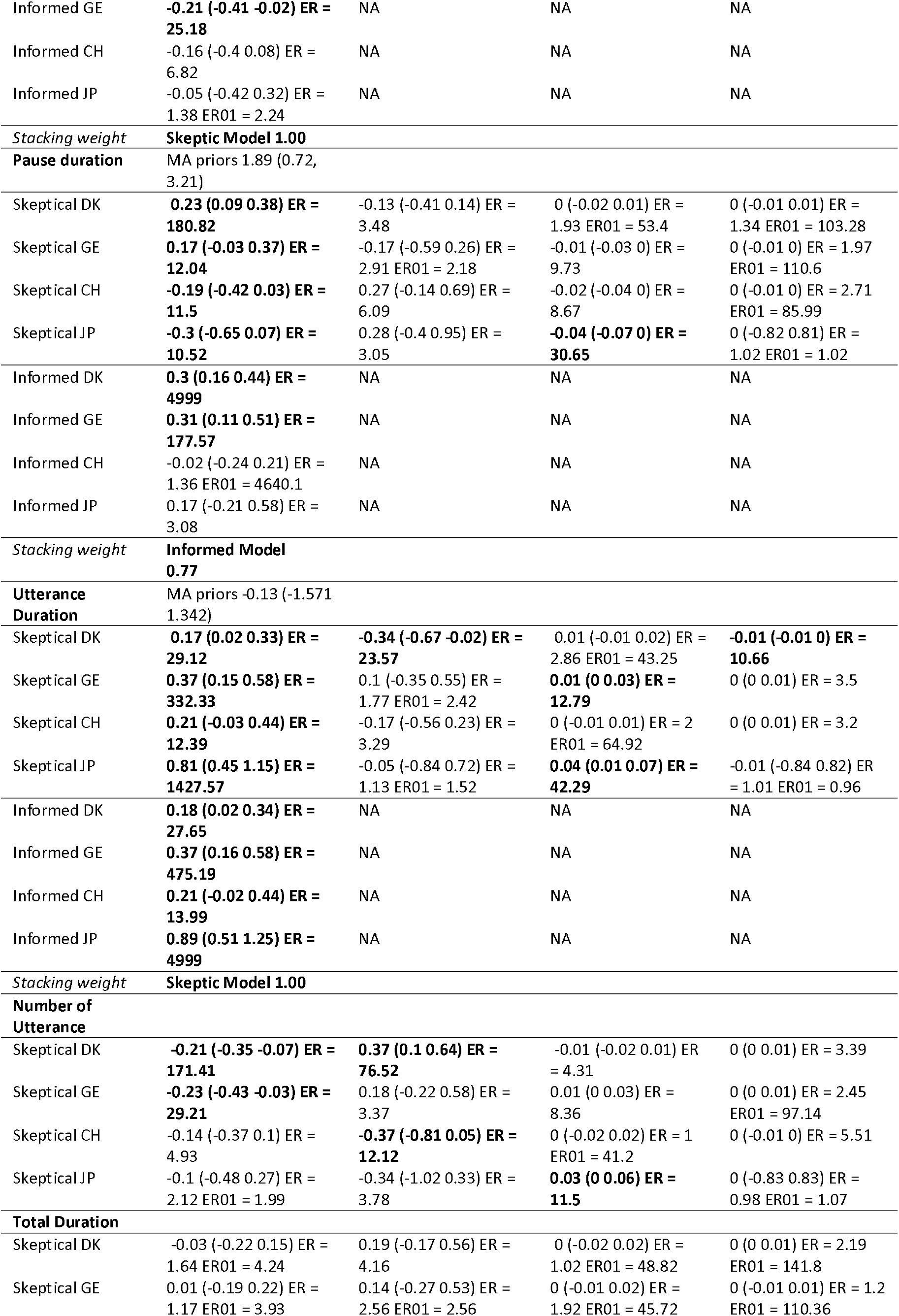
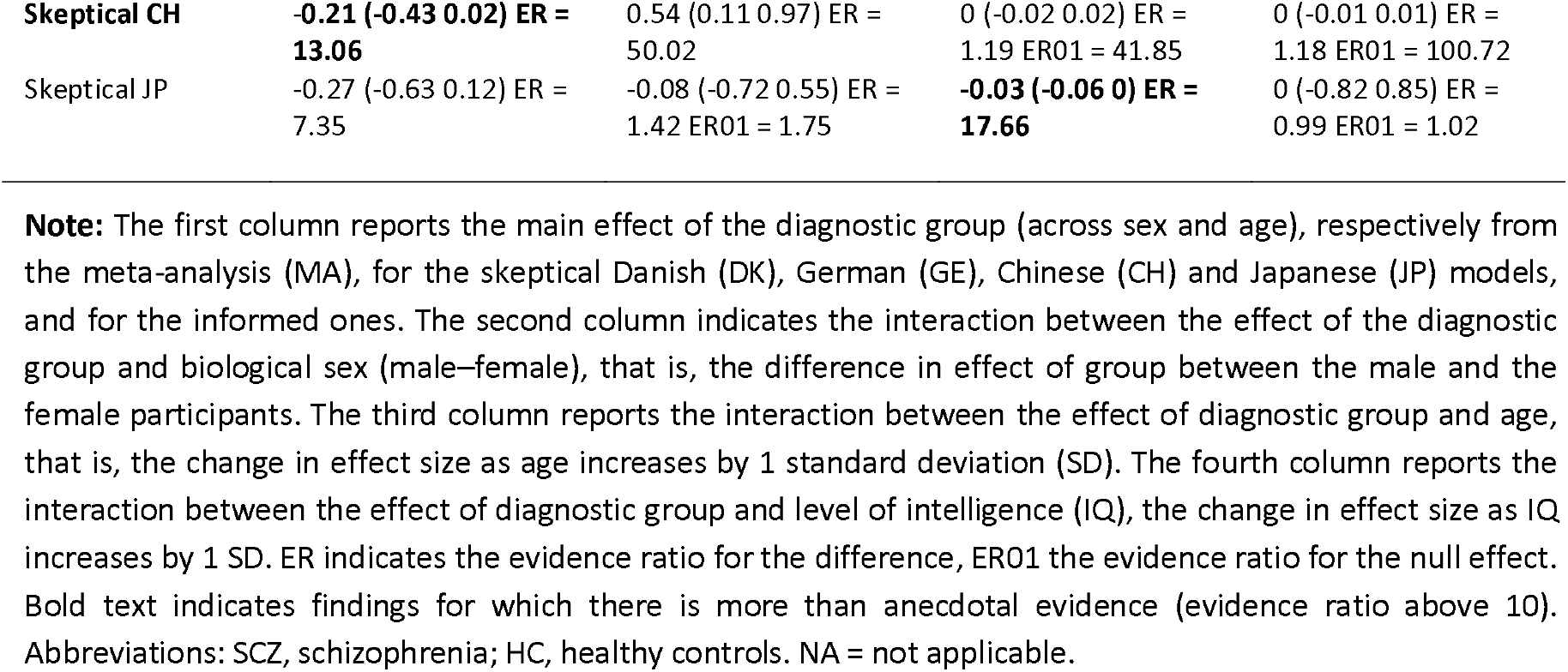
Estimated standardized mean difference (HC-schizophrenia) for the eight acoustic measures present in the meta-analysis, as estimated separately by the meta-analytically informed and the skeptical models

**Figure 1.**
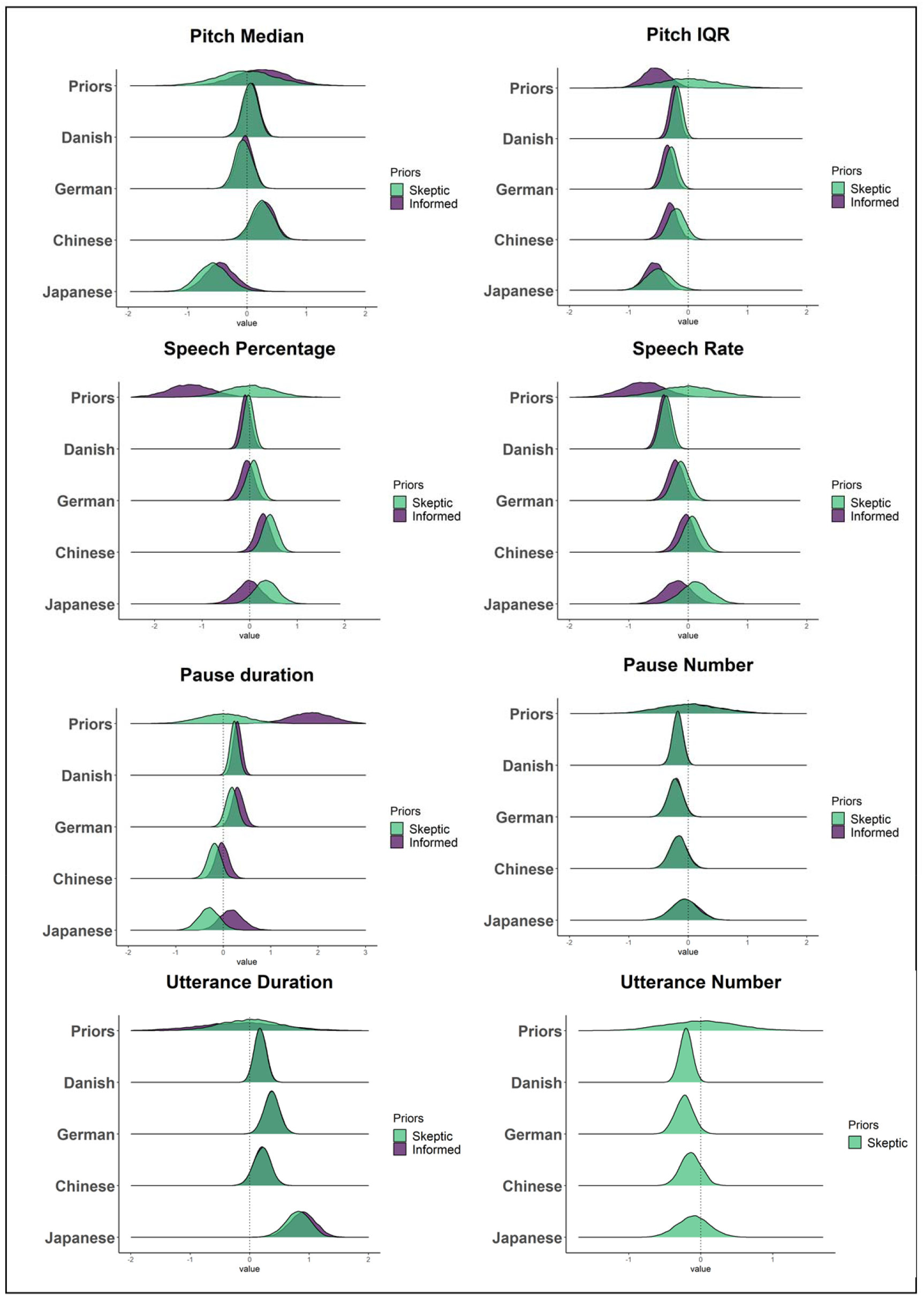
Comparing meta-analysis, skeptical expectations and results. Each panel presents a separate acoustic measure, with the x-axis corresponding to standardized mean differences (schizophrenia - HC) equivalent to Hedges’ g, with estimates above 0 indicating higher scores for patients with schizophrenia. The overlap between the skeptical and meta-analytic posterior distributions suggests no real advantage in using meta-analytic informed priors compared to skeptic ones.

Biological sex, age and level of intelligence of the participants also affected the group differences, although inconsistently across languages.

Our results are generally confirmed when only the 6-second segments of the audio recordings are analyzed, although with some differences: the results of the robustness analysis (see **SM7**) tend to be more consistent with the results of the meta-analysis^3^ (e.g., reduced speech rate also in German, reduced speech percentage in Danish, no evidence of reduced pause duration in Chinese and Japanese). Interestingly, the robustness analysis also showed that patients with schizophrenia were more likely to give very short responses to the task than controls (total recording duration < 6sec, see **SM7**).

### Novel Acoustic features

The detailed results are reported in **Table S5_A**. Generally, we found some evidence for reduced formants median frequency and formants variability, and for increased median relative amplitude (H1H2) and reduced H1H2 variability. However, these findings are small and not robust across languages. Results for the other novel features are very uncertain or inconsistent across languages. As for more traditional features, we found that sex, age, and level of intelligence again affect several differences.

### Effect of Symptoms

Detailed results and comparison with meta-analytic findings are reported in **Table 3**. Clinical features generally correlated with acoustic features, and these associations were in line with meta-analytic priors. However, we did not find reliable and robust associations across all languages. The associations are generally stronger for temporal-duration measures (lower speech percentage, increased pause duration and reduced speech rate are associated with higher flat affect, alogia and negative symptom severity), whereas pitch measures (median and IQR) are more weakly associated with clinical ratings. Most of the correlations are between small and moderate (5-16% of explained variance), and vary across languages and rating scales.

**Table 3.**
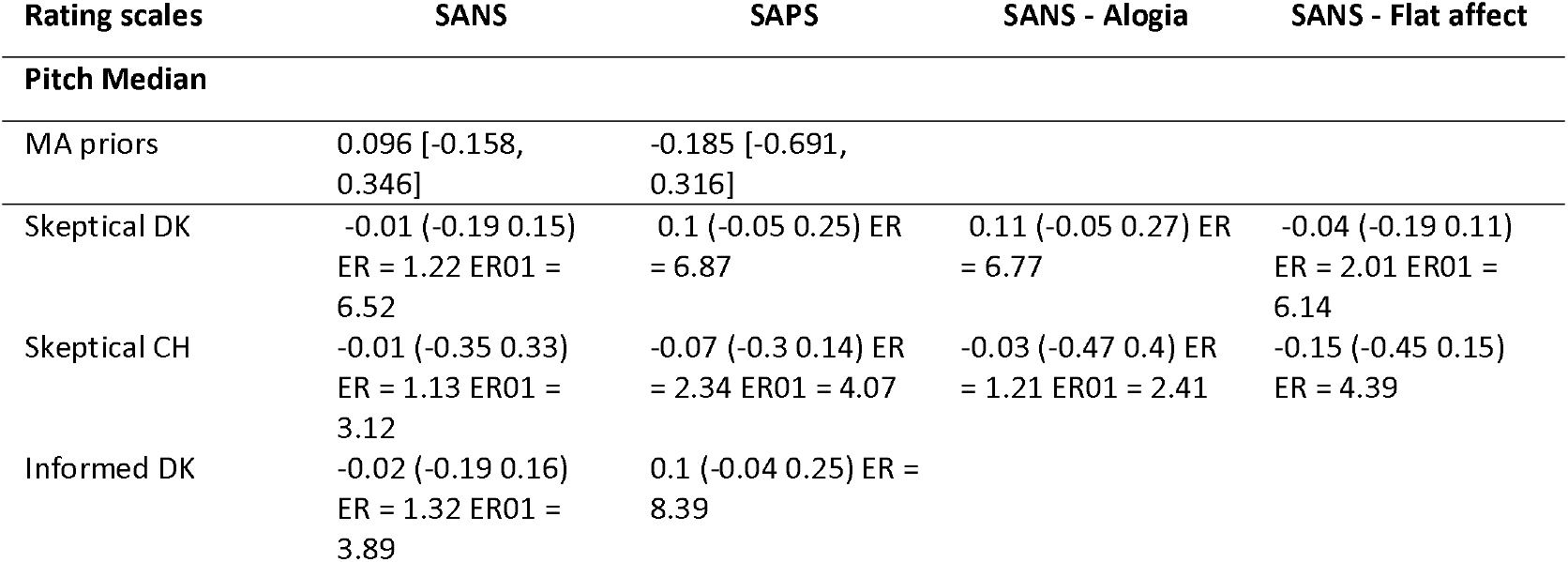
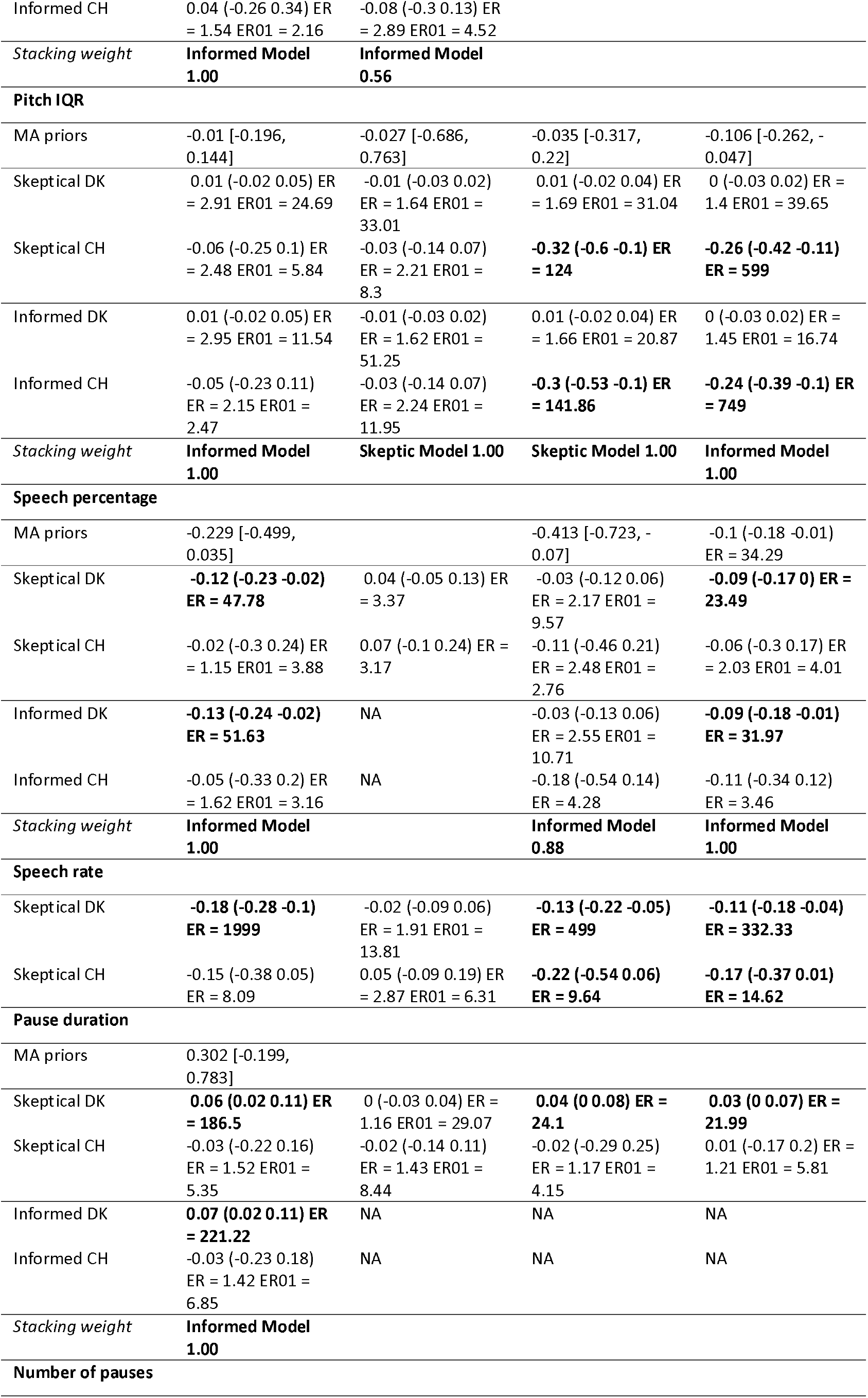
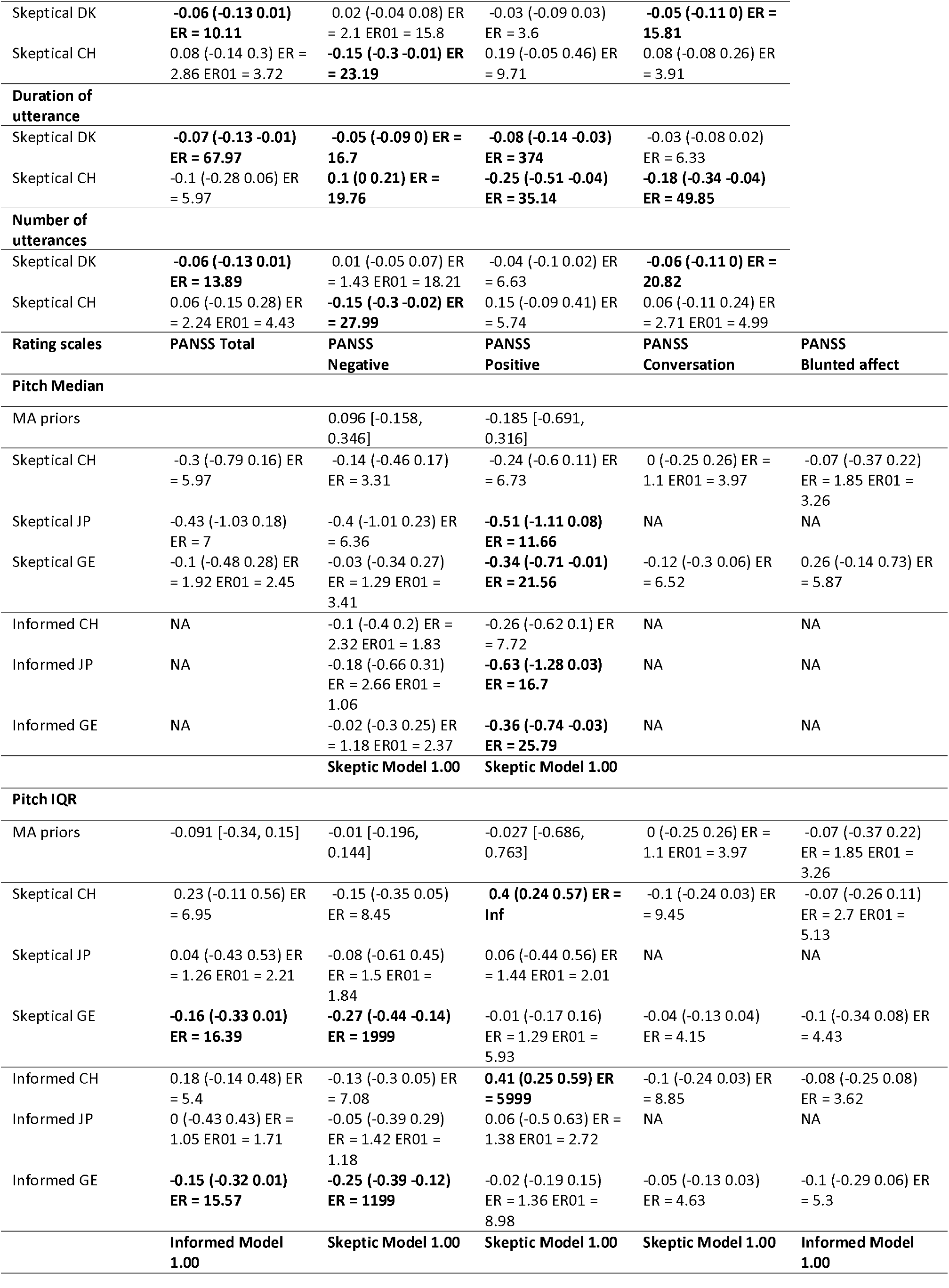
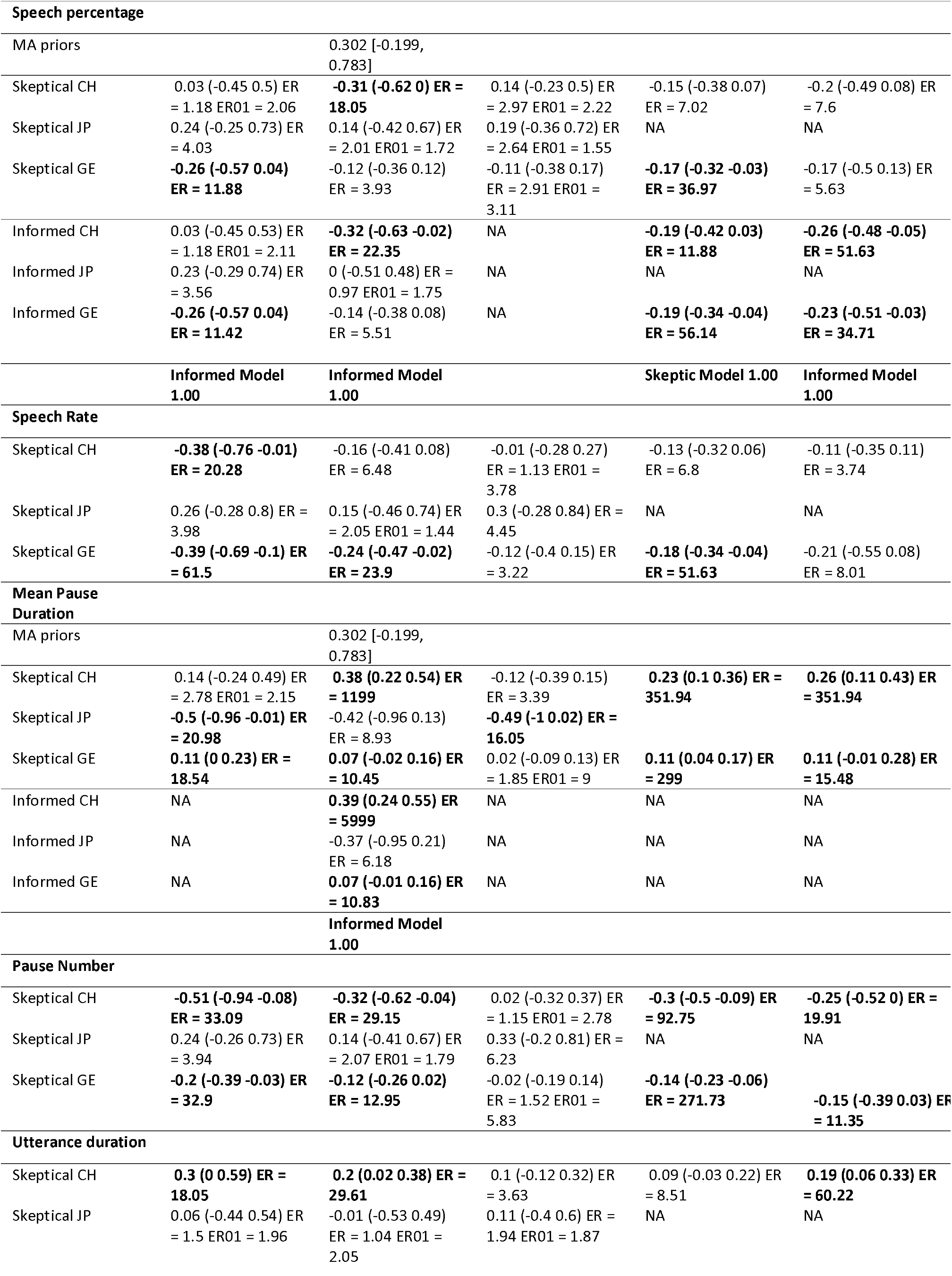
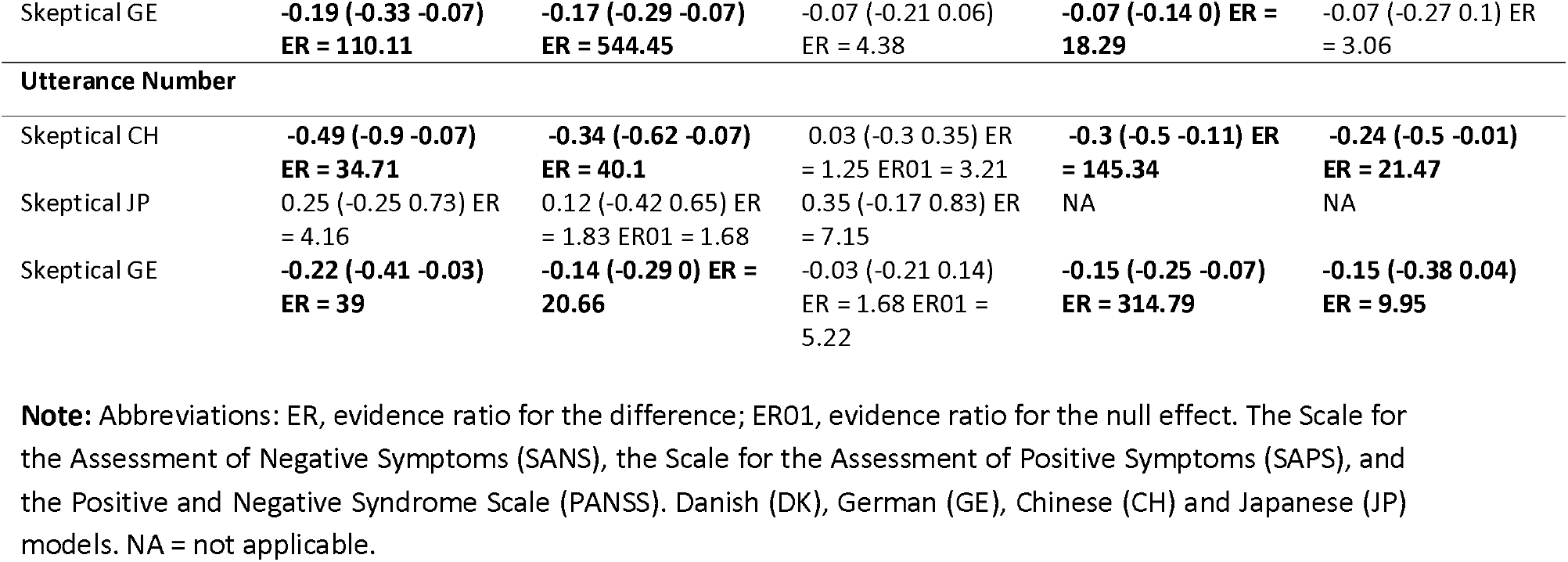
Estimated standardized relation between acoustic and clinical features. ER indicates the evidence ratio for the difference, ER01 the evidence ratio for the null effect.

### Medication, duration of illness and social functioning (PSP)

We found that medication was related to vocal patterns, but inconsistently across languages. The more widespread patterns were that patients who use high D2R occupancy drugs, compared to patients who use low D2R occupancy drugs, show reduced pitch variability, higher number of pauses, longer utterance duration and higher speech rate. Further, higher drug dosage (CPZ equivalents) was associated with lower pitch median, lower speech rate, increased pause and utterance duration and reduced total number of words (see **Table S6_B**). We generally found no difference between patients with FES and patients with chronic schizophrenia, and only weak evidence supporting a role of illness duration: acoustic atypicalities are present already at disease onset and not just associated with a longer and more severe course of the disorder (see **Table S6_C**). Finally, we found that increased speech percentage and reduced pause duration were associated with higher scores on PSP, although only in the Danish corpus (see **Table S6_D**).

## Discussion

In the present paper, we aimed at developing a critical, cumulative scientific approach to the understanding of vocal and prosodic atypicalities in schizophrenia. Relying on a previous meta-analysis^3^ of the field, we systematically assessed the generalizability of established and novel acoustic markers on a new large cross-linguistic dataset. We also assessed whether explicitly incorporating previous findings as informed priors would increase the generalizability of the results and provide additional insights on heterogeneity of the findings compared to previous literature.

### Is there a universal generalizable acoustic profile of schizophrenia?

Our study assessed the generalizability of findings across a heterogeneous dataset (4 different languages). In other words, we assessed whether previous findings would be shown in a new study applying analogous experimental and/or statistical procedures, i.e., replication^47^, “to populations with for instance a different language, age distribution, or other demographic and clinical characteristics”, i.e., generalization^48^. We only found a minimal generalizable acoustic profile of schizophrenia: reduced pitch variability and increased utterance duration, albeit with modest effect sizes. Given the heterogeneity of previous studies and uncertainty about publication bias reported in a previous meta-analysis^3^, even these minimal cross-linguistically generalized findings are far from trivial.

One possible mechanism for the generalizable acoustic profile is a relation to negative symptoms, emotional and effort-related ones. Reduced pitch variability is related to monotone speech and flat affect^49–51^, and increased utterance duration to lower energy and increased vocal effort^52,53^. Further, the most promising of the novel features we investigated (e.g., reduced formant frequency and increased H1H2, albeit not consistent across all languages) also fit this explanation^54–56^. Reduced formant frequencies have been found to be associated with clinical ratings of blunted affect and alogia in schizophrenia^55^, and a decrease in articulatory effort in patients with depression^28^. Furthermore, many studies in patients with depression have shown that an increase in H1H2 can be considered one of the acoustic indicators of breathiness and associated with psychomotor retardation^57^. However, we should be skeptical of any simplistic explanation as yet, since we did not find a cross-linguistically robust association between these acoustic features and clinical ratings of negative symptoms. In particular, we argue that heterogeneity of the studies and samples has not been insufficiently accounted for, so far.

### Source of heterogeneity in the voice profiles of patients with schizophrenia

The second crucial contribution of this study is highlighting the importance and complexity of clinical, socio-demographic, contextual (e.g., speech task) and linguistic differences in assessing vocal markers of schizophrenia.

### Clinical heterogeneity

Individuals with schizophrenia present wildly heterogeneous constellations of clinical features: symptoms, onset and duration of the disorder, as well as medications^24,27^. Moreover, some of the clinical ratings are based on the perceptual assessment of speech features, and accordingly, we should expect acoustic heterogeneity co-varying with clinical heterogeneity^36,54,55^. Indeed, we found associations between acoustic parameters and clinical ratings, with duration aspects being more closely related to clinical ratings, in line with meta-analytic findings^3^. Lower proportion of speech and reduced speech rate, as well as longer pause duration were generally associated with higher ratings of negative symptoms and, in particular, alogia and flat affect.

However, the acoustic atypicalities were generally smaller than differences in clinical ratings^33,43^, and their relation to clinical ratings was often inconsistent across languages and rating scales. This might have several explanations. The first is that the acoustic features analyzed are only a subset of those actually used by clinicians to produce clinical ratings of alogia and blunted affect^15^. Different approaches using larger sets of acoustic features and machine learning techniques could be required to better characterize the acoustic markers of clinical ratings^54–56^. Second, the divergence between acoustic features and clinical ratings can be explained by the fact that these two signals have different temporal (i.e., precision with respect to time) and spatial (i.e., precision with respect to environmental changes) resolution, thus expecting high convergence between the two may not be realistic^49^. Third, different clinical scales might not be fully overlapping in the symptoms they include and in their definition of the symptoms^58,59^, and linguistic and cultural differences may also affect the frequency, expression and rating of symptoms, thus generating several inconsistencies^60,61^.

Different clinical profiles also often imply different medication profiles, and different medications can differentially impact vocal production^41^. Indeed, D2R drug occupancy and medication dosage were shown to relate to acoustic patterns, albeit inconsistently across languages. This suggests that the field needs a more fine-grained assessment of the different medications involved in schizophrenia, its comorbidities and their impact on voice.

Finally, we found no differences between patients with FES and chronic patients, and a limited role of illness duration on acoustic profiles: vocal atypicalities are already present at the onset of the disease and not only associated with a longer and more severe course of the disorder, thus having the potential for tracking its development and monitoring the symptomatology over time.

### Socio-demographic heterogeneity

Another crucial source of heterogeneity in acoustic patterns is socio-demographic heterogeneity. Indeed, we found several reliable effects of sex and age on vocal atypicalities. However, the picture is currently very sparse, and more than identifying systematic effects, we rather recommend socio-demographic variables be taken explicitly into account. Promising progress has been made in normative modeling^62,63^, which relies on large samples to provide expectations, accounting for clinical and socio-demographic features, and assess individual deviations from such expectations. This approach is particularly relevant in light of recent evidence showing how computational speech analysis may be prone to serious bias determined by socio-demographic factors, such as racial identity^64^, as it may help to identify such potential bias. An alternative approach is to collect larger samples and use propensity scores^65,66^ to better match patients and controls and account for potential confounders. These approaches may also help to tackle the problem of selection bias, as most previous studies have used convenience samples with relatively homogeneous clinical and sociodemographic features (e.g., chronic or FES patients), that may not be fully representative of the schizophrenia spectrum and its heterogeneity.

### Linguistic heterogeneity

Not least, albeit previously neglected, linguistic and cultural differences can also play a role. We modeled each language separately to account for the heterogeneity in the different corpora, thus allowing us to compare results across languages. In general, we found that atypical voice patterns were more similar within Germanic (Danish and German) and non-Germanic (Japanese and Chinese) language families. The most prominent results of the previous meta-analysis^3^ (reduced speech percentage and speech rate, longer pause duration) were not consistently found across all languages in our study. One possible source of this discrepancy is the amount of noise and bias present in previous studies. Meta-analyses are often afflicted by publication bias, and heterogeneous quality in the estimates being collated, which often results in consistent discrepancies with multi-lab replications^67^. However, a complementary explanation is that quantitative vocal aspects may vary with the languages being studied, and that vocal atypicalities reported to be abnormal in schizophrenia may not be universally expressed in the same way. For instance, prosodic use of speech rate and pauses to create emphasis may differ across languages. Thus, it is possible that decreased prosodic emphasis is a universal feature of schizophrenia, but the acoustic ways in which this can be measured vary across languages. Vocal patterns must also be considered in relation to their cultural and linguistic context^8,68^, and the generalizability across social and linguistic groups should be systematically tested in future studies.

### Acoustic heterogeneity and speech task

The use of a common standardized task (i.e., the ATT^41^) allowed us to compare voice patterns across the different corpora. This is particularly important considering that a previous meta-analysis^3^ has shown that varying speech tasks may yield different outcomes, and recent studies further support this finding^55,51^. Further, our task included repeated measurements and allowed us to model intra-speaker variability, which previous studies found to be relevant^69–71^. Even by controlling for intra-speaker variability and keeping the task constant, we found large and important differences in voice patterns between the corpora. Further, we noticed that the patients produced very short responses (< 6 sec) much more frequently than controls, and when excluding these trials, we observed that results were more in line with meta-analytic findings^3^. This suggests that acoustic patterns may be task-related (with the task also interacting with language) and not robust across different tasks.

### Limitations and future perspectives

The large differences in terms of the clinical and socio-demographic features within our corpus is a limitation of the present study. For example, the sample size differs across languages, and the samples in the different languages are not exactly matched in terms of the relevant clinical features, such as medication or symptom profiles. Thus, larger sample sizes might be more representative than smaller ones, and partially different subpopulations might be investigated across languages. Further, even if we kept the task and the experimental procedure constant across sites, a more controlled acoustic setting, with high-quality headset microphones placed at a constant distance from the mouth, would have allowed to further reduce potential differences across the different recording settings. This variability, albeit limited, between our corpora in terms of these features, may have contributed to the differences in the main results; however, it also provided an opportunity to assess the generalizability of results across more varied conditions and thus provided more generalizable results. Moreover, although we collected a larger multilingual sample compared to previous studies^3,72^, an ideal sample systematically representative of the schizophrenic spectrum and its clinical and sociodemographic variability would be even larger^63^. Future efforts should thus be directed to build a large open cross-linguistic corpus of speech recordings of patients with schizophrenia able to capture linguistic, cultural, socio-economic and clinical variability. This multilingual corpus may represent an ideal benchmark dataset for testing the reliability and generalizability (e.g., out of sample predictability) of voice analysis results in schizophrenia^73^, and the necessary ground for assessing its clinical applicability^74^. Not least, future studies should focus more on cross-diagnostic comparisons aimed at capturing symptom dimensions which extend over a single disorder^75,76^, and implement longitudinal designs able to test more complex hypothesis on the interaction between antipsychotic medication type and dosage, clinical (e.g., illness severity and duration) and sociodemographic (e.g., sex differences^77^) characteristics, and speech production.

Another limitation is that we focused on single features as markers of schizophrenia, and did not vary the speech task. For example, the specific social context in which the speech task takes place, the social actors involved in it, and the communicative goal to be fulfilled can influence speech production and thus acoustic patterns. Speaking to a superior vs a peer, having a formal interview vs. an informal chat, all involve different prosodic patterns. Individual participants might perceive the experiment context differently from each other, some as more formal than others; and this is further complicated by cultural factors affecting how interactions with researchers are perceived and dealt with, and therefore the acoustic patterns. Further, linguistic and vocal patterns are inherently multidimensional, with different acoustic features interacting with each other, and cross-linguistic variations potentially affecting these interactions. Looking at single features could thus be reductive: future studies should focus more on examining patterns of shared variance across features^78-80^, their relations with different speech tasks (in terms of social and cognitive demands)^78,79^, and with linguistic variability^8,80^. However, this would require testing more fine-grained hypotheses on mechanisms relying on formal linguistic theory^68^ and on psychopathological functioning theory.

## Conclusion

Overall, we found scarce evidence for a universal, distinctive vocal pattern that characterizes schizophrenia: vocal patterns are highly heterogeneous with different sources of heterogeneity interacting with each other at different levels. These results raise some questions on generalizability of previous findings and on the possibility to cumulatively build on them^81^. However, they also indicate where future attempts may be directed: a larger shared multilingual corpus representative of the heterogeneity of the schizophrenic spectrum, a more explicit focus on multidimensional acoustic patterns and their relations with speech tasks, and a self-correcting cumulative approach. These are the necessary conditions to develop effective clinical applications in the near future that can target different ranges of patients and address the issue of potential bias.

## Data Availability

The original speech recordings cannot be shared as they are considered identifiable data, in line with our consent forms and current data privacy regulations.

https://osf.io/h4wj6/

## Acknowledgments

A.P is supported by a Marie Skłodowska-Curie Actions – H2020-MSCA-IF-2018 grant (ID: 832518, Project: MOVES). A.S is supported by the Carlsberg Foundation. K. K has been supported by the Japan Society for the Promotion of Science (JSPS) (PE 07550). The project has been supported by seed funding from the Interacting Minds Center, Aarhus University.

