## Supplementary Material for "Voice patterns as markers of schizophrenia: building a cumulative generalizable approach via a cross-linguistic and meta-analysis based investigation"

The supplementary materials contain the following sections:

- SM1: Description of the voice recordings corpus

- SM2: Animated Triangles Task (ATT)

- SM3: Audio preprocessing and feature extraction

- SM4: Meta-analytic results, model priors, model fitting

- SM5: Novel Acoustic features

- SM6: Analysis on medication, duration of illness and Personal and Social Performance Scale (PSP), and relations with symptoms

- SM7: Analyses on segments of 6 seconds

- SM8: Software implementation notes

- SM9: A cumulative yet self-correcting approach

**S1 – Description of the voice recordings corpus**

**Danish corpus**

The patient and control samples for the present study were collected within three consecutive studies recruiting patients at the same clinical location, i.e., at OPUS - Clinic for people with schizophrenia (SCZ), Aarhus University Hospital Risskov, in the period from 2009 to 2018^1–3^.

The patients had been diagnosed by experienced psychiatrists according to ICD-10 criteria. Exclusion criteria were a history of neurological disorder, severe head trauma, or substance abuse problem according to ICD-10. Patients were excluded if they did not understand spoken Danish sufficiently to understand the testing procedures or if they had an estimated premorbid IQ below 70 based on their history.

All patients with schizophrenia, except the sample reported in Veddum et al. (2019)^3^, were interviewed with the Scale for Assessment of Negative Symptoms (SANS), the Scale for Assessment of Positive Symptoms (SAPS), and the Personal Social Performance Scale (PSP), a measure of social functioning in schizophrenia).

A summary of the medication taken by the patients is reported in **Table S6_A**.

**Control groups**

Patients and non-clinical controls were matched on age, sex, and level of education (except sample 3, matched only on age and sex). Control participants did not have a history of neurological or mental illness (and neither had their first-degree relatives), severe head injury or drug- or alcohol dependence according to ICD-10 criteria. Demographics and social functioning of the patients and their matched controls are summarized in **Table 1.**

**IQ**

Verbal intelligence was estimated from two subtests (Vocabulary and Similarities) from WAIS-III (Wechsler Adult Intelligence Scale, Third edition^4^). The two subtests were chosen based on their high correlation with the verbal WAIS-III IQ-score.

**Ethics**

The participants received written and oral information about the project, and written informed consent was obtained before inclusion. The study was approved by The Central Denmark Region Committees on Biomedical Research Ethics (Ref: M-2009–0035; Ref: 2007-58-0010) and the Danish Data Protection Agency. The project complied with the Helsinki Declaration of 1975, as revised in 2008.

**Chinese corpus**

The patient and control samples were collected within two consecutive studies recruiting patients at the same clinical location, i.e., Renmin Hospital of Wuhan University^1,5^, and a part of the data (n = 41 participants) has not yet been published).

Patients met the diagnostic criteria for schizophrenia according to ICD-10 and were diagnosed by psychiatrists.

Exclusion criteria were a history of severe head trauma or neurological illness or if they had a substance abuse problem according to the ICD-10. Patients with an estimated premorbid IQ below 70 based on previous history or who were unable to understand spoken Chinese well enough to understand testing instructions were also excluded. All of the Chinese patients were of Han Chinese ethnicity.

Patients were interviewed with the Scale for Assessment of Negative Symptoms (SANS), the Scale for Assessment of Positive Symptoms (SAPS), the Personal Social Performance Scale (PSP).

**Control group**

There were no differences between patients and non-clinical controls on age and sex at group level. The exclusion criteria for the healthy subjects were the same as for patients. In addition, healthy control subjects were excluded if they, or a first-degree relative, met any psychiatric diagnosis according to ICD-10.

**IQ**

Verbal intelligence was estimated from Vocabulary subtests from WAIS-III (Wechsler Adult Intelligence Scale, Third edition)^4^. The subtest was chosen based on its high correlation with the verbal WAIS-III IQ-score^4^.

**Ethics**

All participants received written and spoken information about the project, and written consent was obtained. The project was approved by the Ethics committee of Renmin Hospital of Wuhan University and the Institutional Review Board of the Institute of Psychology, the Chinese Academy of Sciences. The authors assert that all procedures contributing to this work comply with the ethical standards of the relevant national and institutional committees on human experimentation and with the Helsinki Declaration of 1975, as revised in 2008.

**Japanese corpus**

The patient and control samples were collected within the study of Koelkebeck et al. (2013)^6^.

The participants in this study included 18 patients with schizophrenia who had been referred to the Department of Neuropsychiatry of Kyoto University Hospital. This group, which comprised both inpatients and outpatients, was medicated and clinically stabilized, and all of the members met the DSM-IV (APA, 1994) criteria for schizophrenia, as determined by a Structured Clinical Interview for DSM-IV Axis I Disorders (SCID I). Exclusion criteria were a history of any other psychiatric disorder.

**Control group**

The control group was composed of 30 healthy participants matched for age and gender with patients with schizophrenia. Exclusion criteria were the presence of any history of other psychiatric disorders, neurological disorders, and the use of psychotropic medications. Written informed consent was obtained from all participants.

**Ethics**

The design of this study was approved by the local Ethics Committee and conforms with the Declaration of Helsinki.

**German corpus**

The patient and control samples for the present study were collected within three consecutive studies recruiting patients at the same clinical location, i.e., the University of Muenster and the LWL-Hospitals Muenster^7–9^ (LWL, from 2005 to 2017).

The patients had been diagnosed with schizophrenia by experienced psychiatrists using the Structured Clinical Interview for DSM-IV SCID-I. Psychopathology was assessed with the Positive and Negative Syndrome Scale (PANSS).

Patients with any history of other psychiatric disorders, neurological disorders, serious head injury, alcohol or illegal drug abuse, or insufficient knowledge of the German language were excluded from the study.

**IQ**

Verbal intelligence was estimated from Vocabulary subtests from WAIS-III (Wechsler Adult Intelligence Scale, Third edition)^4^ in Pedersen et al., (2009) and Koelkebeck et al. (2010). The subtest was chosen based on its high correlation with the verbal WAIS-III IQ-score. Verbal intelligence was estimated from Multiple Choice Vocabulary Test (Mehrfachwahl-Wortschatz-Intelligenztest; MWT-B)^10^ in Koelkebeck et al., (2017). The MWT-B measures crystallized intelligence on the basis of verbal capacities.

**Control group**

Healthy controls (HC) were matched with patients for age, sex and education. Healthy participants with no history of Axis-I DSM-IV diagnoses (SCID), illegal drug use, alcohol abuse or addiction, or neurological disorders served as a control group. Subjects with any first-degree relatives with a history of mental disorders were excluded from the study.

**Ethics**: After hearing a complete description of the study, written informed consent was obtained from all participants. The studies were approved by the Ethics Committee of the State Chamber of Physicians Westphalia-Lippe and the University of Muenster and has been carried out in accordance with the Declaration of Helsinki.

**Power-analysis**

**We relied** **(admittedly) on the post-hoc power analyses reported in the meta-analysis of Parola et al. (2020). The smallest reliable effect size found in the meta-analysis was a Cohen’s d of 0.6 for pitch variability. Relying on G*power (Faul et al., 2009), and taking those results at face value, a statistical power of 95% would require 74 participants per group with one measure each, and 12 participants per group with 8 repeated measures each (assuming a correlation of 0.3 between repeated measures) as in our study. With the exact sample size and repeated measures available in the current study, we can achieve an a posteriori 95% statistical power for effect sizes (Cohen’s d) of 0.15 for Danish, 0.20 for German, 0.23 for Chinese and 0.42 for Japanese. Our results indicated effect sizes of the differences in pitch variability between patients and controls of: -0.19 (-0.32 -0.05) for the Danish corpus, -0.29 (-0.46 -0.11) for the German corpus, -0.2 (-0.43 0.04) for the Chinese corpus and -0.5 (-0.85 -0.13) for the Japanese corpus (see results section). These data thus confirm that our samples and repeated-measurement design have the required statistical power for detecting the effects of interest.**

**S2 – Animated Triangles Task**

Voice recordings were collected using the Animated Triangles Task^11,12^. The task is generally used to measure theory of mind (ToM) and involves between eight to twelve video clips representing an interaction between animated geometrical shapes (triangles). In some of the clips the two triangles are moving randomly and unintentionally (e.g., bouncing off walls) (4 clips), and in the other clips the triangles are interacting intentionally to influence the mental state of one another (e.g., a larger triangle trying to convince a small one to leave a closure) (4 clips) or merely performing an activity alone or together (4 clips). The duration of each animation is approximately 40 seconds. The participants were asked to provide an interpretation of what was going on in each animation and their answers were audio-recorded. Not all studies used the 4 pure Action clips.

As shown in a recent meta-analysis^13^, free speech tasks and dialogic tasks with social requirements, compared to constrained speech (e.g., reading), had significantly larger effects both in contrasting patients and controls and in assessing symptomatology. The Animated Triangle Task is thus a more suitable task (compared to, e.g. a constrained speech task) for assessing whether voice patterns can be a marker of schizophrenia and comparing them across languages^14^.

**S3 – Voice recordings setting, preprocessing and features extraction**

**Voice recording setting**

The recording setting within each language was kept uniform:

•	Equipment: we used external medium quality audio recorders (e.g., Zoom H2n Handy Recorder for Danish data, or Olympus Digital Voice Recorder DW-90 for German data, link: https://www.amazon.it/Olympus-Digital-Voice-Recorder-DW-90/dp/B00005OBDS) placed near the participant and on the side of the laptop on which the triangle task was presented.

•	Procedure: participants sat in a silent room in front of a laptop on which the Triangle task was presented, and the experimenter sat near them. After providing instructions, the experimenter started the program. After viewing each video on the laptop screen, participants were asked to give an interpretation - addressed to the experimenter -, of what was happening in each animation, and their responses were audio-recorded.

•	Instructions: the administration procedure and the instructions for the Triangle task are standardized (see Abell et al., 2000^11^; Castelli et al., 2000^12^) and were therefore consistent across settings and samples.”

**Preprocessing**

After the audio-recordings were collected, the speech preprocessing procedure was performed: all audio recordings were carefully listened to in order to identify potential audio quality issues that would lead to the exclusion of the recording. All audio recordings were then manually timecoded, to identify the segments corresponding to the participants’ video description, thus excluding instructions, the prompts (e.g., “Can you say anything more about that?”) as well as questions, backchannels such as (‘hmmm’, or ‘ok’, etc.) provided by the examiner. Once the relevant audio segments had been identified, we proceeded to clean the audio. Background room noise, reverberation and hum were removed from the audio recordings using iZotope RX 6 ElementsTM. Long-term average spectra for each recording were inspected for possible noise artefacts and further cleaned if any were found.

This ensured that all recordings analyzed had adequate audio quality. While semi-automatic algorithms may be used for automatic or semi-automatic removal of cross-talk and background noise removal, and have been employed in previous studies (e.g., de Boer et al., 2021^42^), they generally produce higher rates of error, especially with non-English languages and atypical speakers, thus tending to introduce systematic bias. In contrast, manual pre-processing, even if more demanding in terms of time and resources, ensures higher quality data and can be used in future works as a benchmark to assess potential bias in the automated processing.

We then first extracted measures of rhythm and duration for the full relevant segment (e.g., one video description) using the open-source script syllablev2 for Praat^15,16^. Six features were selected in order to build on the previous meta-analysis^13^ (see **Supplementary Material – S3**): *speech rate* (number of words per second), *proportion of spoken time*, *average pause duration* (in seconds), *average number of pauses* (per second), *average sentence duration* (in seconds), *average number of sentences* (per second) (for a more detailed description see **Table S3_A**)

We extracted 13 pitch and voice quality measures every 10 milliseconds using Covarep for Matlab^17^. The measures included both spectral and glottal properties of voice: fundamental frequency (pitch), formants frequency (F1- F2 -F3 -F4 -F5), relative amplitude (H1H2), quasi-open-quotient (QOQ), normalized amplitude quotient (NAQ), harmonic richness factor (HRF), parabolic spectral parameter (PSP), maxima dispersion quotient (MDQ), harmonic to noise ratio (HNR) (see **Table S3_B** for a description). Median and interquartile range (IQR) were calculated for each of these measures using R^18^. This yielded 26 measures of pitch and voice quality, for a total of 32 including those of rhythm.

The choice of pitch and rhythm measures was motivated by their widespread use in the study of vocal markers of schizophrenia^13^. The choice of voice quality features was motivated by an informal review of the recent speech processing literature (e.g., for an overview, Cummins et al., 2015; Cohen et al., 2020^19,20^). While the distance between recorders and participants was roughly similar across languages and participants, participants could still move through the experiment and the distance between mouth and microphone cannot be assumed to be constant. Since some acoustic features based on intensity are more likely to be affected by the distance, we excluded them from the analyses.

We opted to use median and IQR of acoustic measures, contrary to more commonly used mean, standard deviation and range, because they are more robust to measurement errors (e.g., erroneous jumps of octave in the pitch detection algorithm). Further, fundamental frequency and other acoustic features of the human voice often present long tailed distributions, in which case mean and standard deviation become strongly correlated and determined by the tail. Median and interquartile range more robustly present independent measures of mode and variance of the distribution, respectively.

**Table S3_A**

| **Traditional Acoustic feature** |  | **Description** |
| --- | --- | --- |
| Fundamental frequency – F0 | Pitch median | Pitch mean reflects the mean frequency of vibrations of the vocal cords during vocal production across the linguistic unit analyzed (phoneme, word, sentence, or the entire speech sample). |
|  | Pitch variability (IQR) | Pitch variability indicates the mean magnitude of changes in pitch across the linguistic unit analyzed (phoneme, word, sentence, or the entire speech sample). |
| Speech production | Speech rate | Speech rate is defined as the number of words per time unit (second). |
|  | Percent time talking | Percent time talking represents the percentage of time the speech sample contained a pitch different from zero, i.e. speaking time, relative to the total time of the speech sample. |
|  | Duration of pauses | Mean pause duration indicates the mean duration of pauses across the entire speech sample. |
|  | Number of pauses | Mean number of pauses across the entire speech sample per time unit (second). |
|  | Duration of utterance | Median utterance duration indicates the median duration from beginning to end of the defined speech sample per time unit (second). |
|  | Number of utterances | Mean number of sentences across the entire speech sample per time unit (second). |

**Table S3_B**

| **Novel Acoustic Features** | **Description** | **Reference** |
| --- | --- | --- |
| Formants frequency (F1- F2 -F3 -F4 -F5) | Range of frequencies in which there is absolute or relative maximum in the sound spectrum. The frequency at the maximum is the formant frequency | (Titze et al., 2015; ANSI, 2004^21,22^) |
| Relative amplitude (H1H2) | Amplitude of the first harmonic relative to the second harmonic. Associated with breathiness. | (Hanson, 1995^23^) |
| Harmonics to Noise Ratio (HNR) | Ratio of harmonics to inharmonic (spectral components which are not a whole number multiple of F0) components. Associated with hoarseness. | (Henrich et al., 2001^24^) |
| Normalized Amplitude Quotient (NAQ) | Parametrization of the glottal closing phase using two amplitude-domain measurements from waveforms estimated by inverse filtering. Associated with breathiness. | (Alku et al., 2002^25^) |
| Quasi-Open-Quotient (QOQ) | Relative open phase duration of a glottal cycle. Associated with breathiness. | (Szkiełkowska et al., 2018^26^) |
| Maxima Dispersion Quotient (MDQ) | Sharpness of the glottal excitation. Associated with tense-lax properties of the voice and relatedly with breathiness. | (Gobl et al., 2015^27^) |
| Parabolic Spectral Parameter (PSP) | A quantification of the glottal volume velocity waveform. Associated with breathiness and tense phonation. | (Alku et al., 1997^28^) |
| Harmonic Richness Factor (HRF) | Ratio between the sum of the amplitudes of harmonics, and the amplitude at the fundamental frequency, quantifying the amount of harmonics in the magnitude spectrum of the glottal source. | (Drugman, Bozkurt, et al., 2012^29^) |

**S4 - Meta-analytic results and model priors**

Meta-analytic effect sizes (ES) were calculated using the dataset of the most recent meta-analysis on the topic ^28^ available here: https://www.biorxiv.org/node/923780.external-links.html. For each acoustic feature we extracted the estimate of the standardized mean difference (SMD; also known as Hedges’ g) between individuals with schizophrenia and HC. To estimate associations between vocal patterns and clinical features, we extracted the raw correlation coefficient (Pearson’s r). The effects were then analyzed using 2-level hierarchical Bayesian regression models to estimate the summary effect sizes and corresponding credible (i.e., Bayesian confidence) intervals. We explicitly modeled the heterogeneity (or σ^2^) in the results of the studies by varying effects by article. A detailed description of the statistical procedure adopted is provided in Parola et al. (2020)^13^. This yielded summary effect sizes for: a) the effect of diagnosis for the following features (**Table S4_A**) - b) the association between clinical features and the acoustic features (see **Table S4_B**).

**Table S4_A.** Main Results of the meta-analysis for the effect of diagnosis on acoustic measures.

| **Acoustic Features** | **Number of participants included in the meta-analysis (female)** | **Number of studies (articles)** | **Estimates -Hedges’g**  **[95% CI]** | **P - value** | **ER (Credibility)** | **Sigma squared**  **[95% CI]** |
| --- | --- | --- | --- | --- | --- | --- |
| Pitch Mean | 103 SZ (22 F)  95 CT (23 F) | 4 (4) | 0.253 [-0.724, 1.304] | .273 | 3.467 (78%) | 1.131 [0.005, 7.19] |
| Pitch variability | 387 SZ (92F)  257 CT (106 F) | 11 (8) | -0.546 [-1.059, -0.095] | .005 | 99.0 (99%) | 0.566 [0.152, 1.64] |
| Proportion of spoken time | 267 SZ (106 F)  211 (98 F) | 11 (9) | -1.26 [-2.257, -0.245] | .001 | 149.943 (99%) | 2.538 [0.787, 7.224] |
| Duration of utterance | 93 SZ (30 F)  72 CT (30 F) | 4 (4) | -0.155 [-2.556, 2.26] | .739 | 1.475 (60%) | 6.045 [0.32, 37.49] |
| Speech rate | 336 SZ (111 F)  259 (107 F) | 11 (9) | -0.75 [-1.514, 0.036] | .015 | 32.473 (97%) | 1.447 [0.467, 3.915] |
| Duration of pauses | 221 SZ (128 F)  150 CT (92 F) | 9 (8) | 1.891 [0.721, 3.213] | < .001 | 234.294 (100%) | 3.129 [0.754, 10.086] |
| Number of pauses | 68 SZ (23 F)  40 CT (13 F) | 5 (4) | 0.046 [-1.225, 1.131] | .782 | 1.321 (57%) | 1.531 [0.017, 8.496] |

**Table S4_B.** Main Results of the meta-analysis for the correlations between acoustic measures and clinical symptoms ratings.

| **Acoustic Features** | **Clinical features** | **Number of studies** | **Number of participants included in the meta-analysis** | **Pearson’s r**  **95% CI** | **P -value** | **ER (credibility)** | **Sigma squared**  **95% CI** |
| --- | --- | --- | --- | --- | --- | --- | --- |
| Pitch Mean | Negative symptoms | 7 (3) | 107 SZ (33 F)  21 | 0.096 [-0.158, 0.346] | .136 | 4.198 (81%) | 0.071 [0.00, 0.345] |
|  | Positive symptoms | 4 (3) | 107 (33 F)  21 | -0.185 [-0.691, 0.316] | .04 | 6.89 (87%) | 0.245 [0, 1.714] |
| Pitch variability | General psychopatology | 5 (4) | 146 (48 F)  22 | -0.091 [-0.34, 0.15] | .3 | 3.84 (79%) | 0.057 [0, 0.354] |
|  | Negative symptoms | 11 (6) | 261 (77 F)  22 | -0.01 [-0.196, 0.144] | .836 | 1.117 (53%) | 0.041 [0.002, 0.135] |
|  | Positive symptoms | 4 (3) | 107 (33 F)  21 | -0.027 [-0.686, 0.763] | .698 | 1.509 (60%) | 0.525 [0.001, 4.294] |
|  | Alogia rating | 9 (7) | 313 (68 F)  26 | -0.035 [-0.317, 0.22] | .465 | 1.5 (60%) | 0.135 [0.032, 0.421] |
|  | Flat affect rating | 13 (10) | 403 (81 F)  30 | -0.106 [-0.262, -0.047] | .044 | 11.719 (92%) | 0.053 [0.009, 0.153] |
| Intensity variability | Flat affect rating | 6 (5) | 158 (22 F)  30 | -0.005 [-0.324, 0.308] | .745 | 1.03 (51%) | 0.117 [0.001, 0.658] |
| Proportion of spoken time | General psychopatology | 5(4) | 124 (35 F)  22 | -0.026 [-0.53, 0.375] | .85 | 1.714 (63%) | 0.268 [0.005, 1.411] |
|  | Negative psychopatology | 9 (5) | 146 (35 F)  22 | -0.229 [-0.499, 0.035] | .198 | 23.29 (96%) | 0.131 [0.027, 0.405] |
|  | Alogia rating | 5 (4) | 138 (23 F)  22 | -0.413 [-0.723, -0.07] | < .001 | 58.259 (98%) | 0.127 [0, 0.805] |
|  | Flat affect rating | 6 (5) | 161 (23 F)  22.5 | -0.384 [-0.612, -0.082] | < .001 | 83.211 (99%) | 0.08 [0, 0.456] |
| Duration of pauses | Negative psychopatology | 4 (4) | 109 (30 F)  24 | 0.302 [-0.199, 0.783] | .003 | 15.667 (94%) | 0.246 [0, 1.754] |

We used ES estimates as informed priors in the analysis. In the meta-analysis, we performed a moderator analysis, which revealed an effect of speech production task both in contrasting patients and controls and in assessing symptomatology. However, given the very limited number of studies for each speech task category, i.e., c*onstrained production, free monological production and social interaction,* and considering that for most of the acoustic features the estimates were not available for all the different speech task categories, we here used the global summary effect size (not divided by task category) for each feature. For ease of comparison with the standardized effect sizes used in the meta-analysis, we standardized our acoustic features, that is, we centered them on the mean, and divided them by the standard deviation, separately for each language. Scaling within language has two important motivations. First, it makes our study more comparable with previous monolingual studies. Second, if one language was to present a different variance in a feature than other languages, cross-linguistic effects would be less comparable.

**Model priors**

**Analysis on differences between patients with schizophrenia and healthy controls**

We first specified the prior distributions for each parameter to be estimated. We adopt a weakly skeptical approach to priors, in that we aim at reducing the prior probability of extreme values for the parameters, while not overly influencing the inference.

Skeptical priors- were specified as normal distribution centered at zero (no group difference), with a standard deviation of 0.5. The expected estimates were thus predominantly included within -.1.5 and 1.5, but could still be swayed by the data. Individual variability was modeled using a positive half-normal prior centered at 0, with a standard deviation of 0.7 from the estimate for the specific group and language of the individual, thus regularizing the inference. Sigma was modeled using a normal distribution centered at 0.3 with a standard deviation of 0.1. Informed priors - when available - were normal distributions based on the meta-analytic effects.

This choice of priors did stem from initial (potentially subjective) intuitions as to plausible ranges of difference in acoustic features between patients with schizophrenia and controls, based on previous experiences on similar modeling practices. However, these choices underwent extensive rigorous checks to ensure the priors to be adequate and only weakly informative^30^, that is, that the plausible values of coherence measures predicted by the model on the base of the priors only would not present odd biases. In particular, we first performed prior predictive checks, making predictions as to the data we would predict based on the priors and likelihood function only, before fitting the model to the actual data (see **Figure S4_A**). After fitting the model to data (and before assessing the inferences in the model and testing hypotheses), we performed prior-posterior update checks. In other words, we plotted prior against posterior estimates for each parameter of interest and assessed the relative impact of the prior. This meant making sure that the posterior estimates had lower variance (more peaked and narrower) than the prior ones, and that they were well within the range covered by the prior expectations. In other words, we checked whether the model had learned from the data (posterior estimates being more confident than the priors), and whether the prior had unduly restrained the range of the posterior estimates (posterior estimates pushing against the low probability boundary set by the prior). For varying effects, which have more explicit regularizing functions (defining the amount of partial pooling across, in this case, participants within the same group), the second concern was more pressing than the former. Prior-posterior update checks are reported below (see **Figure S4_B** and **Figure S4_C**). Note that all priors were thus assessed and this motivated some adjustments. The adjustment of the priors is purely motivated by the need to ensure good model quality: an a priori reasonable region of the parameter space to search during the model fitting, the evaluation of potential biases introduced by priors in the inference, etc. The adjustment of priors is not motivated by the results, and it is indeed performed before any hypothesis testing.

**Figure S4_A** –Plot of the prior predictive checks (light blue) and observed data (bold blue). Note that the four language groups (DK, GE, JP, CH) are given the same prior expectations for all parameters, thus implicitly assuming no difference between them (so that any evidence for a difference that we might find would come exclusively from the data and not from the priors).

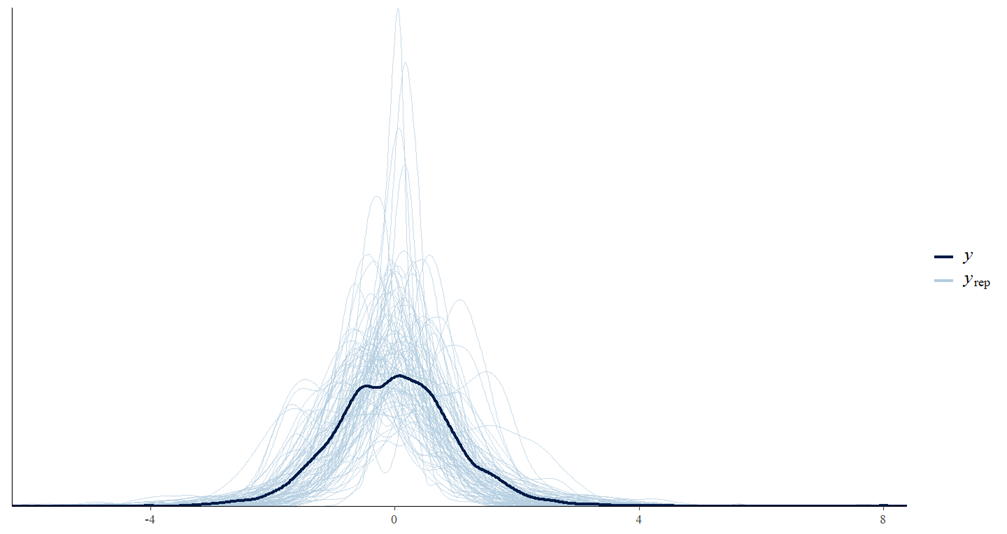

**FIGURE S4_B.** Plot of the prior-posterior predictive check. Prior (light blue) and posterior (dark blue) distributions *for the intercept* in the model *for the Danish language* for speech rate.

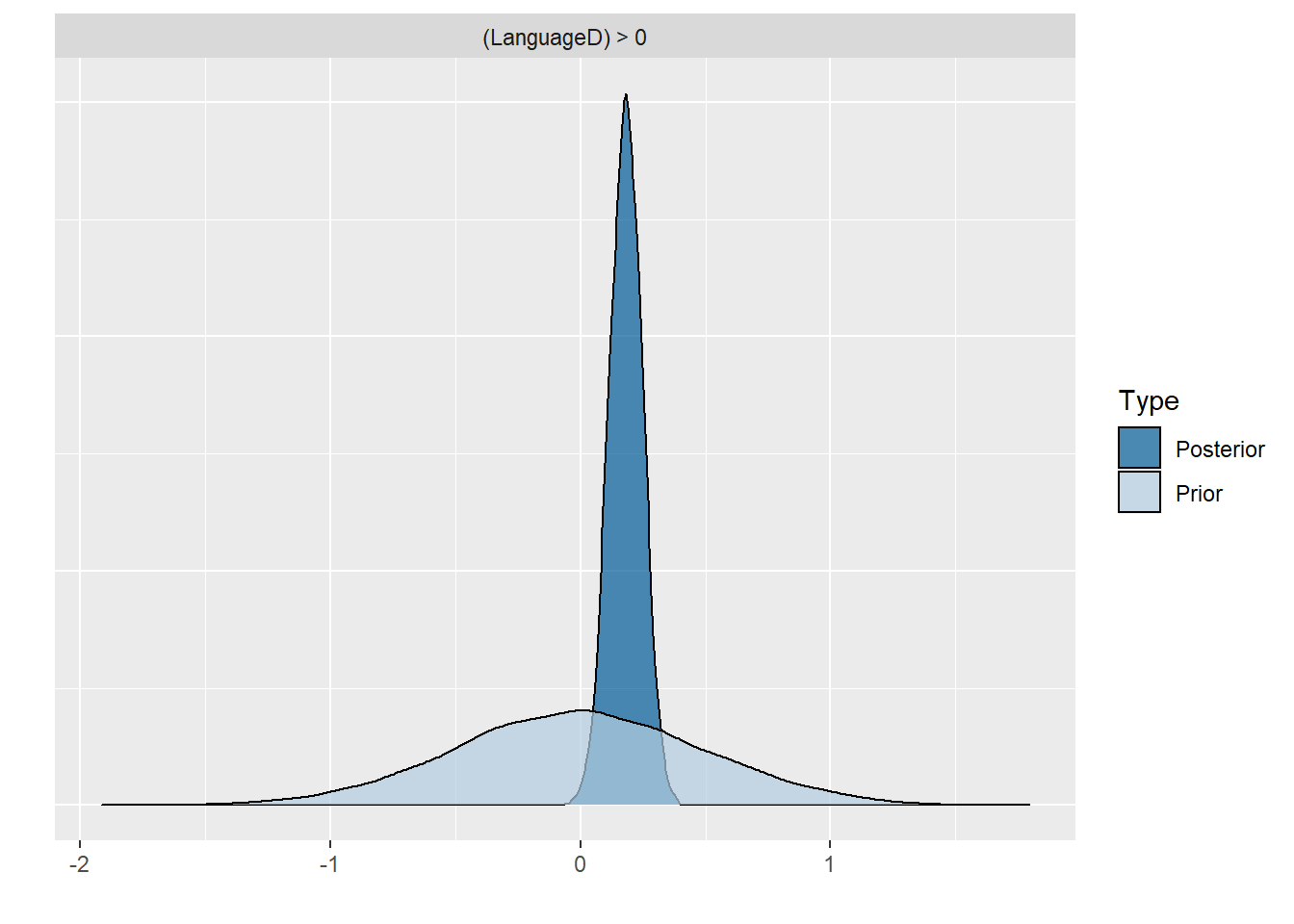

**FIGURE S4_C.** Plot of the prior-posterior predictive check. Prior (light blue) and posterior (dark blue) distributions for the *effect of diagnosis (SCZ-HC) for the Danish language* for speech rate.

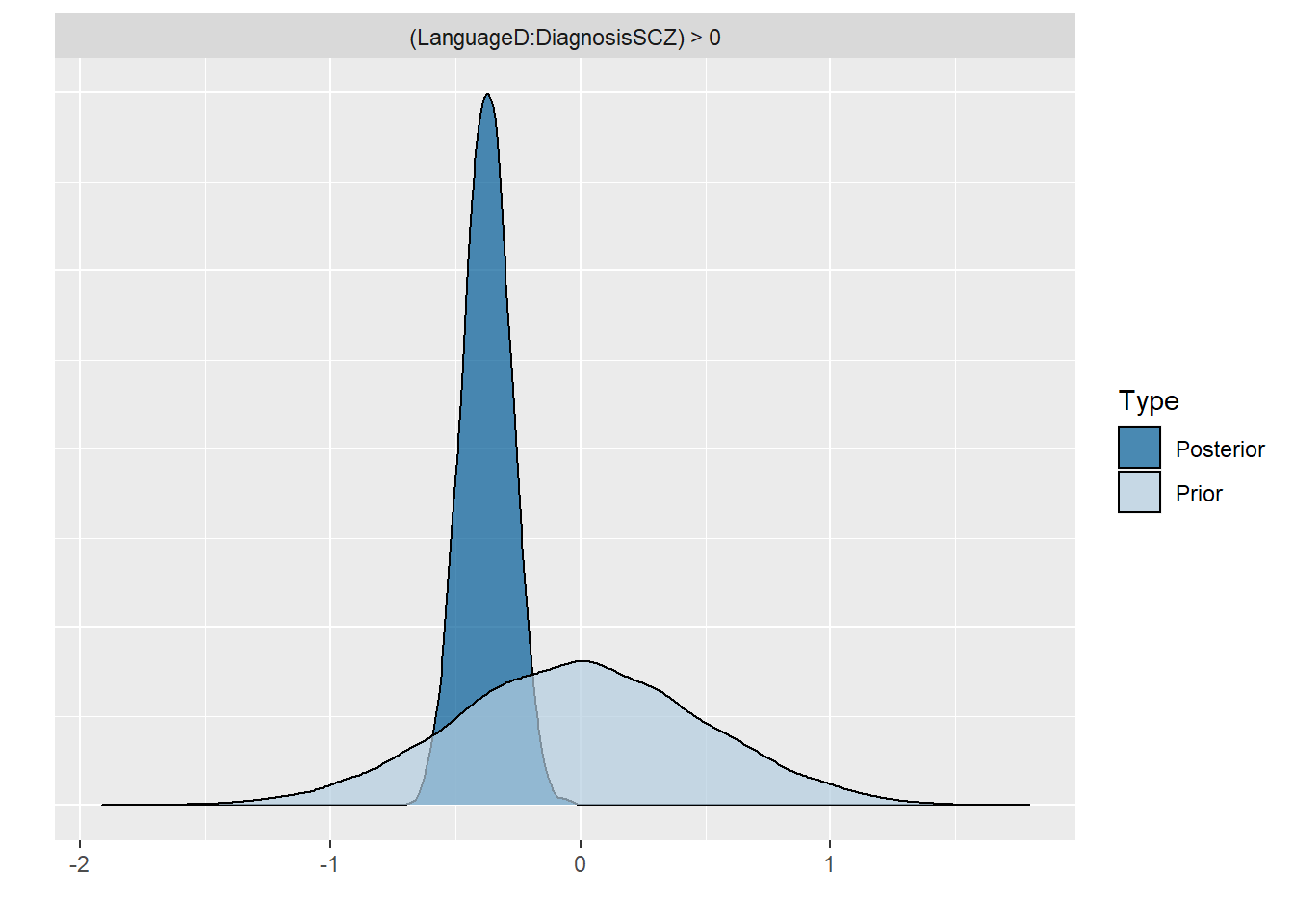

**Figure S4_D.** Plot of the posterior predictive check. We overlay the distribution of the actual data for the speech rate feature (bold blue line) with the distribution of 100 simulations from the fitted model (predictions, each simulation a pale blue line). This allows us to identify potential biases in the model.

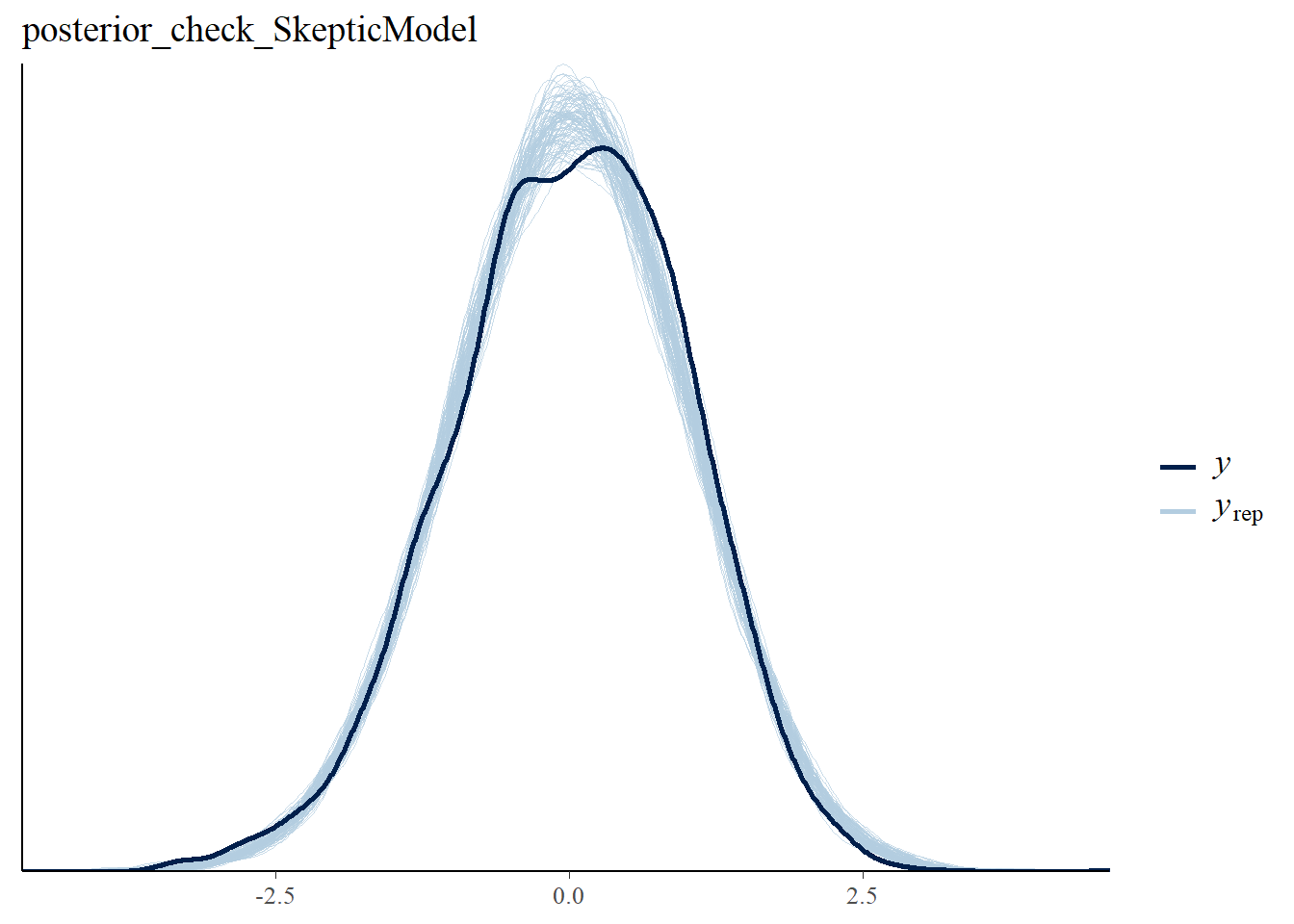

**Model fitting**

The models were fitted using Hamiltonian MonteCarlo samplers with 2 parallel chains with 2,000 iterations each, an adapt delta of 0.99 and a maximum tree depth of 20 in order to ensure no divergence in the estimation process. The quality of the models was assessed beyond the prior predictive checks and prior-posterior update checks discussed in the previous paragraph. We performed posterior predictive checks, akin to traditional residual checks, to assess whether the model was able to capture the empirical distribution of the data and did not present obvious biases, such as overestimating low values. We made sure that R^ statistics were lower than 1.05, that is, that the independent Markov chains used to identify the posterior distributions converged on their estimates. We ensured that no divergences were generated in the process of estimation, that is, that the possible parameter values were satisfactorily explored, without specific ranges of values being excessively difficult to evaluate. We checked that the number of effective bulk and tail samples was above 200, that is, that the model fitting process was able to explore the possible parameter values and was able to sample at least 200 independent possible values. See figures below for predictive checks, and prior-posterior update checks.

**Model assessment**

We then reported the estimated difference by group in terms of mean difference separately by language, 95% Compatibility Intervals (CIs, indicating the probable range of difference, assuming the model is correct) and Evidence Ratio (ER, evidence in favor of the effect observed against alternative hypotheses). When ER was weak (below 10, that is, ten times as much evidence for the effect as for alternative hypotheses), we also calculated the ER in favor of the null hypothesis. Note that given the standardization of the outcome variables, the effect size is equivalent to Hedges’ g, that is, is expressed in units of standard deviations. To assess the effects of using previous literature informed and skeptical priors, we report the same model estimates for both models. Further, we adopted a Leave-One-Out model comparison framework estimating the model’s out-of-sample performance, in other words, estimating the ability of the model to generalize to new data (. We then calculated their relative stacking weight based on Leave-One-Out Information Criteria, assessing the probability of each model to be better than the others^31^. This procedure informs us as to whether adding information from previous findings in our statistical models enabled us to create more robust models, with greater chances of having replicable findings in new studies.

**Analysis on the association between clinical symptoms and acoustic features**

For ease of comparison with the previous literature cited in the meta-analysis reporting Pearson correlation coefficients, we scaled the acoustic features separately for each language, that is, we first subtracted the minimum values and divided the result by range, i.e. the difference between the maximum and minimum values, to bring all values into the zero to one range.

We chose not to standardize the clinical features as z-scores, as that would assume a linear relation between acoustic and clinical features. Clinical features are more adequately modeled as ordinal variables, where a monotonic relation is assumed but non-linear forms are possible. In other words, while we expect that if there is a change in acoustic patterns when moving from a SANS score of 0 to a score of 1, we should see a change in the same direction when moving from 1 to 2, but the size of the change might be different. Note that in case of linear changes, the current model gives comparable estimates to more traditional models^32^.

This analysis was performed on the SCZ group only. Indeed, even if clinical ratings were available for many of the HCs, the SANS scale is designed specifically for assessing symptomatology in individuals with neuropsychiatric disorders and not for capturing individual variability in healthy individuals. As a result, healthy controls present a median score of 0 for the different ratings and minimal variability in scores, which potentially introduces a bias into the model. Finally, informed priors from the previous meta-analysis were available only for patients with SCZ.

Skeptical priors were specified as normal distribution centered at zero (no association between acoustic features and clinical ratings), and a standard deviation of .3. The expected estimates were thus predominantly included within -0.9 and 0.9, but could still be swayed by the data. Intercepts were specified as normal distribution centered at 0.5 and a standard deviation of 0.3. Individual variability was modeled using a positive half-normal prior centered at 0, with a standard deviation of 0.1 from the estimate for the specific group and language of the individual, thus regularizing the inference.

Note that model quality checks (done before actually assessing the results see Model fitting section and see Figure **S4_*E,* S4_*F,* S4_*G,* S4_*H***, in particular prior posterior comparisons, indicated that informed priors were too confident in their expectations and strongly limited learning from data. We therefore increased uncertainty by multiplying the standard deviation by 3, upon which the model was better able to also learn from the data, the variance in the posteriors being much lower than in the priors^33^.

We then compared results across the informed and skeptical models following the same procedure described in the previous paragraphs.

**Figure S4_E**. Plot of the prior predictive checks (light blue) and observed data (bold blue) for the relationship between symptoms (SANS Global score) and acoustic features. Note that the four language groups (DK, GE, JP, CH) are given the same prior expectations for all parameters, thus implicitly assuming no difference between them (so that any evidence for a difference that we might find would come exclusively from the data and not from the priors).

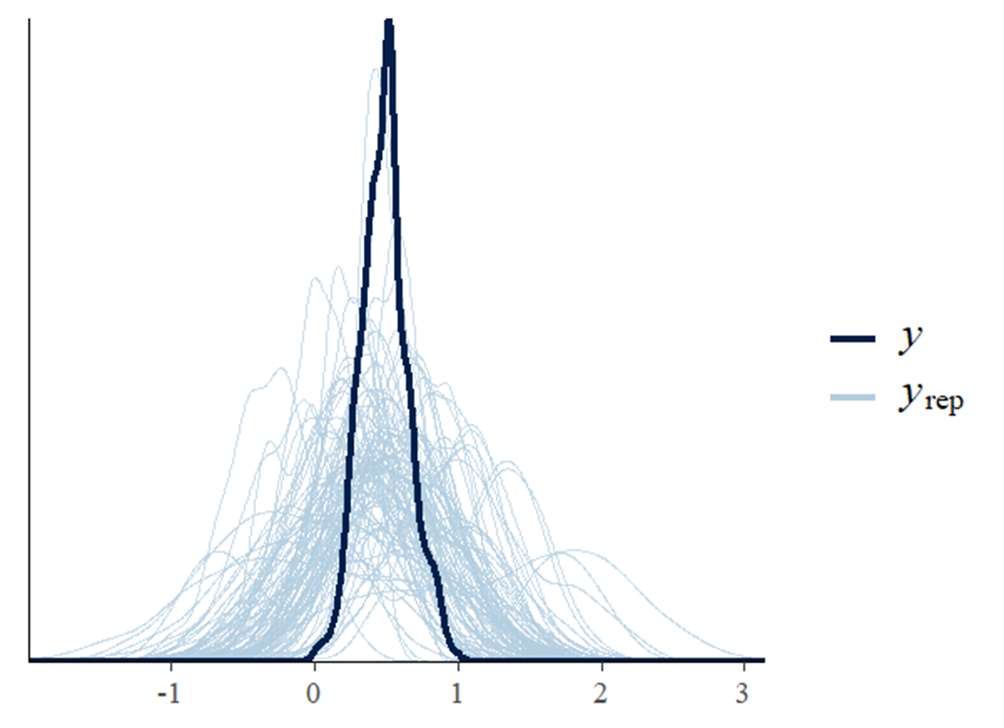

**FIGURE S4_F.** Plot of the prior-posterior predictive check. Prior (light blue) and posterior (dark blue) distributions *for the intercept* in the model *for the Danish language* for the relationship between symptoms (SANS Global alogia score) and acoustic features (speech rate).

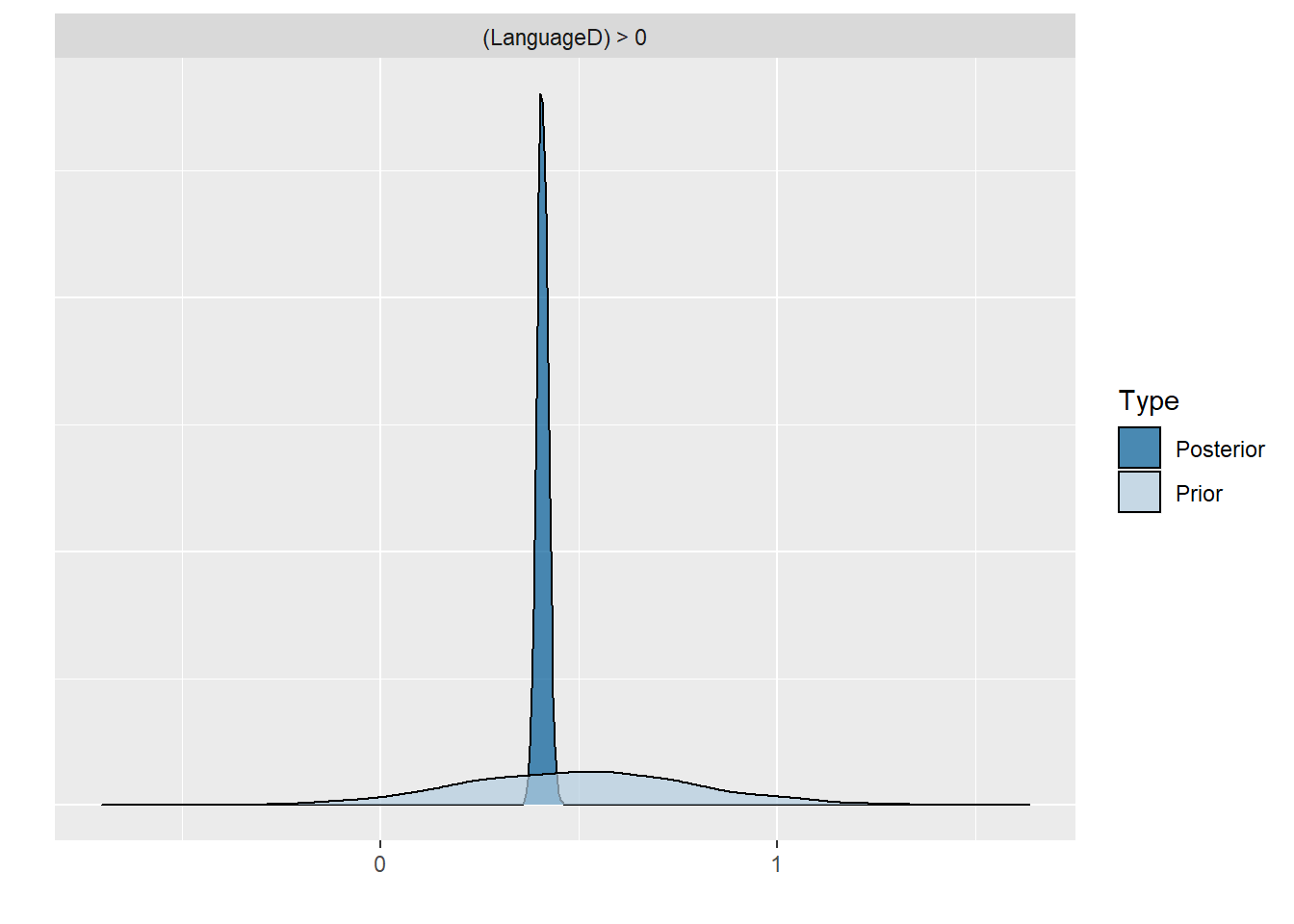

**FIGURE S4_G.** Plot of the prior*-posterior predictive check. P*rior (light blue) and posterior (dark blue) distributions for the effect of diagnosis (SCZ-HC) for the Danish language for the relationship between symptoms (SANS alogia score) and acoustic features (speech rate).

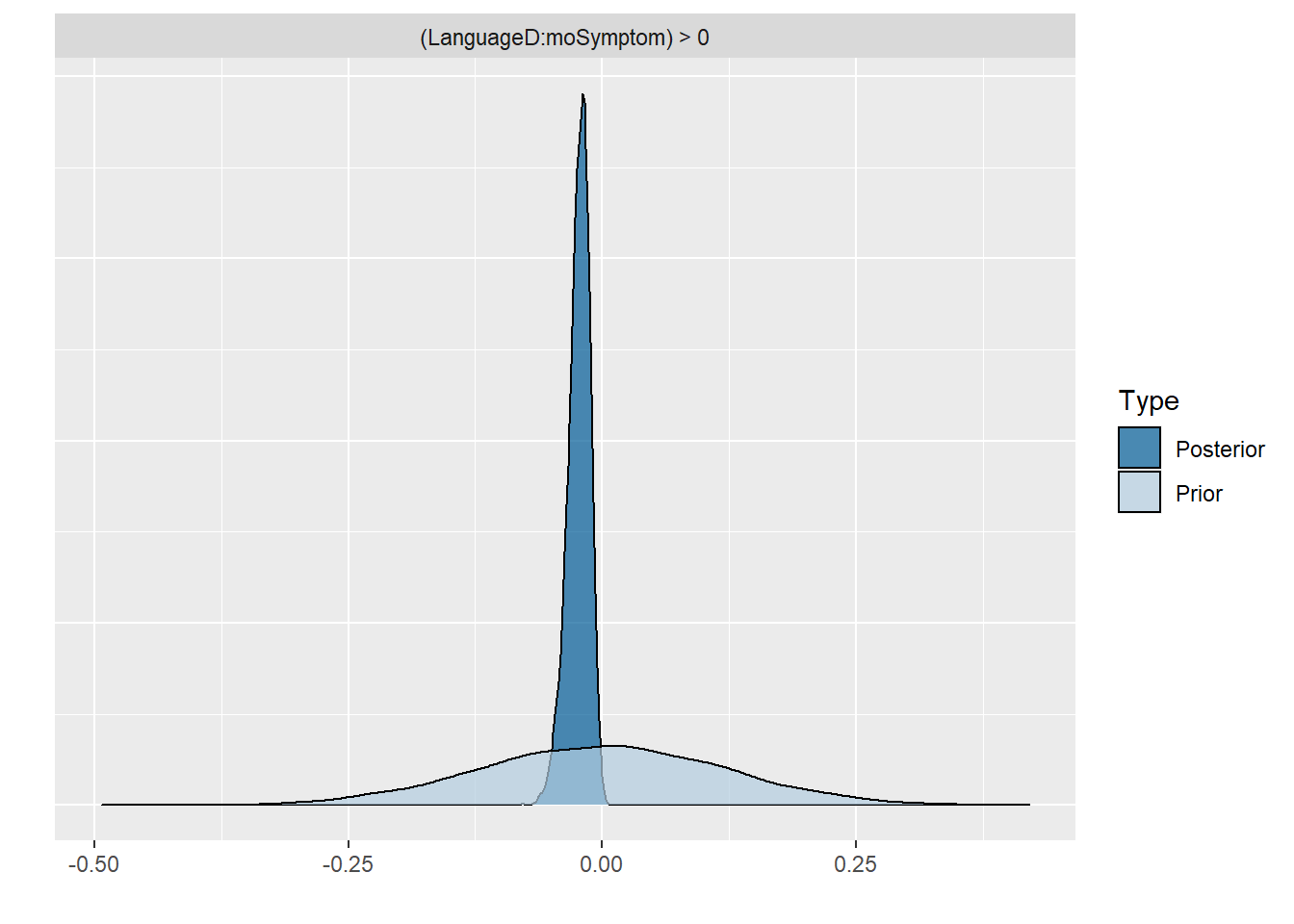

**Figure S4_H.** Plot of the posterior predictive check. We overlay the distribution of the actual data for the relationship between symptoms (SANS Global alogia score) and coherence measures (Speech rate) with the distribution of 100 simulations from the fitted model (predictions, each simulation a pale blue line). This allows us to identify potential biases in the model.

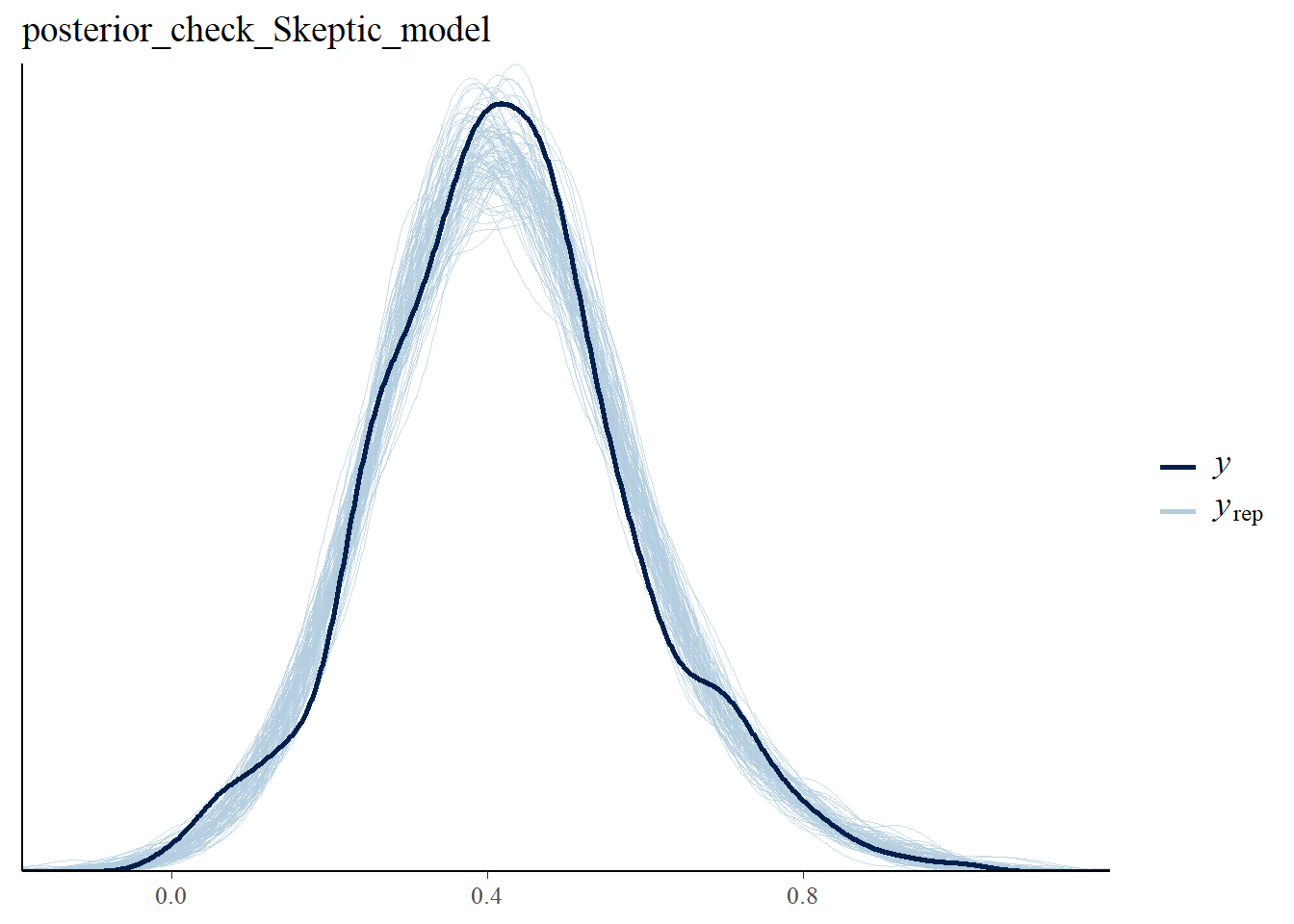

**S5 – Novel acoustic features**

**S5.1 Effect of diagnosis**

The detailed results are reported in Table **S5_A**. Generally, we found some evidence for reduced formants median frequency and formants variability, and for increased median relative amplitude (H1H2) and reduced H1H2 variability. However, these findings are small and not robust across languages. Results for the other novel features are very uncertain or inconsistent across languages. As for more traditional features, we found that gender, age, and level of intelligence does in some cases affect the differences.

**Table S5_A**

| **Acoustic Features** | **Group (CT – SCZ)** | **Group * Gender (M-F)** |
| --- | --- | --- |
| **F1_Median** |  |  |
| Skeptical DK | -0.13 (-0.36 0.08) ER = 5.58 | 0.33 (-0.04 0.7) ER = 12.85 |
| Skeptical CH | **-0.27 (-0.54 -0.01) ER = 21.22** | 0.41 (-0.09 0.91) ER = 10.12 |
| Skeptical JP | **-0.56 (-0.95 -0.17) ER = 88.29** | -0.15 (-0.89 0.59) ER = 1.71 ER01 = 1.54 |
| Skeptical GE | -0.1 (-0.35 0.15) ER = 3 ER01 = 2.57 | -0.11 (-0.54 0.31) ER = 1.98 ER01 = 2.48 |
| **F1_IQR** |  |  |
| Skeptical DK | -**0.23 (-0.42 -0.04) ER = 38.37** | 0.21 (-0.17 0.57) ER = 4.73 |
| Skeptical CH | -0.16 (-0.41 0.08) ER = 6.38 | 0.12 (-0.29 0.53) ER = 2.18 ER01 = 1.9 |
| Skeptical JP | 0.03 (-0.34 0.41) ER = 1.25 ER01 = 2.26 | 0.19 (-0.49 0.88) ER = 2.09 ER01 = 1.6 |
| Skeptical GE | -0.05 (-0.29 0.19) ER = 1.86 ER01 = 3.17 | 0.16 (-0.26 0.59) ER = 2.76 ER01 = 2.26 |
| **F2_Median** |  |  |
| Skeptical DK | -0.11 (-0.33 0.11) ER = 3.96 | 0.33 (-0.08 0.74) ER = 10.21 |
| Skeptical CH | **-0.21 (-0.46 0.04) ER = 11.25** | -0.08 (-0.56 0.4) ER = 1.56 ER01 = 1.32 |
| Skeptical JP | 0.3 (-0.11 0.71) ER = 8.19 | 0.35 (-0.39 1.08) ER = 3.72 |
| Skeptical GE | -0.17 (-0.4 0.07) ER = 7.27 | -0.02 (-0.47 0.42) ER = 1.11 ER01 = 2.57 |
| **F2_IQR** |  |  |
| Skeptical DK | **-0.2 (-0.4 0.01) ER = 16.99** | 0.16 (-0.22 0.54) ER = 3.14 |
| Skeptical CH | -0.18 (-0.44 0.08) ER = 7.17 | 0.06 (-0.36 0.51) ER = 1.43 ER01 = 1.46 |
| Skeptical JP | -0.24 (-0.67 0.2) ER = 4.62 | 0.12 (-0.53 0.76) ER = 1.66 ER01 = 1.68 |
| Skeptical GE | -0.04 (-0.24 0.16) ER = 1.76 ER01 = 3.6 | 0.29 (-0.11 0.68) ER = 7.47 |
| **F3_Median** |  |  |
| Skeptical DK | -0.09 (-0.3 0.12) ER = 3.36 | 0.32 (-0.1 0.74) ER = 8.89 |
| Skeptical CH | **-0.34 (-0.61 -0.08) ER = 61.5** | -0.34 (-0.82 0.16) ER = 6.5 |
| Skeptical JP | 0.13 (-0.31 0.55) ER = 2.22 ER01 = 1.63 | -0.15 (-0.89 0.59) ER = 1.64 ER01 = 1.51 |
| Skeptical GE | -0.17 (-0.41 0.06) ER = 7.82 | 0.01 (-0.44 0.46) ER = 1.04 ER01 = 2.7 |
| **F3_IQR** |  |  |
| Skeptical DK | -0.14 (-0.35 0.08) ER = 5.83 | 0.21 (-0.22 0.62) ER = 3.61 |
| Skeptical CH | **0.25 (-0.01 0.51) ER = 16.01** | -0.1 (-0.47 0.28) ER = 2.07 ER01 = 0.95 |
| Skeptical JP | -0.24 (-0.67 0.2) ER = 4.78 | -0.33 (-0.98 0.34) ER = 3.89 |
| Skeptical GE | 0 (-0.22 0.22) ER = 1.01 ER01 = 3.73 | 0.03 (-0.41 0.46) ER = 1.19 ER01 = 2.7 |
| **F4_Median** |  |  |
| Skeptical DK | -0.13 (-0.36 0.09) ER = 5.55 | 0.3 (-0.13 0.72) ER = 7.32 |
| Skeptical CH | **-0.47 (-0.72 -0.23) ER = 2499** | -0.09 (-0.55 0.39) ER = 1.64 ER01 = 0.02 |
| Skeptical JP | 0.14 (-0.27 0.55) ER = 2.52 ER01 = 1.75 | -0.14 (-0.88 0.6) ER = 1.59 ER01 = 1.53 |
| Skeptical GE | -0.1 (-0.33 0.13) ER = 3.11 | 0.18 (-0.28 0.65) ER = 2.78 ER01 = 2.18 |
| **F4_IQR** |  |  |
| Skeptical DK | **-0.2 (-0.41 0) ER = 18.53** | 0.32 (-0.07 0.71) ER = 10.03 |
| Skeptical CH | 0.03 (-0.22 0.28) ER = 1.36 ER01 = 3.38 | -0.62 (-1.07 -0.16) ER = 75.92 |
| Skeptical JP | 0 (-0.39 0.39) ER = 1.02 ER01 = 2.06 | 0 (-0.72 0.74) ER = 0.98 ER01 = 1.6 |
| Skeptical GE | 0.07 (-0.15 0.29) ER = 2.56 ER01 = 3.25 | -0.08 (-0.45 0.29) ER = 1.82 ER01 = 3.01 |
| **F5_Median** |  |  |
| Skeptical DK | -0.14 (-0.36 0.08) ER = 6.84 | 0.29 (-0.12 0.69) ER = 7.3 |
| Skeptical CH | **-0.41 (-0.65 -0.16) ER = 415.67** | 0.13 (-0.34 0.59) ER = 2.08 ER01 = 0.08 |
| Skeptical JP | -0.02 (-0.36 0.33) ER = 1.16 ER01 = 2.45 | -0.51 (-1.18 0.17) ER = 8.07 |
| Skeptical GE | -0.09 (-0.3 0.13) ER = 2.99 ER01 = 2.98 | 0.24 (-0.16 0.65) ER = 5.15 |
| **F5_IQR** |  |  |
| Skeptical DK | -0.14 (-0.34 0.07) ER = 6.32 | 0.13 (-0.27 0.53) ER = 2.4 ER01 = 2.5 |
| Skeptical CH | -0.02 (-0.26 0.23) ER = 1.17 ER01 = 3.44 | 0.33 (-0.12 0.78) ER = 7.8 |
| Skeptical JP | -0.14 (-0.47 0.19) ER = 3.14 | -0.01 (-0.66 0.64) ER = 1.03 ER01 = 1.71 |
| Skeptical GE | 0.13 (-0.07 0.33) ER = 5.6 | -0.1 (-0.51 0.31) ER = 1.83 ER01 = 2.6 |
| **H1H2_Median** |  |  |
| Skeptical DK | **0.22 (0.03 0.41) ER = 37.02** | -0.19 (-0.47 0.08) ER = 6.84 |
| Skeptical CH | **0.29 (-0.03 0.61) ER = 13.77** | -0.13 (-0.54 0.29) ER = 2.27 ER01 = 0.93 |
| Skeptical JP | -0.32 (-0.79 0.16) ER = 6.86 | 1.28 (0.5 2) ER = 168.49 |
| Skeptical GE | 0.03 (-0.23 0.29) ER = 1.34 ER01 = 3.41 | 0.16 (-0.29 0.6) ER = 2.71 ER01 = 2.15 |
| **H1H2_IQR** |  |  |
| Skeptical DK | **-0.19 (-0.33 -0.04) ER = 45.95** | 0.37 (0.09 0.66) ER = 60.73 |
| Skeptical CH | **-0.39 (-0.64 -0.13) ER = 174.44** | 0.18 (-0.3 0.65) ER = 2.75 ER01 = 0.14 |
| Skeptical JP | 0.09 (-0.33 0.51) ER = 1.83 ER01 = 1.82 | -0.33 (-1.07 0.41) ER = 3.41 |
| Skeptical GE | -0.01 (-0.23 0.21) ER = 1.22 ER01 = 3.66 | -0.16 (-0.52 0.2) ER = 3.25 |
| **QOQ_Median** |  |  |
| Skeptical DK | 0.09 (-0.08 0.26) ER = 3.82 | 0.12 (-0.13 0.37) ER = 3.55 |
| Skeptical CH | 0.01 (-0.25 0.27) ER = 1.15 ER01 = 3.22 | 0.32 (-0.15 0.81) ER = 6.39 |
| Skeptical JP | 0.03 (-0.36 0.41) ER = 1.3 ER01 = 2.16 | 0.96 (0.23 1.63) ER = 55.18 |
| Skeptical GE | 0.12 (-0.1 0.34) ER = 4.68 | 0.02 (-0.4 0.44) ER = 1.16 ER01 = 2.65 |
| **QOQ_IQR** |  |  |
| Skeptical DK | -0.01 (-0.17 0.15) ER = 1.14 ER01 = 5.13 | 0.09 (-0.15 0.34) ER = 2.66 ER01 = 3.73 |
| Skeptical CH | 0.03 (-0.2 0.26) ER = 1.47 ER01 = 3.46 | 0.12 (-0.32 0.57) ER = 2.03 ER01 = 3.46 |
| Skeptical JP | -0.33 (-0.66 0.03) ER = 14.43 | 0.42 (-0.29 1.1) ER = 5.21 |
| Skeptical GE | -0.04 (-0.22 0.14) ER = 1.7 ER01 = 4.14 | -0.27 (-0.63 0.08) ER = 8.94 |
| **NAQ_Median** |  |  |
| Skeptical DK | 0.1 (-0.07 0.26) ER = 5.01 | 0.13 (-0.13 0.39) ER = 3.95 |
| Skeptical CH | -0.01 (-0.27 0.26) ER = 1.15 ER01 = 3.18 | 0.34 (-0.12 0.82) ER = 7.64 |
| Skeptical JP | -0.02 (-0.38 0.33) ER = 1.22 ER01 = 2.33 | 1.08 (0.42 1.67) ER = 162.93 |
| Skeptical GE | 0.09 (-0.12 0.31) ER = 3.29 | 0.09 (-0.35 0.52) ER = 1.77 ER01 = 2.54 |
| **NAQ_IQR** |  |  |
| Skeptical DK | 0.1 (-0.07 0.27) ER = 5.37 | 0.06 (-0.17 0.3) ER = 2.01 ER01 = 4.62 |
| Skeptical CH | -0.03 (-0.28 0.22) ER = 1.41 ER01 = 3.28 | 0.19 (-0.27 0.65) ER = 3.01 |
| Skeptical JP | -0.37 (-0.72 0) ER = 19.33 | 0.85 (0.12 1.55) ER = 33.97 |
| Skeptical GE | 0.06 (-0.14 0.26) ER = 2.25 ER01 = 3.59 | -0.01 (-0.4 0.37) ER = 1.1 ER01 = 2.91 |
| **HNR_Median** |  |  |
| Skeptical DK | **0.26 (0.08 0.44) ER = 127.21** | -0.01 (-0.34 0.33) ER = 1.16 ER01 = 3.47 |
| Skeptical CH | **0.42 (0.14 0.7) ER = 150.52** | 0.03 (-0.46 0.52) ER = 1.17 ER01 = 0.13 |
| Skeptical JP | -0.19 (-0.59 0.21) ER = 3.77 | 0.27 (-0.47 1) ER = 2.69 ER01 = 1.33 |
| **Skeptical GE** | **-0.28 (-0.53 -0.03) ER = 28.07** | 0.13 (-0.34 0.59) ER = 2.08 ER01 = 2.2 |
| **HNR_IQR** |  |  |
| Skeptical DK | **0.35 (0.16 0.52) ER = 2499** | -0.13 (-0.39 0.14) ER = 3.87 |
| Skeptical CH | 0.09 (-0.2 0.38) ER = 2.13 ER01 = 2.52 | 0.49 (0.01 0.96) ER = 20.98 |
| Skeptical JP | 0.26 (-0.21 0.71) ER = 4.87 | 0.85 (-0.03 1.61) ER = 17.18 |
| Skeptical GE | 0.26 (0.02 0.5) ER = 27.9 | 0.37 (-0.05 0.79) ER = 12.33 |
| **PSP_Median** |  |  |
| Skeptical DK | -0.1 (-0.28 0.08) ER = 4.46 | -0.17 (-0.47 0.13) ER = 4.77 |
| Skeptical CH | -0.18 (-0.43 0.06) ER = 7.73 | 0.07 (-0.39 0.53) ER = 1.58 ER01 = 1.66 |
| Skeptical JP | 0.22 (-0.12 0.55) ER = 5.95 | -0.86 (-1.42 -0.27) ER = 102.09 |
| Skeptical GE | 0.03 (-0.18 0.24) ER = 1.47 ER01 = 3.74 | 0.09 (-0.3 0.47) ER = 1.85 ER01 = 2.82 |
| **MDQ_Median** |  |  |
| Skeptical DK | 0.09 (-0.1 0.28) ER = 3.42 | -0.06 (-0.41 0.28) ER = 1.62 ER01 = 3.34 |
| Skeptical CH | **0.21 (-0.04 0.46) ER = 10.7** | 0.66 (0.28 1.02) ER = 499 |
| Skeptical JP | -**0.45 (-0.89 -0.02) ER = 21.62** | 0.06 (-0.66 0.79) ER = 1.21 ER01 = 1.56 |
| Skeptical GE | -0.04 (-0.28 0.18) ER = 1.65 ER01 = 3.62 | 0 (-0.41 0.42) ER = 0.99 ER01 = 2.76 |
| **MDQ_IQR** |  |  |
| Skeptical DK | -0.02 (-0.18 0.13) ER = 1.43 ER01 = 4.99 | -0.37 (-0.66 -0.07) ER = 39.32 |
| Skeptical CH | -0.04 (-0.28 0.21) ER = 1.48 ER01 = 3 | -0.19 (-0.67 0.29) ER = 2.94 ER01 = 3 |
| Skeptical JP | -0.06 (-0.4 0.28) ER = 1.67 ER01 = 2.22 | -0.72 (-1.33 -0.06) ER = 26.93 |
| Skeptical GE | -0.02 (-0.22 0.17) ER = 1.36 ER01 = 4.09 | -0.06 (-0.44 0.35) ER = 1.5 ER01 = 2.81 |

**S5.2 Relation between novel acoustic features and clinical symptoms**

Clinical features generally correlate with novel acoustic features, in particular formants median and IQR are negatively related to SANS and SAPS ratings, and to alogia and flat affect ratings. However, we did not find reliable and robust association across all languages and scales (see **Table S5_B**).

**Table S5_B**

| **Rating scales** | **SANS** | **SAPS** | **SANS - Alogia** | **SANS - Flat affect** |  |
| --- | --- | --- | --- | --- | --- |
| **F1 Median** |  |  |  |  |  |
| Skeptical DK | **-0.05 (-0.1 -0.01) ER = 28.7** | **-0.06 (-0.1 -0.02) ER = 499** | -0.02 (-0.05 0.02) ER = 3.26 | 0 (-0.03 0.04) ER = 1.41 ER01 = 28.46 |  |
| Skeptical CH | 0.04 (-0.13 0.22) ER = 1.78 ER01 = 5.46 | **-0.1 (-0.21 0) ER = 18.35** | **-0.24 (-0.48 -0.04) ER = 41.86** | **-0.14 (-0.28 0) ER = 21.3** |  |
| **F1 IQR** |  |  |  |  |  |
| Skeptical DK | -0.04 (-0.12 0.04) ER = 3.22 | **-0.1 (-0.17 -0.03) ER = 126.66** | **0 (-0.07 0.07) ER = 1.07 ER01 = 14.97** | 0 (-0.07 0.06) ER = 1.1 ER01 = 15.7 |  |
| Skeptical CH | 0.1 (-0.15 0.36) ER = 2.64 ER01 = 3.51 | -0.05 (-0.21 0.12) ER = 2.07 ER01 = 5.64 | -0.08 (-0.44 0.27) ER = 1.82 ER01 = 2.97 | **-0.19 (-0.44 0.03) ER = 11.15** |  |
| **F2 Median** |  |  |  |  |  |
| Skeptical DK | -0.05 (-0.15 0.04) ER = 4.17 | **-0.1 (-0.19 -0.02) ER = 46.24** | -0.01 (-0.1 0.07) ER = 1.56 ER01 = 11.34 | -0.01 (-0.09 0.07) ER = 1.3 ER01 = 13.52 |  |
| Skeptical CH | 0.03 (-0.17 0.22) ER = 1.46 ER01 = 5.13 | **-0.14 (-0.27 -0.03) ER = 49.85** | -0.16 (-0.43 0.09) ER = 6.07 | -0.06 (-0.23 0.11) ER = 2.5 ER01 = 5.04 |  |
| **F2 IQR** |  |  |  |  |  |
| Skeptical DK | **-0.13 (-0.27 0.02) ER = 12.92** | **-0.2 (-0.33 -0.08) ER = 1999** | -0.07 (-0.19 0.05) ER = 4.55 | -0.01 (-0.12 0.1) ER = 1.28 ER01 = 9.28 |  |
| Skeptical CH | 0.01 (-0.22 0.24) ER = 1.17 ER01 = 4.93 | -0.06 (-0.21 0.08) ER = 2.97 ER01 = 5.13 | -0.06 (-0.38 0.24) ER = 1.66 ER01 = 3.39 | -0.03 (-0.25 0.18) ER = 1.47 ER01 = 4.52 |  |
| **F3 Median** |  |  |  |  |  |
| Skeptical DK | -0.09 (-0.27 0.06) ER = 4.85 | **-0.19 (-0.32 -0.06) ER = 199** | 0 (-0.13 0.13) ER = 1.16 ER01 = 8.58 | -0.01 (-0.13 0.11) ER = 1.31 ER01 = 7.96 |  |
| Skeptical CH | 0.02 (-0.2 0.25) ER = 1.29 ER01 = 4.3 | **-0.13 (-0.27 0) ER = 16.96** | -0.07 (-0.34 0.2) ER = 2.2 ER01 = 3.42 | 0.02 (-0.19 0.22) ER = 1.44 ER01 = 4.66 |  |
| **F3 IQR** |  |  |  |  |  |
| Skeptical DK | -0.12 (-0.29 0.04) ER = 8.1 | -0.12 (-0.29 0.04) ER = 8.1 | -0.01 (-0.16 0.13) ER = 1.26 ER01 = 6.87 | 0 (-0.13 0.13) ER = 1.04 ER01 = 7.44 |  |
| Skeptical CH | -0.19 (-0.46 0.07) ER = 8.84 | -0.19 (-0.46 0.07) ER = 8.84 | -0.21 (-0.58 0.11) ER = 5.93 | -0.17 (-0.42 0.09) ER = 6.45 |  |
| **F4 Median** |  |  |  |  |  |
| Skeptical DK | -0.12 (-0.34 0.07) ER = 6.01 | **-0.25 (-0.42 -0.08) ER = 145.34** | -0.03 (-0.2 0.14) ER = 1.57 ER01 = 5.57 | -0.03 (-0.18 0.12) ER = 1.87 ER01 = 6.23 |  |
| Skeptical CH | 0.16 (-0.04 0.39) ER = 10.01 | -0.02 (-0.14 0.11) ER = 1.5 ER01 = 7.86 | -0.08 (-0.34 0.17) ER = 2.58 ER01 = 3.69 | 0.08 (-0.09 0.25) ER = 3.95 |  |
| **F4 IQR** |  |  |  |  |  |
| Skeptical DK | -0.07 (-0.19 0.04) ER = 5.79 | **-0.14 (-0.24 -0.04) ER = 114.38** | -0.01 (-0.12 0.09) ER = 1.33 ER01 = 9.78 | 0 (-0.1 0.1) ER = 1.02 ER01 = 10.92 |  |
| Skeptical CH | -0.04 (-0.21 0.14) ER = 1.81 ER01 = 5.7 | 0.08 (-0.03 0.18) ER = 8.51 | **-0.21 (-0.45 -0.01) ER = 21.73** | **-0.11 (-0.26 0.02) ER = 10.45** |  |
| **F5 Median** |  |  |  |  |  |
| Skeptical DK | -0.13 (-0.34 0.07) ER = 5.72 | **-0.28 (-0.46 -0.1) ER = 399** | -0.01 (-0.18 0.17) ER = 1.13 ER01 = 5.89 | -0.03 (-0.2 0.15) ER = 1.58 ER01 = 5.33 |  |
| Skeptical CH | 0.12 (-0.1 0.37) ER = 4.19 | **-0.21 (-0.36 -0.07) ER = 95.77** | 0 (-0.13 0.13) ER = 0.99 ER01 = 7.73 | 0 (-0.21 0.22) ER = 0.98 ER01 = 5.4 |  |
| **F5 IQR** |  |  |  |  |  |
| Skeptical DK | -0.12 (-0.31 0.05) ER = 6.83 | -0.25 (-0.39 -0.12) ER = 999 | -0.04 (-0.18 0.11) ER = 2.11 ER01 = 6.64 | 0 (-0.13 0.13) ER = 1.05 ER01 = 7.53 |  |
| Skeptical CH | -0.07 (-0.26 0.13) ER = 2.88 ER01 = 4.67 | -0.13 (-0.25 -0.02) ER = 28.27 | -0.15 (-0.37 0.06) ER = 8.05 | **-0.11 (-0.26 0.02) ER = 10.41** |  |
| **H1H2 Median** |  |  |  |  |  |
| Skeptical DK | 0.08 (-0.09 0.26) ER = 3.49 | 0.11 (-0.03 0.26) ER = 8.79 | **0.17 (0.01 0.33) ER = 27.99** | 0.01 (-0.12 0.15) ER = 1.3 ER01 = 7.38 |  |
| Skeptical CH | -0.02 (-0.3 0.28) ER = 1.25 ER01 = 3.73 | **0.15 (-0.01 0.32) ER = 15.09** | 0.27 (-0.14 0.7) ER = 6.45 | 0.05 (-0.24 0.34) ER = 1.56 ER01 = 3.48 |  |
| **H1H2 IQR** |  |  |  |  |  |
| Skeptical DK | -0.04 (-0.11 0.02) ER = 7.81 | **-0.07 (-0.12 -0.02) ER = 72.17** | **-0.08 (-0.13 -0.03) ER = 170.43** | 0.01 (-0.04 0.06) ER = 1.56 ER01 = 19.47 |  |
|  | 0.08 (-0.1 0.27) ER = 3.47 | -0.07 (-0.19 0.05) ER = 5.15 | 0.13 (-0.14 0.44) ER = 3.81 | 0.04 (-0.15 0.23) ER = 1.85 ER01 = 5.04 |  |
| **QOQ Median** |  |  |  |  |  |
| Skeptical DK | 0.05 (-0.13 0.24) ER = 1.9 ER01 = 5.55 | 0.07 (-0.08 0.22) ER = 3.72 | **0.14 (-0.02 0.3) ER = 12.04** | -0.02 (-0.16 0.12) ER = 1.38 ER01 = 7.31 |  |
| Skeptical CH | 0.23 (-0.12 0.6) ER = 5.85 | **0.21 (-0.02 0.44) ER = 14.08** | **0.45 (0.01 0.92) ER = 20.28** | 0.1 (-0.25 0.45) ER = 2.35 ER01 = 2.66 |  |
| **QOQ IQR** |  |  |  |  |  |
| Skeptical DK | 0.04 (-0.06 0.15) ER = 2.96 ER01 = 8.11 | 0.03 (-0.06 0.12) ER = 2.81 ER01 = 8.74 | 0.04 (-0.05 0.13) ER = 3.35 | 0.01 (-0.08 0.09) ER = 1.39 ER01 = 11.24 |  |
| Skeptical CH | -0.02 (-0.21 0.17) ER = 1.43 ER01 = 5.37 | 0.08 (-0.04 0.19) ER = 6.33 | -0.04 (-0.25 0.18) ER = 1.69 ER01 = 4.75 | -0.09 (-0.24 0.04) ER = 6.99 |  |
| **NAQ Median** |  |  |  |  |  |
| Skeptical DK | 0.03 (-0.13 0.2) ER = 1.75 ER01 = 5.72 | 0.06 (-0.08 0.2) ER = 2.89 ER01 = 5.83 | **0.15 (-0.01 0.32) ER = 16.39** | 0 (-0.14 0.14) ER = 1.02 ER01 = 6.86 |  |
| Skeptical CH | 0.21 (-0.14 0.59) ER = 5.07 | **0.2 (-0.03 0.43) ER = 13.02** | **0.51 (0.09 0.96) ER = 45.88** | 0.14 (-0.2 0.49) ER = 3.15 |  |
| **NAQ IQR** |  |  |  |  |  |
| Skeptical DK | 0.06 (-0.07 0.21) ER = 3.49 | 0.05 (-0.06 0.16) ER = 3.5 | **0.14 (0.02 0.28) ER = 36.97** | 0.03 (-0.07 0.14) ER = 2.28 ER01 = 8.64 |  |
| Skeptical CH | 0.03 (-0.22 0.28) ER = 1.36 ER01 = 4.36 | **0.18 (0.03 0.34) ER = 36.27** | **0.27 (-0.02 0.61) ER = 14.79** | 0.06 (-0.16 0.29) ER = 2.04 ER01 = 4.22 |  |
| **HNR Median** |  |  |  |  |  |
| Skeptical DK | **-0.13 (-0.24 -0.03) ER = 59** | 0.04 (-0.06 0.14) ER = 3.3 | -0.04 (-0.13 0.05) ER = 2.87 ER01 = 9 | **-0.09 (-0.17 -0.01) ER = 23.29** |  |
| Skeptical CH | **-0.26 (-0.57 0.01) ER = 15.9** | **0.25 (0.07 0.44) ER = 119** | -0.25 (-0.64 0.12) ER = 7.02 | -0.13 (-0.41 0.14) ER = 3.69 |  |
| **HNR IQR** |  |  |  |  |  |
| Skeptical DK | **-0.03 (-0.07 0.01) ER = 11.32** | 0.02 (-0.01 0.06) ER = 6.74 | **-0.03 (-0.07 0) ER = 16.75** | 0.01 (-0.03 0.04) ER = 1.44 ER01 = 26.12 |  |
| Skeptical CH | -0.04 (-0.31 0.21) ER = 1.43 ER01 = 4.22 | **-0.17 (-0.34 0) ER = 20.98** | 0.01 (-0.21 0.23) ER = 1.07 ER01 = 4.56 | -0.01 (-0.36 0.33) ER = 1.05 ER01 = 3.07 |  |
| **PSP Median** |  |  |  |  |  |
| Skeptical DK | -0.02 (-0.16 0.13) ER = 1.35 ER01 = 6.68 | 0.09 (-0.04 0.22) ER = 6.58 | -0.07 (-0.21 0.06) ER = 3.91 | -0.06 (-0.18 0.06) ER = 4.05 |  |
| Skeptical CH | -0.13 (-0.42 0.14) ER = 3.37 | **-0.28 (-0.45 -0.12) ER = 749** | **-0.38 (-0.75 -0.05) ER = 35.59** | -0.08 (-0.33 0.17) ER = 2.21 ER01 = 3.41 |  |
| **PSP IQR** |  |  |  |  |  |
| Skeptical DK | 0.01 (-0.13 0.16) ER = 1.3 ER01 = 6.89 | 0.05 (-0.07 0.18) ER = 3.01 | -0.06 (-0.2 0.08) ER = 3.12 | -0.01 (-0.14 0.11) ER = 1.34 ER01 = 8.13 |  |
| Skeptical CH | **-0.29 (-0.68 0.04) ER = 12.1** | **-0.31 (-0.54 -0.1) ER = 170.43** | **-0.51 (-0.97 -0.11) ER = 54.56** | -0.12 (-0.45 0.19) ER = 2.77 ER01 = 2.72 |  |
| **MDQ Median** |  |  |  |  |  |
| Skeptical DK | -0.02 (-0.16 0.12) ER = 1.34 ER01 = 7.32 | 0.12 (0 0.24) ER = 18.8 | 0.07 (-0.06 0.2) ER = 4.22 | -0.03 (-0.14 0.09) ER = 1.84 ER01 = 8.02 |  |
| Skeptical CH | -0.07 (-0.37 0.2) ER = 1.9 ER01 = 3.15 | -0.19 (-0.38 -0.02) ER = 25.55 | -0.11 (-0.45 0.22) ER = 2.61 ER01 = 2.78 | -0.1 (-0.33 0.13) ER = 3.54 |  |
| **MDQ IQR** |  |  |  |  |  |
| Skeptical DK | 0.01 (-0.08 0.09) ER = 1.48 ER01 = 10.85 | -0.06 (-0.13 0.01) ER = 11.88 | -0.02 (-0.09 0.05) ER = 1.9 ER01 = 13.04 | 0.05 (-0.02 0.11) ER = 6.85 |  |
| Skeptical CH | 0.11 (-0.15 0.37) ER = 3.18 | 0.07 (-0.1 0.23) ER = 3.1 | 0.1 (-0.21 0.44) ER = 2.22 ER01 = 2.96 | 0.04 (-0.18 0.26) ER = 1.53 ER01 = 4.31 |  |
| **HRF Median** |  |  |  |  |  |
| Skeptical DK | 0 (-0.2 0.2) ER = 1.13 ER01 = 5.15 | -0.12 (-0.28 0.07) ER = 5.97 | 0.04 (-0.13 0.21) ER = 1.89 ER01 = 5.35 | -0.12 (-0.31 0.06) ER = 6.41 |  |
| Skeptical CH | 0 (-0.36 0.34) ER = 1.08 ER01 = 3.01 | -0.03 (-0.28 0.2) ER = 1.39 ER01 = 4.54 | 0.06 (-0.28 0.4) ER = 1.67 ER01 = 2.71 | -0.09 (-0.58 0.4) ER = 1.72 ER01 = 1.96 |  |
| **HRF IQR** |  |  |  |  |  |
| Skeptical DK | -0.03 (-0.12 0.04) ER = 3.18 | -0.08 (-0.14 -0.02) ER = 52.1 | 0.01 (-0.05 0.07) ER = 1.79 ER01 = 16.47 | -0.05 (-0.11 0.02) ER = 6.95 |  |
| Skeptical CH | -0.04 (-0.25 0.16) ER = 1.68 ER01 = 4.7 | 0.06 (-0.06 0.19) ER = 3.79 | -0.04 (-0.21 0.12) ER = 2.11 ER01 = 5.61 | -0.07 (-0.32 0.17) ER = 2.22 ER01 = 3.89 |  |
| **Rating scales** | **PANSS Total** | **PANSS**  **Negative** | **PANSS**  **Positive** | **PANSS Conversation** | **PANSS**  **Blunted affect** |
| **F1 Median** |  |  |  |  |  |
| Skeptical CH | 0.04 (-0.3 0.35) ER = 1.39 ER01 = 2.99 | **-0.21 (-0.4 -0.02) ER = 25.55** | 0.16 (-0.04 0.37) ER = 9.93 | -0.09 (-0.23 0.04) ER = 7.15 | **-0.17 (-0.35 0) ER = 19.34** |
| Skeptical JP | -0.25 (-0.78 0.31) ER = 3.31 | -0.18 (-0.78 0.41) ER = 2.22 ER01 = 1.56 | -0.11 (-0.7 0.47) ER = 1.67 ER01 = 1.55 |  |  |
| Skeptical GE | -0.04 (-0.36 0.27) ER = 1.47 ER01 = 3.35 | -0.1 (-0.33 0.13) ER = 3.19 | 0.03 (-0.28 0.32) ER = 1.41 ER01 = 3.28 | -0.09 (-0.22 0.05) ER = 5.99 | 0.15 (-0.11 0.47) ER = 4.64 |
| **F1 IQR** |  |  |  |  |  |
| Skeptical CH | 0.02 (-0.35 0.38) ER = 1.21 ER01 = 2.82 | **-0.21 (-0.43 0.01) ER = 17.07** | 0.16 (-0.09 0.41) ER = 6.05 | **-0.13 (-0.28 0.01) ER = 16.19** | **-0.18 (-0.38 0.01) ER = 16.6** |
| Skeptical JP | -0.24 (-0.58 0.11) ER = 7.32 | -0.22 (-0.61 0.16) ER = 5.26 | **-0.29 (-0.61 0.04) ER = 13.12** |  |  |
| Skeptical GE | 0.22 (-0.1 0.55) ER = 7.28 | -0.07 (-0.33 0.17) ER = 2.03 ER01 = 3.93 | **0.35 (0.08 0.64) ER = 44.8** | -0.09 (-0.26 0.07) ER = 4.99 | 0.01 (-0.34 0.35) ER = 1.19 ER01 = 3.5 |
| **F2 Median** |  |  |  |  |  |
| Skeptical CH | -0.29 (-0.68 0.09) ER = 8.43 | **-0.21 (-0.45 0.02) ER = 13.56** | 0.03 (-0.26 0.31) ER = 1.33 ER01 = 3.65 | **-0.17 (-0.35 0.01) ER = 14.79** | -0.11 (-0.36 0.12) ER = 3.67 |
| Skeptical JP | **-0.39 (-0.78 0.03) ER = 16.19** | -0.33 (-0.8 0.15) ER = 7.8 | **-0.37 (-0.78 0.05) ER = 12.61** |  |  |
| Skeptical GE | -0.22 (-0.53 0.06) ER = 8.46 | **-0.21 (-0.44 0) ER = 18.35** | -0.12 (-0.42 0.15) ER = 3.09 | 0.02 (-0.12 0.18) ER = 1.37 ER01 = 6.7 | -0.07 (-0.39 0.23) ER = 2 ER01 = 3.74 |
| **F2 IQR** |  |  |  |  |  |
| Skeptical CH | -0.26 (-0.66 0.13) ER = 6.23 | -0.09 (-0.36 0.19) ER = 2.39 ER01 = 3.12 | -0.14 (-0.45 0.14) ER = 3.71 | -0.03 (-0.24 0.18) ER = 1.28 ER01 = 4.61 | 0.01 (-0.23 0.24) ER = 1.23 ER01 = 4.28 |
| Skeptical JP | -0.03 (-0.58 0.54) ER = 1.14 ER01 = 1.77 | 0.02 (-0.57 0.6) ER = 1.13 ER01 = 1.76 | 0.02 (-0.58 0.6) ER = 1.11 ER01 = 1.67 |  |  |
| Skeptical GE | -0.01 (-0.22 0.2) ER = 1.12 ER01 = 5.1 | -0.11 (-0.27 0.05) ER = 6.82 | **-0.15 (-0.35 0.02) ER = 13.35** | -0.02 (-0.12 0.07) ER = 1.62 ER01 = 11.03 | 0.05 (-0.16 0.29) ER = 1.97 ER01 = 4.88 |
| **F3 Median** |  |  |  |  |  |
| Skeptical CH | -0.2 (-0.63 0.24) ER = 3.68 | 0.1 (-0.18 0.39) ER = 2.7 ER01 = 3.05 | -0.06 (-0.38 0.25) ER = 1.67 ER01 = 3.18 | 0.02 (-0.19 0.22) ER = 1.33 ER01 = 5.26 | 0.12 (-0.13 0.37) ER = 3.8 |
| Skeptical JP | **-0.44 (-0.78 -0.07) ER = 31.26** | **-0.4 (-0.8 0.05) ER = 13.78** | **-0.37 (-0.78 0.05) ER = 13.71** |  |  |
| Skeptical GE | -0.04 (-0.3 0.22) ER = 1.51 ER01 = 3.99 | **-0.15 (-0.35 0.03) ER = 9.68** | -0.07 (-0.31 0.18) ER = 2.17 ER01 = 3.72 | 0.01 (-0.13 0.15) ER = 1.13 ER01 = 7.49 | 0.02 (-0.27 0.31) ER = 1.3 ER01 = 3.88 |
| **F3 IQR** |  |  |  |  |  |
| Skeptical CH | -0.18 (-0.62 0.26) ER = 3.03 | 0.09 (-0.22 0.38) ER = 2.15 ER01 = 2.94 | **-0.38 (-0.68 -0.08) ER = 47.39** | 0.16 (-0.05 0.37) ER = 8.87 | 0.03 (-0.23 0.3) ER = 1.29 ER01 = 3.81 |
| Skeptical JP | -0.17 (-0.56 0.22) ER = 3.59 | -0.14 (-0.59 0.29) ER = 2.45 ER01 = 2 | -0.16 (-0.57 0.27) ER = 2.94 ER01 = 1.93 |  |  |
| Skeptical GE | -0.06 (-0.3 0.18) ER = 2 ER01 = 4.06 | -0.04 (-0.22 0.15) ER = 1.74 ER01 = 5.17 | -0.1 (-0.32 0.13) ER = 3.33 | 0.08 (-0.04 0.21) ER = 6.87 | 0.04 (-0.2 0.29) ER = 1.61 ER01 = 4.24 |
| **F4 Median** |  |  |  |  |  |
| Skeptical CH | -0.12 (-0.49 0.25) ER = 2.41 ER01 = 2.52 | -0.05 (-0.28 0.18) ER = 1.81 ER01 = 4.48 | 0 (-0.26 0.27) ER = 1.02 ER01 = 3.82 | -0.01 (-0.21 0.17) ER = 1.09 ER01 = 5.27 | -0.05 (-0.27 0.15) ER = 1.91 ER01 = 4.87 |
| Skeptical JP | -0.28 (-0.69 0.15) ER = 6.97 | -0.2 (-0.66 0.27) ER = 3.24 | -0.28 (-0.68 0.14) ER = 6.75 | 0.06 (-0.09 0.22) ER = 2.89 ER01 = 5.54 | -0.05 (-0.38 0.26) ER = 1.5 ER01 = 3.49 |
| Skeptical GE | -0.05 (-0.34 0.23) ER = 1.58 ER01 = 3.43 | -0.11 (-0.33 0.09) ER = 4.32 | -0.01 (-0.29 0.26) ER = 1.13 ER01 = 3.96 |  |  |
| **F4 IQR** |  |  |  |  |  |
| Skeptical CH | **0.33 (-0.01 0.66) ER = 17.75** | 0.13 (-0.09 0.35) ER = 5.45 | 0 (-0.27 0.26) ER = 1.04 ER01 = 3.97 | 0.09 (-0.06 0.25) ER = 5.63 | 0.11 (-0.08 0.31) ER = 4.93 |
| Skeptical JP | 0.26 (-0.25 0.76) ER = 4.35 | 0.28 (-0.27 0.81) ER = 4.42 | 0.25 (-0.25 0.75) ER = 4.17 |  |  |
| Skeptical GE | -0.14 (-0.34 0.05) ER = 8.65 | -0.1 (-0.25 0.04) ER = 6.91 | **-0.15 (-0.35 0.03) ER = 10.47** | -0.01 (-0.09 0.07) ER = 1.57 ER01 = 12.77 | -0.06 (-0.24 0.1) ER = 2.76 ER01 = 5.83 |
| **F5 Median** |  |  |  |  |  |
| Skeptical CH | -0.31 (-0.75 0.13) ER = 6.97 | -0.15 (-0.45 0.14) ER = 4.34 | -0.02 (-0.37 0.31) ER = 1.17 ER01 = 3.17 | -0.09 (-0.32 0.13) ER = 2.87 ER01 = 3.63 | -0.14 (-0.43 0.14) ER = 3.97 |
| Skeptical JP | -0.08 (-0.45 0.3) ER = 1.88 ER01 = 2.64 | -0.03 (-0.44 0.38) ER = 1.27 ER01 = 2.54 | 0.08 (-0.31 0.47) ER = 1.79 ER01 = 2.51 |  |  |
| Skeptical GE | 0.11 (-0.06 0.3) ER = 6.13 | 0.02 (-0.12 0.15) ER = 1.41 ER01 = 7.76 | **0.14 (-0.02 0.3) ER = 12.42** | 0.03 (-0.05 0.12) ER = 2.34 ER01 = 11.46 | 0.04 (-0.15 0.23) ER = 1.87 ER01 = 5.69 |
| **F5 IQR** |  |  |  |  |  |
| Skeptical CH | -0.09 (-0.48 0.3) ER = 1.81 ER01 = 2.33 | -0.06 (-0.3 0.18) ER = 1.92 ER01 = 3.85 | -0.19 (-0.47 0.09) ER = 6.13 | 0.1 (-0.08 0.28) ER = 5.7 | -0.06 (-0.29 0.15) ER = 2.11 ER01 = 4.17 |
| Skeptical JP | **0.28 (-0.04 0.59) ER = 12.61** | 0.24 (-0.14 0.62) ER = 6.38 | 0.19 (-0.18 0.55) ER = 4.55 |  |  |
| Skeptical GE | **0.24 (0.01 0.48) ER = 20.82** | **0.3 (0.13 0.51) ER = 599** | 0.03 (-0.19 0.26) ER = 1.39 ER01 = 4.64 | **0.1 (-0.02 0.22) ER = 10.88** | 0.06 (-0.21 0.34) ER = 1.73 ER01 = 4 |
| **H1H2 Median** |  |  |  |  |  |
| Skeptical CH | **-0.45 (-0.72 -0.18) ER = 165.67** | 0 (-0.19 0.2) ER = 1.04 ER01 = 5.21 | **-0.22 (-0.42 -0.02) ER = 27.17** | -0.04 (-0.17 0.09) ER = 2.28 ER01 = 6.78 | 0 (-0.17 0.19) ER = 0.99 ER01 = 5.76 |
| Skeptical JP | 0.02 (-0.62 0.66) ER = 1.08 ER01 = 1.56 | 0.13 (-0.53 0.79) ER = 1.72 ER01 = 1.36 | -0.02 (-0.71 0.66) ER = 1.12 ER01 = 1.48 |  |  |
| Skeptical GE | -0.05 (-0.39 0.28) ER = 1.53 ER01 = 2.95 | 0.06 (-0.2 0.33) ER = 1.95 ER01 = 3.46 | **-0.36 (-0.71 -0.05) ER = 42.8** | -0.02 (-0.21 0.15) ER = 1.28 ER01 = 5.86 | 0 (-0.32 0.31) ER = 0.99 ER01 = 3.86 |
| **H1H2 IQR** |  |  |  |  |  |
| Skeptical CH | 0.11 (-0.18 0.4) ER = 2.95 ER01 = 2.74 | **-0.14 (-0.31 0.03) ER = 11.55** | 0.15 (-0.04 0.33) ER = 9.47 | -0.06 (-0.18 0.06) ER = 3.87 | -0.1 (-0.27 0.05) ER = 5.98 |
| Skeptical JP | -0.23 (-0.68 0.22) ER = 4.31 | -0.14 (-0.63 0.36) ER = 2.09 ER01 = 1.74 | -0.11 (-0.59 0.36) ER = 1.98 ER01 = 1.89 |  |  |
| Skeptical GE | -0.03 (-0.25 0.19) ER = 1.54 ER01 = 4.33 | -0.07 (-0.24 0.11) ER = 2.98 ER01 = 4.71 | 0.09 (-0.12 0.31) ER = 2.89 ER01 = 3.74 | 0.05 (-0.06 0.18) ER = 2.93 ER01 = 6.98 | -0.08 (-0.34 0.12) ER = 2.85 ER01 = 4.3 |
| **QOQ Median** |  |  |  |  |  |
| MA priors |  |  |  |  |  |
| Skeptical CH | -0.33 (-0.82 0.18) ER = 6.03 | -0.04 (-0.4 0.32) ER = 1.35 ER01 = 2.58 | **-0.3 (-0.68 0.08) ER = 10.15** | -0.1 (-0.38 0.17) ER = 2.63 ER01 = 3.14 | -0.08 (-0.38 0.24) ER = 1.99 ER01 = 3.19 |
| Skeptical JP | -0.16 (-0.71 0.4) ER = 2.23 ER01 = 1.73 | -0.15 (-0.74 0.45) ER = 1.97 ER01 = 1.58 | -0.22 (-0.79 0.37) ER = 2.93 ER01 = 1.4 |  |  |
| Skeptical GE | 0.1 (-0.27 0.49) ER = 2.06 ER01 = 2.4 | 0.21 (-0.07 0.52) ER = 7.88 | -0.2 (-0.59 0.17) ER = 4.61 | 0.07 (-0.16 0.27) ER = 2.55 ER01 = 3.96 | -0.04 (-0.46 0.35) ER = 1.21 ER01 = 2.79 |
| **QOQ IQR** |  |  |  |  |  |
| Skeptical CH | -0.05 (-0.47 0.36) ER = 1.39 ER01 = 2.39 | -0.17 (-0.43 0.1) ER = 6.15 | 0.01 (-0.29 0.31) ER = 1.12 ER01 = 3.37 | **-0.17 (-0.34 0.02) ER = 12.82** | -0.12 (-0.34 0.1) ER = 4.45 |
| Skeptical JP | 0.12 (-0.31 0.56) ER = 2.13 ER01 = 2.1 | 0.16 (-0.33 0.63) ER = 2.56 ER01 = 1.75 | 0.06 (-0.41 0.53) ER = 1.39 ER01 = 2.15 |  |  |
| Skeptical GE | -0.1 (-0.3 0.1) ER = 3.64 | -0.05 (-0.2 0.1) ER = 2.32 ER01 = 5.96 | **-0.17 (-0.39 0.02) ER = 14.08** | -0.04 (-0.13 0.05) ER = 3.23 | **-0.16 (-0.42 0.03) ER = 9.53** |
| **NAQ Median** |  |  |  |  |  |
| Skeptical CH | -0.39 (-0.89 0.13) ER = 7.78 | 0.01 (-0.35 0.38) ER = 1.05 ER01 = 2.73 | **-0.35 (-0.74 0.03) ER = 14.54** | -0.06 (-0.35 0.22) ER = 1.71 ER01 = 3.48 | -0.02 (-0.34 0.31) ER = 1.23 ER01 = 3.14 |
| Skeptical JP | -0.32 (-0.84 0.19) ER = 5.73 | -0.3 (-0.82 0.23) ER = 5.01 | -0.4 (-0.91 0.15) ER = 9.1 |  |  |
| Skeptical GE | 0.07 (-0.28 0.41) ER = 1.73 ER01 = 2.86 | 0.13 (-0.15 0.43) ER = 3.68 | -0.16 (-0.51 0.17) ER = 3.84 | 0.08 (-0.13 0.26) ER = 3.42 | -0.05 (-0.48 0.33) ER = 1.34 ER01 = 2.73 |
| **NAQ IQR** |  |  |  |  |  |
| Skeptical CH | -0.24 (-0.72 0.24) ER = 4 | -0.05 (-0.38 0.26) ER = 1.5 ER01 = 3.04 | -0.23 (-0.58 0.12) ER = 6.48 | -0.13 (-0.38 0.14) ER = 3.69 | -0.06 (-0.34 0.23) ER = 1.77 ER01 = 3.33 |
| Skeptical JP | -0.31 (-0.85 0.26) ER = 4.54 | -0.21 (-0.79 0.39) ER = 2.59 ER01 = 1.39 | -0.29 (-0.86 0.29) ER = 3.83 |  |  |
| Skeptical GE | -0.05 (-0.32 0.22) ER = 1.58 ER01 = 3.74 | 0.04 (-0.16 0.24) ER = 1.59 ER01 = 4.89 | **-0.2 (-0.46 0.03) ER = 11.45** | 0.05 (-0.08 0.18) ER = 3.07 | -0.05 (-0.37 0.23) ER = 1.52 ER01 = 3.92 |
| **HNR Median** |  |  |  |  |  |
| Skeptical CH | -0.11 (-0.53 0.3) ER = 2.2 ER01 = 2.06 | 0.18 (-0.08 0.44) ER = 7.13 | **-0.29 (-0.56 -0.02) ER = 23.59** | **0.17 (-0.01 0.35) ER = 16.34** | **0.21 (0 0.43) ER = 20.2** |
| Skeptical JP | 0.05 (-0.46 0.54) ER = 1.32 ER01 = 1.95 | -0.02 (-0.54 0.5) ER = 1.08 ER01 = 1.99 | -0.11 (-0.61 0.39) ER = 1.92 ER01 = 1.78 |  |  |
| Skeptical GE | -0.03 (-0.26 0.2) ER = 1.31 ER01 = 4.41 | 0.04 (-0.13 0.22) ER = 1.81 ER01 = 5.36 | -0.17 (-0.39 0.05) ER = 7.96 | -0.04 (-0.18 0.08) ER = 2.14 ER01 = 7.48 | -0.12 (-0.39 0.11) ER = 4.34 |
| **HNR IQR** |  |  |  |  |  |
| Skeptical CH | -0.08 (-0.42 0.26) ER = 1.88 ER01 = 2.79 | 0.2 (0.01 0.39) ER = 24.1 | -0.2 (-0.43 0.02) ER = 13.56 | 0.1 (-0.04 0.25) ER = 6.89 | **0.14 (-0.03 0.31) ER = 11.5** |
| Skeptical JP | -0.21 (-0.8 0.38) ER = 2.77 ER01 = 1.39 | -0.17 (-0.78 0.46) ER = 2.04 ER01 = 1.39 | -0.29 (-0.89 0.32) ER = 3.67 |  |  |
| Skeptical GE | **0.28 (-0.05 0.59) ER = 12.27** | **0.25 (0.01 0.51) ER = 23.1** | -0.03 (-0.33 0.27) ER = 1.23 ER01 = 3.36 | 0.06 (-0.12 0.25) ER = 2.35 ER01 = 4.57 | 0.02 (-0.35 0.38) ER = 1.21 ER01 = 3.19 |
| **PSP Median** |  |  |  |  |  |
| Skeptical CH | -0.11 (-0.57 0.33) ER = 1.92 ER01 = 1.94 | -0.08 (-0.38 0.2) ER = 2.12 ER01 = 3.01 | -0.1 (-0.42 0.21) ER = 2.29 ER01 = 2.82 | 0.08 (-0.14 0.28) ER = 2.9 ER01 = 3.68 | -0.13 (-0.43 0.14) ER = 3.17 |
| Skeptical JP | 0.05 (-0.39 0.47) ER = 1.45 ER01 = 2.31 | 0.04 (-0.42 0.5) ER = 1.33 ER01 = 2.13 | 0.19 (-0.24 0.61) ER = 3.64 |  |  |
| Skeptical GE | -0.05 (-0.37 0.28) ER = 1.54 ER01 = 3.12 | -0.08 (-0.34 0.16) ER = 2.31 ER01 = 3.61 | 0.15 (-0.15 0.48) ER = 3.89 | -0.01 (-0.19 0.22) ER = 1.37 ER01 = 4.57 | **0.32 (-0.02 0.75) ER = 14.23** |
| **PSP IQR** |  |  |  |  |  |
| Skeptical CH | -0.01 (-0.54 0.49) ER = 1.1 ER01 = 2 | -0.14 (-0.51 0.21) ER = 2.9 ER01 = 2.32 | 0.06 (-0.33 0.44) ER = 1.52 ER01 = 2.51 | 0.03 (-0.24 0.29) ER = 1.4 ER01 = 3.63 | -0.15 (-0.5 0.16) ER = 3.16 |
| Skeptical JP | 0.04 (-0.34 0.41) ER = 1.37 ER01 = 2.5 | 0.03 (-0.39 0.43) ER = 1.28 ER01 = 2.42 |  |  |  |
| Skeptical GE | -0.15 (-0.4 0.09) ER = 5.89 | -0.12 (-0.31 0.07) ER = 5.8 | 0.01 (-0.21 0.23) ER = 1.17 ER01 = 4.76 | 0.1 (-0.29 0.46) ER = 2.15 ER01 = 2.37 | -0.08 (-0.2 0.05) ER = 6.46 |
| **MDQ Median** |  |  |  |  |  |
| Skeptical CH | **-0.52 (-0.9 -0.13) ER = 60.22** | -0.06 (-0.34 0.23) ER = 1.69 ER01 = 3.4 | **-0.4 (-0.69 -0.12) ER = 71.29** | 0.06 (-0.17 0.26) ER = 2.18 ER01 = 3.96 | -0.07 (-0.33 0.18) ER = 2.07 ER01 = 3.79 |
| Skeptical JP | **-0.39 (-0.85 0.1) ER = 10.61** | -0.35 (-0.83 0.16) ER = 7.05 | -0.36 (-0.82 0.11) ER = 9.03 | -0.07 (-0.17 0.04) ER = 6.07 | 0.11 (-0.15 0.42) ER = 3.03 |
| Skeptical GE | **-0.2 (-0.46 0.05) ER = 10.01** | **-0.19 (-0.38 -0.01) ER = 22.17** | -0.1 (-0.35 0.14) ER = 3.05 |  |  |
| **MDQ IQR** |  |  |  |  |  |
| Skeptical CH | 0.33 (-0.1 0.75) ER = 9.17 | -0.03 (-0.32 0.25) ER = 1.34 ER01 = 3.57 | **0.38 (0.07 0.69) ER = 39.27** | -0.05 (-0.25 0.16) ER = 2.04 ER01 = 4.38 | 0.09 (-0.17 0.36) ER = 2.43 ER01 = 3.15 |
| Skeptical JP | 0.25 (-0.29 0.76) ER = 3.73 | 0.17 (-0.4 0.72) ER = 2.4 ER01 = 1.5 | 0.3 (-0.25 0.81) ER = 5.07 |  |  |
| Skeptical GE | 0.03 (-0.18 0.25) ER = 1.48 ER01 = 4.42 | 0.05 (-0.11 0.22) ER = 2.25 ER01 = 5.05 | 0.07 (-0.14 0.27) ER = 2.54 ER01 = 4.32 | 0.02 (-0.08 0.12) ER = 1.48 ER01 = 10.37 | -0.03 (-0.24 0.16) ER = 1.53 ER01 = 5.97 |
| Skeptical GE |  |  |  |  |  |
| **HRF Median** |  |  |  |  |  |
| Skeptical CH | 0.37 (-0.13 0.85) ER = 8.88 | 0 (-0.35 0.33) ER = 0.97 ER01 = 2.98 | 0.28 (-0.08 0.62) ER = 9.62 | -0.05 (-0.31 0.21) ER = 1.76 ER01 = 3.53 | -0.01 (-0.33 0.31) ER = 1.06 ER01 = 3.26 |
| Skeptical JP | 0.38 (-0.29 1.02) ER = 4.78 | 0.29 (-0.36 0.97) ER = 3.43 | 0.47 (-0.21 1.13) ER = 7 |  |  |
| Skeptical GE | **0.34 (0.01 0.69) ER = 20.9** | 0.14 (-0.11 0.4) ER = 4.92 | **0.53 (0.21 0.9) ER = 499** | 0.08 (-0.05 0.22) ER = 5.49 | -0.13 (-0.47 0.17) ER = 3.07 |
| **HRF IQR** |  |  |  |  |  |
| Skeptical CH | 0.16 (-0.26 0.56) ER = 2.82 ER01 = 2.05 | -0.07 (-0.32 0.19) ER = 1.91 ER01 = 3.33 | 0.04 (-0.26 0.32) ER = 1.51 ER01 = 3.64 | 0.03 (-0.15 0.21) ER = 1.54 ER01 = 5.45 | -0.08 (-0.3 0.14) ER = 2.55 ER01 = 4.22 |
| Skeptical JP | 0.06 (-0.46 0.55) ER = 1.42 ER01 = 1.98 | 0 (-0.56 0.52) ER = 1.02 ER01 = 2.11 | 0.24 (-0.26 0.71) ER = 3.91 |  |  |
| Skeptical GE | -0.12 (-0.42 0.17) ER = 3.09 | **-0.23 (-0.47 -0.01) ER = 23** | **0.28 (0 0.58) ER = 19.27** | -0.07 (-0.22 0.08) ER = 3.95 | -0.16 (-0.52 0.15) ER = 4.1 |

**S6 – Analysis on Medication, Duration of Illness and Personal and Social Performance Scale (PSP)**

**S6.1 Effect of Medication**

The antipsychotic medications were divided into two categories based on their mechanism of action, that is high versus low dopamine occupancy of D2 receptor (D2R).

Atypical antipsychotics (AAPs) are weak D2 receptor antagonists, and they involve other receptor targets (e.g regulating dopamine and other neurotransmitters) besides D2 antagonism. AAPs never show a high D2R occupancy (above 70-80%) and this is why they cause reduced side effects such as parkinsonism and tardive dyskinesia (TD), apathy and anhedonia^34,35^.

In contrast, the optimal D2 receptor occupancy in the striatum for typical antipsychotics (TAPs) medication is very high (between 70 to 80%), and this explains why extrapyramidal symptoms (EPS) occur frequently, as well as other side effects such as parkinsonism, hyperprolactinemia, apathy and anhedonia, which are all related to the block of D2 receptors activity.

Patients were thus divided into two categories based on the different mechanisms of action, that is patients with (1) low D2R occupancy, i.e., Clozapine, Olanzapine, Paliperidone, Quetiapine and (2) high D2R occupancy, i.e., Aripiprazole, Amisulpride, Risperidone, Ziprasidone, Sertindole. Aripiprazole is generally categorized as a D2R antagonist because it could block completely the effects of either dopamine or quinpirole in different systems expressing the D2 receptor, although it also has agonistic effects (see Shapiro et al., 2003^43^). Antipsychotic drug dosages were converted into chlorpromazine equivalents (CPZ) to assess the effect of dosage across drugs (see Leucht et al., 2014). A summary table of medication data are shown in **Table S6_A**.

We used Bayesian multilevel regression models with each acoustic feature as outcome and CPZ equivalent dosage as predictor. We separately assessed the effect of CPZ dosage for each language, and modeled varying effects of participants, i.e., intercepts and slopes, separately for each group and language. For each acoustic feature we built a model with weakly informative priors, that is expectations of no effect of CPZ dosage.

**Table S6_A.** Frequency of the medication category (high vs. low D2R) for patients in the different corpora (Danish, Chinese, German)

|  | Danish corpus | | Chinese corpus | | German corpus | |
| --- | --- | --- | --- | --- | --- | --- |
|  | High D2R | Low D2R | High D2R | Low D2R | High D2R | Low D2R |
| Chlorpromazine equivalent (mg), mean (SD) | 41 | 44 | 11 | 18 | 12 | 7 |
| Antipsychotic medication, n | 441.7 (262.8) | 689.44 (445.9) | 399.1 (268.5) | 722.2 (393.7) | 620.6 (321.6) | 808.6 (641.7) |
| Aripiprazole | 37 |  | 1 |  | 4 |  |
| Amisulpride |  |  | 3 |  | 1 |  |
| Risperidone | 3 |  | 6 |  | 5 |  |
| Ziprasidone |  |  | 1 |  |  |  |
| Sertindole | 1 |  |  |  | 2 |  |
| Clozapine |  | 4 |  | 5 |  | 5 |
| Olanzapine |  | 6 |  | 6 |  | 2 |
| Paliperidone |  | 7 |  | 3 |  |  |
| Quietapine |  | 27 |  | 4 |  |  |

Fig. S6_A. Molecular targets of atypical antipsychotic medications AAPs.

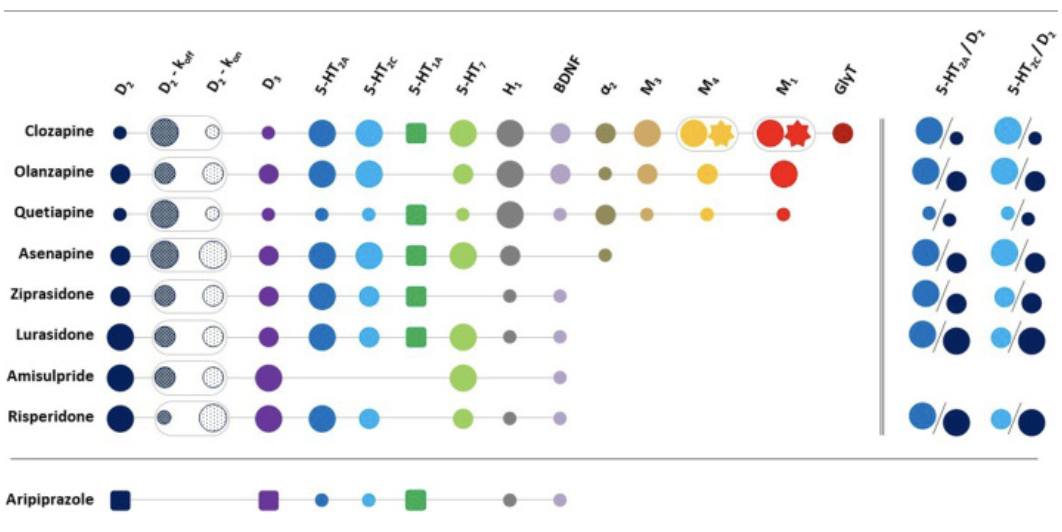

**Note:** List of the most relevant targets involved in the mechanism of action of AAPs based on receptor occupancy. Values are reported as high (●), medium(● )and low( ●).●, ■ and represent receptor antagonism, partial agonism and positive allosterism, respectively. and represent koff and kon values for the D2 receptor and ● represents BDNF production, while represents positive allosterism by the clozapine metabolite, norclozapine, at M1 and M4 receptors. Aripiprazole is shown at the bottom for its different mechanism of action. Finally, the 5-HT2A/D2 and 5-HT2C/D2 receptor affinity ratios are included on the right. Clozapine covers a wide-range of molecular targets among all AAPs, while risperidone and amisulpride are mostly limited to just a few, and this might explain clozapine's superiority among AAPs.

Figure taken from Aringhieri et al. (2018)^35^ , RightsLink licence number 5233570834080.

**Table S6_B.** Comparison between groups using different antipsychotic medication (high D2R vs. low D2R), and effect of CPZ dosage, on each acoustic feature

| **Acoustic features** | **Group** |  |  | **CPZ dosage** |
| --- | --- | --- | --- | --- |
| **Pitch Median** | Low D2R – HC | High D2R – HC | High D2R – Low D2R |  |
| Skeptical DK | -0.02 (-0.29 0.24) ER = 1.14 ER01 = 2.97 | 0.08 (-0.2 0.36) ER = 2.26 ER01 = 2.6 | 0.1 (-0.24 0.44) ER = 2.03 ER01 = 3 | 0.01 (-0.04 0.06) ER = 1.49 ER01 = 1682.85 |
| Skeptical CH | **0.6 (0.21 0.97) ER = 157.73** | 0.15 (-0.32 0.61) ER = 2.53 ER01 = 1.54 | -0.44 (-0.98 0.12) ER = 9.62 | -0.02 (-0.09 0.05) ER = 2.18 ER01 = 1025.11 |
| Skeptical GE | -0.32 (-0.75 0.12) ER = 8.12 | -0.21 (-0.56 0.15) ER = 4.78 | 0.12 (-0.42 0.66) ER = 1.86 ER01 = 2.04 | **-0.05 (-0.1 -0.01) ER = 50.55** |
| **Pitch IQR** | Low D2R – HC | High D2R – HC | High D2R – Low D2R |  |
| Skeptical DK | -0.04 (-0.21 0.14) ER = 1.78 ER01 = 4.39 | **-0.32 (-0.5 -0.14) ER = 499** | **-0.28 (-0.49 -0.07) ER = 69.92** | -0.01 (-0.05 0.02) ER = 2.85 ER01 = 1986.71 |
| Skeptical CH | -0.2 (-0.49 0.1) ER = 6.01 | -0.23 (-0.58 0.12) ER = 6.24 | -0.04 (-0.44 0.37) ER = 1.28 ER01 = 2.85 | 0.01 (-0.05 0.07) ER = 1.43 ER01 = 1305.26 |
| Skeptical GE | **-0.35 (-0.59 -0.1) ER = 89.91** | **-0.47 (-0.68 -0.27) ER = 9999** | -0.13 (-0.39 0.14) ER = 3.97 | -0.01 (-0.04 0.03) ER = 1.57 ER01 = 2450.85 |
| **Speech Rate** | Low D2R – HC | High D2R – HC | High D2R – Low D2R |  |
| Skeptical DK | **-0.44 (-0.65 -0.22) ER = 2499** | **-0.19 (-0.41 0.03) ER = 12.35** | **0.24 (-0.02 0.52) ER = 13.43** | 0.01 (-0.03 0.05) ER = 1.54 ER01 = 2089.11 |
| Skeptical CH | -0.03 (-0.38 0.32) ER = 1.29 ER01 = 2.4 | 0.13 (-0.25 0.53) ER = 2.44 ER01 = 1.75 | 0.16 (-0.31 0.62) ER = 2.56 ER01 = 2.09 | **-0.05 (-0.11 0.01) ER = 13.88** |
| Skeptical GE | 0.24 (-0.19 0.67) ER = 4.77 | -0.06 (-0.41 0.3) ER = 1.58 ER01 = 2.26 | -0.3 (-0.83 0.24) ER = 4.87 | 0.03 (-0.01 0.07) ER = 9.33 |
| **Speech Percentage** | Low D2R – HC | High D2R – HC | High D2R – Low D2R |  |
| MA_Priors_ | -0.66 (+0.29 -0.29) | -0.76 (+0.27 -0.27) | -0.08 (+0.31 -0.31) |  |
| Skeptical DK | -0.07 (-0.31 0.17) ER = 2.23 ER01 = 3.26 | 0.17 (-0.07 0.41) ER = 7 | 0.24 (-0.06 0.55) ER = 9.42 | 0.02 (-0.02 0.06) ER = 2.9 ER01 = 1643.61 |
| Skeptical CH | **0.34 (-0.01 0.69) ER = 16.57** | 0.29 (-0.12 0.7) ER = 7.38 | -0.05 (-0.52 0.44) ER = 1.33 ER01 = 2.28 | **-0.05 (-0.11 0) ER = 15.69** |
| Skeptical GE | **0.38 (-0.08 0.83) ER = 10.86** | 0.28 (-0.11 0.66) ER = 7.67 | -0.1 (-0.66 0.47) ER = 1.56 ER01 = 1.99 | **0.04 (0 0.08) ER = 21.88** |
| Informed DK | -0.23 (-0.44 -0.02) ER = 25.39 | -0.07 (-0.29 0.16) ER = 2.24 ER01 = 94.41 | 0.24 (-0.06 0.55) ER = 9.42 |  |
| Informed CH | -0.1 (-0.42 0.22) ER = 2.34 ER01 = 16.6 | **-0.29 (-0.65 0.05) ER = 11.21** | -0.19 (-0.61 0.23) ER = 3.35 |  |
| Informed GE | **0.38 (-0.08 0.83) ER = 10.86** | -0.28 (-0.65 0.07) ER = 9.71 | -0.03 (-0.54 0.46) ER = 1.17 ER01 = 1.4 |  |
| **Pause Number Min** |  |  |  |  |
| Skeptical DK | -0.09 (-0.28 0.1) ER = 3.75 | **-0.2 (-0.4 0) ER = 20.28** | -0.11 (-0.36 0.14) ER = 3.3 | 0.02 (-0.01 0.05) ER = 9.43 |
| Skeptical CH | **-0.25 (-0.55 0.05) ER = 11.02** | 0.11 (-0.24 0.47) ER = 2.23 ER01 = 2.01 | **0.35 (-0.08 0.78) ER = 10.11** | -0.04 (-0.09 0.01) ER = 8.51 |
| Skeptical GE | 0.22 (-0.15 0.59) ER = 5.2 | -0.07 (-0.37 0.25) ER = 1.77 ER01 = 2.6 | -0.29 (-0.74 0.17) ER = 5.8 | 0.01 (-0.03 0.04) ER = 1.52 ER01 = 2344.86 |
| **Pause Duration** |  |  |  |  |
| MA_Priors | 0.17 (+0.28 -0.28) | 0.73 (+0.27 -0.27) | 0.59 (+0.32 -0.32) |  |
| Skeptical DK | **0.22 (0.01 0.43) ER = 22.98** | 0.16 (-0.06 0.38) ER = 7.55 | -0.06 (-0.35 0.23) ER = 1.75 ER01 = 3.65 | -0.01 (-0.04 0.02) ER = 2.8 ER01 = 2178.4 |
| Skeptical CH | 0.01 (-0.29 0.31) ER = 1.05 ER01 = 2.72 | -0.26 (-0.61 0.11) ER = 7.2 | -0.26 (-0.69 0.17) ER = 5.85 | **0.09 (0.03 0.14) ER = 249** |
| Skeptical GE | -0.23 (-0.62 0.15) ER = 5.21 | 0.01 (-0.32 0.33) ER = 1.11 ER01 = 2.61 | 0.24 (-0.24 0.72) ER = 4.18 | 0 (-0.04 0.03) ER = 1.18 ER01 = 2332.77 |
| Informed DK | **0.23 (0.03 0.43) ER = 31.36** | **0.29 (0.08 0.5) ER = 85.21** | -0.06 (-0.35 0.23) ER = 1.75 ER01 = 3.57 |  |
| Informed CH | 0.11 (-0.17 0.39) ER = 0.34 ER01 = 1.66 | 0.18 (-0.13 0.52) ER = 4.77 | 0.07 (-0.32 0.48) ER = 1.61 ER01 = 4 |  |
| Informed GE | -0.04 (-0.37 0.29) ER = 1.47 ER01 = 1.7 | **0.3 (0.01 0.61) ER = 22.75** | **0.35 (-0.07 0.76) ER = 10.71** |  |
| **Number of utterance** |  |  |  |  |
| MA_Priors |  |  |  |  |
| Skeptical DK | -0.12 (-0.31 0.07) ER = 6.09 | **-0.22 (-0.42 -0.02) ER = 30.35** | -0.1 (-0.35 0.15) ER = 2.84 ER01 = 3.65 | 0.02 (-0.01 0.06) ER = 8.05 |
| Skeptical CH | -0.22 (-0.53 0.09) ER = 7.16 | 0.09 (-0.29 0.46) ER = 1.85 ER01 = 2.06 | 0.3 (-0.16 0.75) ER = 6.26 | -0.04 (-0.09 0.02) ER = 7.42 |
| Skeptical GE | 0.19 (-0.19 0.55) ER = 4.18 | -0.09 (-0.4 0.21) ER = 2.29 ER01 = 2.29 | -0.28 (-0.72 0.18) ER = 5.76 | 0 (-0.03 0.04) ER = 1.11 ER01 = 2453.86 |
| **Utterance Duration** |  |  |  |  |
| MA_Priors_High vs HC | -0.42 (+0.29 -0.29) | -0.25 (+0.26 -0.26) | 0.19 (+0.32 -0.32) |  |
| Skeptical DK | 0.07 (-0.14 0.29) ER = 2.53 ER01 = 3.33 | **0.37 (0.15 0.59) ER = 383.62** | **0.3 (0.02 0.57) ER = 26.7** | 0.01 (-0.03 0.04) ER = 1.45 ER01 = 2114.64 |
| Skeptical CH | **0.5 (0.17 0.83) ER = 102.09** | 0.18 (-0.23 0.59) ER = 3.27 | -0.32 (-0.8 0.18) ER = 5.94 | 0 (-0.02 0.02) ER = 1 ER01 = 4282.24 |
| Skeptical GE | 0.28 (-0.22 0.78) ER = 4.87 | **0.54 (0.11 0.94) ER = 42.48** | 0.25 (-0.37 0.87) ER = 2.93 ER01 = 1.44 | **0.05 (0.01 0.08) ER = 39.32** |
| Informed DK | -0.04 (-0.23 0.16) ER = 1.74 ER01 = 6.06 | 0.24 (0.04 0.44) ER = 37.61 | **0.3 (0.02 0.57) ER = 26.7** |  |
| Informed CH | 0.18 (-0.14 0.49) ER = 0.21 ER01 = 2.67 | -0.06 (-0.39 0.26) ER = 1.57 ER01 = 2.07 | -0.24 (-0.67 0.19) ER = 4.69 |  |
| Informed GE | 0.28 (-0.22 0.78) ER = 4.87 | 0.14 (-0.21 0.47) ER = 3.08 | 0.27 (-0.22 0.78) ER = 4.38 |  |
| **Total Number of words** |  |  |  |  |
| MA_Priors_High vs HC | -0.45 (+0.29 -0.29) | -1.18 (+0.28 -0.28) | -0.76 (+0.33 -0.33) |  |
| Skeptical DK | **-0.3 (-0.51 -0.08) ER = 80.3** | **-0.19 (-0.41 0.03) ER = 11.47** | 0.11 (-0.14 0.37) ER = 3.13 | **-0.04 (-0.07 0) ER = 25.67** |
| Skeptical CH | -0.06 (-0.37 0.26) ER = 1.67 ER01 = 2.59 | -0.23 (-0.6 0.14) ER = 5.49 | -0.17 (-0.6 0.26) ER = 2.83 ER01 = 2.21 | 0.01 (-0.05 0.07) ER = 1.59 ER01 = 1282.31 |
| Skeptical GE | -0.09 (-0.47 0.3) ER = 1.87 ER01 = 1.97 | -0.21 (-0.54 0.11) ER = 6.28 | -0.12 (-0.58 0.34) ER = 2.04 ER01 = 2.13 | 0 (-0.03 0.04) ER = 1.18 ER01 = 2340.37 |
| Informed DK | **-0.4 (-0.6 -0.2) ER = 2499** | **-0.42 (-0.63 -0.21) ER = 3332.33** | 0.11 (-0.14 0.37) ER = 3.13 |  |
| Informed CH | **-0.27 (-0.56 0.01) ER = 17.62** | **-0.71 (-1.04 -0.4) ER = Inf** | **-0.45 (-0.83 -0.07) ER = 33.84** |  |
| Informed GE | **-0.32 (-0.66 0.01) ER = 17.76** | **-0.62 (-0.93 -0.33) ER = 4999** | -0.3 (-0.72 0.11) ER = 7.5 |  |

**S6.2 Effect of illness onset and duration**

To assess the role of illness onset and test whether vocal atypicalities are a long-term consequence of chronicity or already present at illness onset, we compared acoustic patterns between patients with first-episode (FES) schizophrenia and chronic patients. We used Bayesian multilevel regression models with each acoustic feature as outcome, illness onset (FES patients, chronic patients and healthy controls) and language (DK, CH, GE) as predictors. We separately assessed the effect of illness onset for each language, and modeled varying effects of participants, i.e., intercepts and slopes, separately for each group and language. For each acoustic feature we built a model with weakly informative priors, that is expectations of no differences between the groups. We can provide results for the comparison between patients with FES and chronic patients only for the Chinese and German corpus, while for the Danish and Japanese corpus, only patients with FES were included or information was missing. These are the ratios of patients with FES and not FES in the different corpora: DK (86 FES, 19 not reported (NR)), GE (22 FES, 39 chronic patients), CH (8FES, 23 chronic patients, 20 NR), JP (14 chronic patients).

To assess the role of duration of illness (DUI), we used Bayesian multilevel regression models with each acoustic feature as outcome and duration of illness as predictor. We separately assessed the effect of DUI for each language, and modeled varying effects of participants, i.e., intercepts and slopes, separately for each group and language. For each acoustic feature we built a model with weakly informative priors, that is expectations of no effect of DUI. This analysis was performed on the SCZ group only.

**Table S6_C.** Comparison between patients with first episode schizophrenia (FES) and chronic patients, and effect of duration of illness (DUI), on each acoustic feature

| **Acoustic features** | **Group** |  |  | **Duration of Illness** |
| --- | --- | --- | --- | --- |
| **Pitch Median** | FES – HC | Chronic – HC | Chronic – FES |  |
| Skeptical DK | -0.02 (-0.24 0.22) ER = 1.26 ER01 = 3.66 |  |  | 0.08 (-0.18 0.33) ER = 2.24 ER01 = 27.8 |
| Skeptical CH | 0.3 (-0.21 0.8) ER = 5.01 | **0.45 (0.09 0.81) ER = 48.26** | 0.15 (-0.43 0.74) ER = 1.94 ER01 = 1.78 | -0.02 (-0.06 0.02) ER = 4.36 |
| Skeptical GE | 0.11 (-0.19 0.43) ER = 2.51 ER01 = 2.35 | --0.15 (-0.4 0.11) ER = 4.89 | -0.25 (-0.62 0.1) ER = 7.38 | -0.01 (-0.04 0.01) ER = 5.49 |
| **Pitch IQR** |  |  |  |  |
| Skeptical DK | **-0.32 (-0.46 -0.19) ER = Inf** |  |  | **-0.12 (-0.26 0.02) ER = 12.53** |
| Skeptical CH | -0.19 (-0.57 0.21) ER = 3.78 | -0.18 (-0.47 0.1) ER = 6.22 | 0.01 (-0.43 0.44) ER = 1.03 ER01 = 2.7 | **0.03 (0 0.06) ER = 19.7** |
| Skeptical GE | **--0.36 (-0.58 -0.13) ER = 155.25** | -0.17 (-0.45 0.11) ER = 5.26 | 0.12 (-0.12 0.37) ER = 3.98 | 0.01 (-0.01 0.02) ER = 3.61 |
| **Speech Rate** |  |  |  |  |
| Skeptical DK | **-0.45 (-0.61 -0.29) ER = Inf** |  |  | 0 (-0.19 0.2) ER = 1.07 ER01 = 39.67 |
| Skeptical CH | 0.04 (-0.39 0.47) ER = 1.3 ER01 = 1.9 | -0.04 (-0.34 0.27) ER = 1.44 ER01 = 2.5 | -0.08 (-0.56 0.4) ER = 1.59 ER01 = 2.19 | **-0.03 (-0.06 0) ER = 17.66** |
| Skeptical GE | -0.11 (-0.42 0.19) ER = 2.49 ER01 = 2.32 | -0.12 (-0.37 0.13) ER = 3.77 | -0.01 (-0.35 0.33) ER = 1.08 ER01 = 3.35 | 0.01 (-0.01 0.03) ER = 3.38 |
| **Speech Percentage** |  |  |  |  |
| MA_Priors_ |  |  |  |  |
| Skeptical DK | -0.08 (-0.27 0.11) ER = 3.15 |  |  | -0.05 (-0.26 0.15) ER = 1.97 ER01 = 37.19 |
| Skeptical CH | 0.17 (-0.27 0.6) ER = 2.87 ER01 = 1.57 | **0.3 (-0.02 0.6) ER = 15.69** | 0.13 (-0.35 0.61) ER = 2.12 ER01 = 2.27 | **-0.03 (-0.06 0) ER = 12.12** |
| Skeptical JP |  |  |  |  |
| Skeptical GE | 0.08 (-0.23 0.39) ER = 2.03 ER01 = 2.33 | 0.03 (-0.23 0.3) ER = 1.31 ER01 = 3.01 | -0.05 (-0.42 0.31) ER = 1.47 ER01 = 2.88 | 0.01 (-0.01 0.03) ER = 3.28 |
| **Pause Number Min** |  |  |  |  |
| **Skeptical DK** | **-0.17 (-0.32 -0.02) ER = 32.11** |  |  | 0.03 (-0.13 0.2) ER = 1.7 ER01 = 48.71 |
| Skeptical CH | -0.29 (-0.7 0.13) ER = 6.76 | -0.08 (-0.37 0.21) ER = 2.11 ER01 = 2.53 | 0.2 (-0.29 0.68) ER = 3.18 | 0 (-0.03 0.04) ER = 1.42 ER01 = 260.38 |
| Skeptical GE | -0.02 (-0.28 0.25) ER = 1.25 ER01 = 2.85 | **-0.32 (-0.54 -0.1) ER = 100.01** | **-0.3 (-0.6 0) ER = 18.57** | 0.02 (0 0.03) ER = 15.18 |
| **Pause Duration** |  |  |  |  |
| Skeptical DK | **0.27 (0.11 0.42) ER = 302.03** |  |  | -0.02 (-0.19 0.15) ER = 1.33 ER01 = 53.76 |
| Skeptical CH | 0.03 (-0.37 0.41) ER = 1.21 ER01 = 2.07 | -0.09 (-0.36 0.19) ER = 2.37 ER01 = 2.5 | -0.11 (-0.55 0.32) ER = 2 ER01 = 2.35 | 0.02 (-0.01 0.05) ER = 7.54 |
| Skeptical GE | 0.09 (-0.19 0.38) ER = 2.35 ER01 = 2.62 | **0.21 (-0.03 0.45) ER = 11.97** | 0.12 (-0.21 0.46) ER = 2.53 ER01 = 2.89 | -0.01 (-0.03 0) ER = 7.06 |
| **Numbert of utterance** |  |  |  |  |
| MA_Priors |  |  |  |  |
| Skeptical DK | **-0.2 (-0.36 -0.05) ER = 69.42** | 0 (-0.82 0.81) ER = 0.99 ER01 = 1.03 | 0.2 (-0.63 1.02) ER = 1.93 ER01 = 1.38 | 0.04 (-0.13 0.2) ER = 1.78 ER01 = 45.04 |
| Skeptical CH | -0.27 (-0.7 0.18) ER = 5.27 | -0.08 (-0.38 0.22) ER = 1.95 ER01 = 2.62 | 0.19 (-0.32 0.68) ER = 2.86 ER01 = 1.89 | 0.01 (-0.03 0.04) ER = 1.68 ER01 = 241.85 |
| Skeptical GE | -0.05 (-0.31 0.22) ER = 1.61 ER01 = 2.98 | **-0.33 (-0.55 -0.11) ER = 137.89** | **-0.28 (-0.58 0.02) ER = 14.87** | **0.02 (0 0.03) ER = 17.18** |
| **Utterance Duration** |  |  |  |  |
| MA_Priors_High vs HC |  |  |  |  |
| Skeptical DK | **0.15 (-0.02 0.32) ER = 12.55** |  | -0.15 (-0.99 0.68) ER = 1.6 ER01 = 1.27 | -0.13 (-0.31 0.05) ER = 6.75 |
| Skeptical CH | **0.41 (-0.05 0.86) ER = 13.43** | **0.3 (-0.01 0.62) ER = 16.89** | -0.11 (-0.6 0.42) ER = 1.77 ER01 = 2.1 | -0.02 (-0.06 0.01) ER = 8.75 |
| Skeptical GE | 0.16 (-0.12 0.44) ER = 5.04 | **0.45 (0.21 0.7) ER = 832.33** | 0.28 (-0.06 0.65) ER = 9.36 | -0.01 (-0.03 0.01) ER = 3.1 |

**S6.3 Effect of Personal and Social Performance scale (PSP)**

To assess the relationship between the different acoustic features and the Personal and Social Performance scale (PSP), we built Bayesian multilevel regression models with each acoustic feature as outcome, and the PSP scale scores as ordinal predictors. We separately assessed the relationship between the different acoustic features and PSP for each language, and modeled varying effects of participants, i.e. intercepts and slopes, separately for each language. This analysis was performed on the SCZ group only.

**Table S6_D.** Association between scores on the Personal and Social Performance scale (PSP) and each acoustic feature

| **Acoustic Features** | **Personal and Social Performance scale (PSP)** |
| --- | --- |
| **Pitch Median** |  |
| Skeptical DK | -0.17 (-0.52 0.18) ER = 3.77 |
| Skeptical CH | 0.01 (-0.57 0.56) ER = 1.04 ER01 = 1.71 |
| **Pitch IQR** |  |
| Skeptical DK | 0.04 (-0.02 0.1) ER = 6.6 |
| Skeptical CH | 0.06 (-0.14 0.27) ER = 2.35 ER01 = 4.41 |
| **Speech Rate** |  |
| Skeptical DK | 0.11 (-0.05 0.27) ER = 6.66 |
| Skeptical CH | 0.25 (-0.17 0.67) ER = 5.51 |
| **Speech Percentage** |  |
| Skeptical DK | **0.16 (-0.04 0.37) ER = 10.09** |
| Skeptical CH | 0.21 (-0.27 0.67) ER = 3.32 |
| **Mean Pause Duration** |  |
| Skeptical DK | **-0.06 (-0.13 0.01) ER = 12.61** |
| Skeptical CH | -0.21 (-0.58 0.16) ER = 4.95 |
| **Pause Number** |  |
| Skeptical DK | 0.03 (-0.13 0.2) ER = 1.71 ER01 = 6.25 |
| Skeptical CH | -0.12 (-0.48 0.25) ER = 2.47 ER01 = 2.39 |
| **Mean Utterance Duration** |  |
| Skeptical DK | 0.1 (-0.04 0.25) ER = 6.46 |
| Skeptical CH | 0 (-0.06 0.07) ER = 1.15 ER01 = 14.66 |
| **Utterance Number** |  |
| Skeptical DK | 0.03 (-0.12 0.18) ER = 1.77 ER01 = 6.26 |
| Skeptical CH | -0.12 (-0.49 0.25) ER = 2.43 ER01 = 2.43 |

**S6.4 Relation between acoustic features and symptoms**

We here provide additional figures for summarizing the most important results on the relation between acoustic features and clinical symptoms reported in Table 3.

**Figure S6_B.** Estimated standardized relation between acoustic and clinical features. ER indicates the evidence ratio for the difference, ER01 the evidence ratio for the null effect. Single data points, regression line and 95% credible Bayesian intervals are represented in light blue for the Danish and in red for Chinese corpora. The outcomes, i.e., the acoustic features such as pitch variability, are represent on the y-axis, while the clinical symptom ratings (e.g., SANS-alogia) are represented on the x-axis.

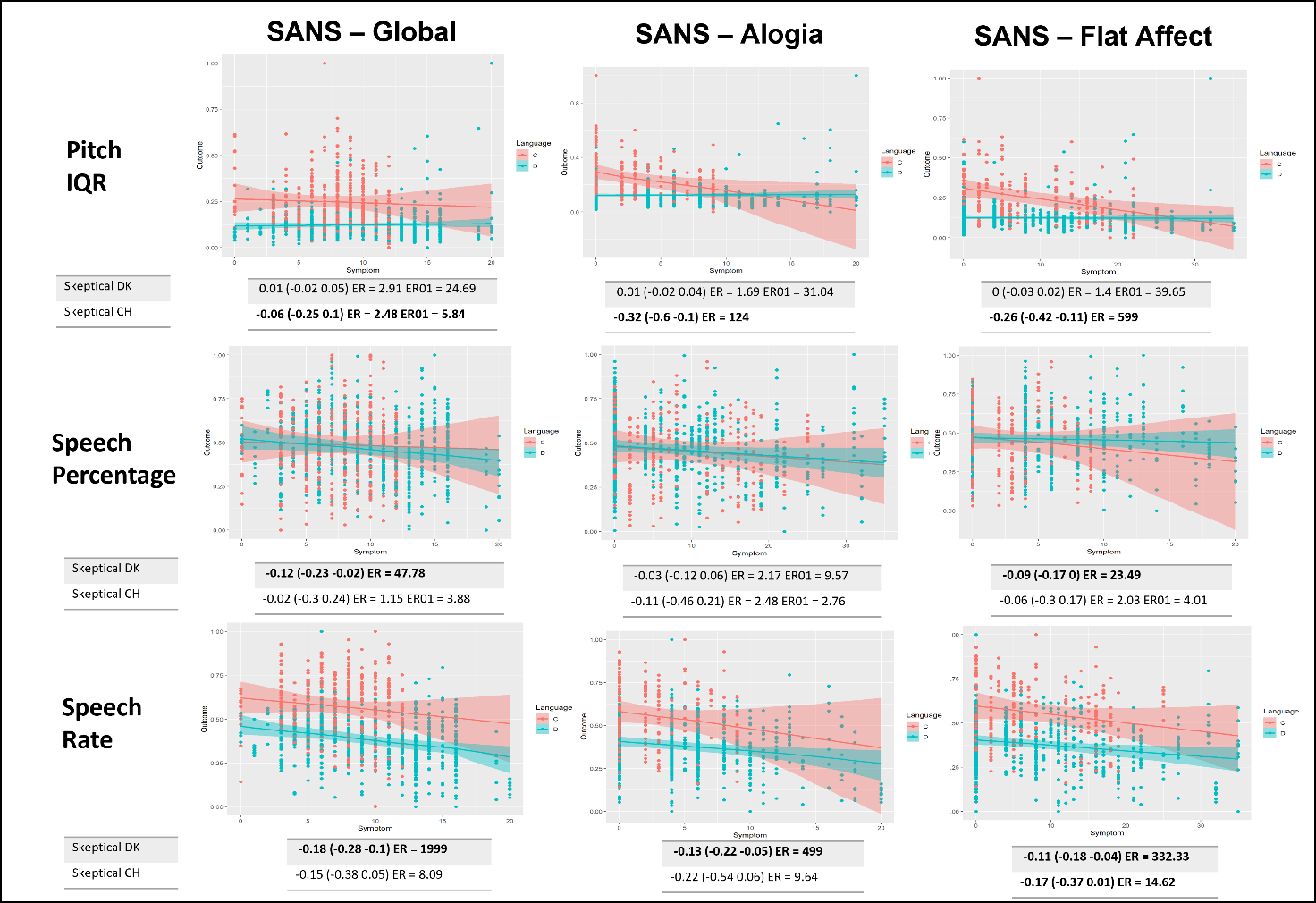

**Figure S6_C.** Estimated standardized relation between acoustic and clinical features. ER indicates the evidence ratio for the difference, ER01 the evidence ratio for the null effect. Single data points, regression line and 95% credible Bayesian intervals are represented in light blue for the Danish and in red for Chinese corpora. The outcomes, i.e., the acoustic features such as pitch variability, are represent on the y-axis, while the clinical symptom ratings (e.g., SANS-alogia) are represented on the x-axis.
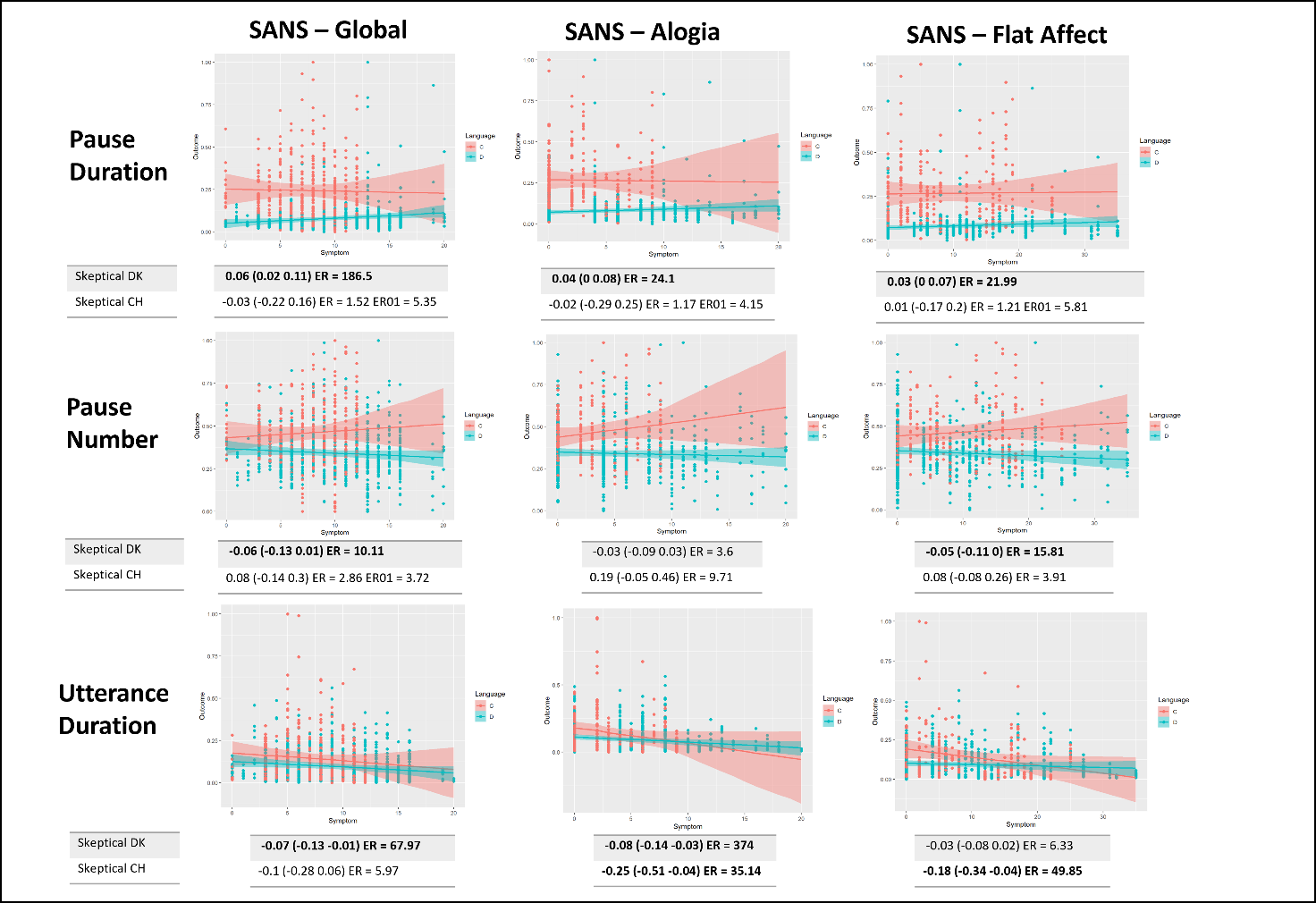

**S7 - Analyses on segments of 6 seconds**

In the analyses reported in the main manuscript, we focused on acoustic analyses of the full recordings of each single trial independently of their length. However, it could be argued that confounds might arise from comparing audios of different lengths. E.g. if a group produced consistently longer recordings, measures of variability might be accordingly increased, simply due to the longer time-series analyzed. To obviate that potential critique, we also systematically clipped all audio recordings in non-overlapping segments of 6 seconds, removing recordings that were too short and final segments that did not reach the full 6 seconds of length. This resulted in 5385 segments produced by patients with SCZ (2187 in Danish, 783 in Chinese, 243 in Japanese, and 1619 in German) and 6407 by controls (2582 in Danish, 854 in Chinese, 374 in Japanese, 1575 in German). We then repeated all analyses on these segments.

If we consider only trials with total duration <6sec, i.e. those trials in which participants provided very short answers to the task, we found that patients with SCZ produced short answers (<6sec) twice as often than controls (21% vs. 11%), resulting in 426 trials for patients with SCZ (218 for Danish, 103 for Chinese, 24 for Japanese, 81 for German) and 232 trials for controls (116 for Danish, 52 for Chinese, 4 for Japanese, 60 for German).

**S7.1 Acoustic features with meta-analytic results**

**Table S7_A.** Estimated standardized mean difference (HC- schizophrenia) for the eight acoustic measures present in the meta-analysis, as estimated separately by the meta-analytically informed and the skeptical models, including in the analysis only segments >=6 seconds

| **Acoustic Features** | **Group (CT – SCZ)** | **Group * Biological sex (M-F)** |
| --- | --- | --- |
| **Pitch Median** | MA priors = 0.25 (-0.72, 1.30) |  |
| Skeptical DK | 0.03 (-0.17 0.23) ER = 1.56 ER01 = 3.92 | 0.13 (-0.07 0.33) ER = 6.34 |
| Skeptical CH | **0.25 (-0.05 0.53) ER = 11** | **0.26 (-0.05 0.58) ER = 11.32** |
| Skeptical JP | **-0.55 (-0.97 -0.12) ER = 51.08** | **0.53 (0.06 0.99) ER = 28.33** |
| Skeptical GE | -0.05 (-0.28 0.17) ER = 1.67 ER01 = 3.18 | 0.07 (-0.27 0.42) ER = 1.71 ER01 = 3.16 |
| Informed DK | 0.06 (-0.13 0.26) ER = 2.33 ER01 = 4.23 | NA |
| Informed CH | 0.29 (0 0.56) ER = 20.14 | NA |
| Informed JP | -0.4 (-0.8 0.04) ER = 14.55 | NA |
| Informed GE | -0.01 (-0.23 0.21) ER = 1.1 ER01 = 4.01 | NA |
| *Stacking weight* | **Informed Model 1.000** |  |
| **Pitch IQR** | MA priors = **-0.55 (- 1.06, 0.09)** |  |
| Skeptical DK | **-0.13 (-0.27 0) ER = 19.62** | 0.12 (-0.13 0.37) ER = 3.93 |
| Skeptical CH | **-0.06 (-0.27 0.16) ER = 2.02 ER01 = 3.43** | -0.04 (-0.44 0.37) ER = 1.24 ER01 = 3.43 |
| Skeptical JP | **-0.36 (-0.63 -0.08) ER = 56.14** | 0.22 (-0.37 0.79) ER = 2.95 ER01 = 1.59 |
| Skeptical GE | **-0.23 (-0.38 -0.07) ER = 100.01** | -0.01 (-0.28 0.27) ER = 1.06 ER01 = 4.22 |
| Informed DK | **-0.19 (-0.31 -0.06) ER = 118.05** | NA |
| Informed CH | **-0.19 (-0.39 0) ER = 18.46** | NA |
| Informed JP | **-0.46 (-0.69 -0.23) ER = 1999** | NA |
| Informed GE | **-0.28 (-0.44 -0.14) ER = 2499** | NA |
| *Stacking weight* |  |  |
| **Speech Rate** | MA priors -0.75 (-1.51, 0.04) |  |
| Skeptical DK | **-0.33 (-0.46 -0.2) ER = Inf** | 0.14 (-0.12 0.39) ER = 4.19 |
| Skeptical CH | 0.01 (-0.19 0.22) ER = 1.17 ER01 = 4.05 | -0.01 (-0.4 0.37) ER = 1.09 ER01 = 4.05 |
| Skeptical JP | 0.12 (-0.25 0.49) ER = 2.59 ER01 = 2.03 | -0.3 (-0.99 0.38) ER = 3.24 |
| **Skeptical GE** | **-0.15 (-0.33 0.03) ER = 11.21** | 0.22 (-0.16 0.59) ER = 5.02 |
| Informed DK | **-0.37 (-0.5 -0.24) ER = Inf** | NA |
| Informed CH | -0.08 (-0.28 0.12) ER = 2.88 ER01 = 21.05 | NA |
| Informed JP | -0.14 (-0.49 0.2) ER = 2.83 ER01 = 13.56 | NA |
| Informed GE | -**0.21 (-0.38 -0.03) ER = 30.95** | NA |
| *Stacking weight* |  |  |
| **Speech Percentage** | MA priors -1.26 (-2.26, 0.25) |  |
| Skeptical DK | **-0.17 (-0.33 -0.01) ER = 23.94** | -0.11 (-0.42 0.19) ER = 2.63 ER01 = 3.13 |
| Skeptical CH | **0.29 (0.05 0.54) ER = 35.23** | -0.18 (-0.65 0.29) ER = 2.83 ER01 = 0.47 |
| Skeptical JP | 0.28 (-0.1 0.65) ER = 7.9 | -0.29 (-0.96 0.39) ER = 3.19 |
| Skeptical GE | -0.04 (-0.26 0.17) ER = 1.56 ER01 = 3.55 | 0.3 (-0.12 0.72) ER = 7.36 |
| Informed DK | **-0.21 (-0.38 -0.05) ER = 67.03** | NA |
| Informed CH | 0.16 (-0.1 0.42) ER = 5.87 | NA |
| Informed JP | -0.03 (-0.44 0.35) ER = 1.19 ER01 = 58 | NA |
| Informed GE | -0.12 (-0.31 0.08) ER = 5.03 | NA |
| Stacking weight |  |  |
| **Pause Number** | MA priors 0.05 (-1.23, 1.13) |  |
| Skeptical DK | **-0.32 (-0.45 -0.19) ER = 9999** | **0.4 (0.13 0.66) ER = 146.06** |
| Skeptical CH | **-0.29 (-0.5 -0.08) ER = 79.65** | -0.29 (-0.71 0.13) ER = 6.59 |
| Skeptical JP | -0.07 (-0.39 0.26) ER = 1.77 ER01 = 2.34 | -0.27 (-0.93 0.41) ER = 2.96 ER01 = 1.46 |
| Skeptical GE | **-0.3 (-0.46 -0.13) ER = 554.56** | 0.15 (-0.17 0.48) ER = 3.4 |
| Informed DK | **-0.32 (-0.44 -0.19) ER = 9999** | NA |
| Informed CH | **-0.29 (-0.5 -0.08) ER = 75.92** | NA |
| Informed JP | -0.06 (-0.38 0.26) ER = 1.66 ER01 = 2.35 | NA |
| Informed GE | **-0.29 (-0.45 -0.13) ER = 525.32** | NA |
| Stacking weight |  |  |
| **Pause duration** | MA priors 1.89 (0.72, 3.21) |  |
| Skeptical DK | **0.3 (0.16 0.43) ER = 9999** | -0.05 (-0.34 0.25) ER = 1.53 ER01 = 3.91 |
| Skeptical CH | 0.02 (-0.14 0.17) ER = 1.38 ER01 = 5.19 | 0.11 (-0.19 0.41) ER = 2.5 ER01 = 5.19 |
| Skeptical JP | -0.21 (-0.48 0.07) ER = 8.64 | 0.36 (-0.27 1) ER = 4.68 |
| Skeptical GE | **0.19 (0.07 0.3) ER = 199** | -0.15 (-0.4 0.11) ER = 4.81 |
| Informed DK | **0.34 (0.21 0.48) ER = Inf** | NA |
| Informed CH | 0.07 (-0.08 0.22) ER = 3.55 | NA |
| Informed JP | -0.02 (-0.31 0.28) ER = 1.26 ER01 = 835.82 | NA |
| Informed GE | **0.22 (0.1 0.34) ER = 832.33** | NA |
| Stacking weight |  |  |
| **Turn Duration** | MA priors -0.13 (-1.571 1.342) |  |
| Skeptical DK | **0.13 (0 0.26) ER = 17.05** | **-0.39 (-0.65 -0.13) ER = 106.53** |
| Skeptical CH | **0.37 (0.02 0.7) ER = 22.58** | -0.16 (-0.69 0.37) ER = 2.31 ER01 = 0.5 |
| Skeptical JP | **0.64 (0.34 0.94) ER = 1110.11** | -0.02 (-0.59 0.57) ER = 1.11 ER01 = 1.93 |
| Skeptical GE | **0.27 (0.1 0.45) ER = 332.33** | 0.14 (-0.23 0.52) ER = 2.73 ER01 = 2.39 |
| Informed DK | **0.13 (0 0.27) ER = 17.98** | NA |
| Informed CH | **0.41 (0.04 0.79) ER = 29.4** | NA |
| Informed JP | **0.69 (0.36 1) ER = 713.29** | NA |
| Informed GE | **0.28 (0.11 0.45) ER = 199** | NA |
| Stacking weight |  |  |
| **Turn Number** |  |  |
| Skeptical DK | **-0.21 (-0.35 -0.07) ER = 171.41** | **0.37 (0.1 0.64) ER = 76.52** |
| Skeptical CH | -0.14 (-0.37 0.1) ER = 4.93 | **-0.37 (-0.81 0.05) ER = 12.12** |
| Skeptical JP | -0.1 (-0.48 0.27) ER = 2.12 ER01 = 1.99 | -0.34 (-1.02 0.33) ER = 3.78 |
| Skeptical GE | **-0.23 (-0.43 -0.03) ER = 29.21** | 0.18 (-0.22 0.58) ER = 3.37 |

**S8 – Software Implementation notes**

Bayesian meta-analyses were calculated using the brms^36^ R interface for Stan^37^. Figures were produced with ggplot2 from the tidyverse package^38^. All analyses were conducted in R using the RStudio IDE^18^. The source code for the analysis is openly available and can be found at: https://osf.io/h4wj6/.

**S9 - A cumulative yet self-correcting approach.**

In this study we showcased a cumulative yet self-critical scientific approach. Current debates contrast a cumulative approach (“standing on the shoulders of giants”) that systematically relies on previous findings (e.g. via systematic reviews and meta-analyses) with the potential unreliability of those very same findings due to questionable research practices, publication bias and other issues^39–41^. In this study, we account for both sides. We rely on a previous systematic review to design the current study, and on the meta-analytic findings to set up priors for our analyses. At the same time, we critically attempt to replicate and generalize previous findings, and compare meta-analytically informed statistical inferences with inferences relying on skeptical priors.

Meta-analytically informed models were not generally more robust and generalizable to new data (less than half of the informed models presents LOO weights above .75, see **Table 2**). In particular, including the meta-analytic findings in our analyses improved the generalizability of our inferences for pitch median, pitch duration, and pause duration; for the remaining models, we instead found that skeptic models performed better than the informed ones, or no difference (See **Figure 1**). This may have different interpretations.

First, these findings are in agreement with the inconsistent replications we found in our results. Indeed, the most prominent results in the meta-analysis (duration features) were not found consistently in our study, and accordingly the usefulness of including meta-analytic informed priors for those measures was limited.

Second, previous meta-analytic estimates were highly unreliable as indicated by the large confidence intervals and high heterogeneity characterizing the estimates, and afflicted by publication bias. Furthermore, we found that the meta-analytic effect sizes tend to vary in relationship with the speech production task used; however, given the very limited number of studies for each speech task category (see **SM4)** and considering that for most of the acoustic features the estimates were not available for all the different speech task categories, we here used the global summary effect size (not divided by task category) for each acoustic feature.

Summing up, these findings indicate that previous studies were not fully representative of our setup (task, and sample), perhaps due to differences in methods, or samples (insufficient sample sizes, heterogeneity of the patient population covered, cultural and linguistic differences, etc.), perhaps due to questionable research practices; but more likely due to a combination of all these factors.

Our results thus suggest that posterior passing, especially when previous meta-analysis suggest the presence of noise and heterogeneity, should not be acritically applied. Nevertheless, even in these conditions, we argue that comparatively applying meta-analytically informed and skeptical priors might provide a check of the inferential robustness (do the results agree across the two models?) **and a measure** of how the current study relates to the previous state of the art of the literature (how well does it fall within the range of the previous studies, or how radically does it suggest that our current study is saying something different?).

While the application of this approach raises some questions on generalizability of previous results, it also suggests a possible future way for increasing replicability and generalizability of previous results. For example, by relying on the suggestions of previous meta-analysis, we found that accounting for the heterogeneity of the disorder (socio-demographical, clinical, linguistic) is crucial. Furthermore, our results suggest the importance of assessing for task-specific effects, not collating too heterogeneous effects. Also, they indicate the need of larger corpora for testing the reliability and generalizability (e.g., out of sample predictability) of voice analysis results and assessing its clinical applicability.
